# The COVID-19 Pandemic as an Opportunity for Unravelling the Causative Association between Respiratory Viruses and Pneumococcus-Associated Disease in Young Children: A Prospective Study

**DOI:** 10.1101/2022.09.06.22279606

**Authors:** Ron Dagan, Bart Adriaan van der Beek, Shalom Ben-Shimol, David Greenberg, Yonat Shemer-Avni, Daniel M. Weinberger, Dana Danino

## Abstract

**BACKGROUND:** In young children, rates of lower respiratory infections (LRI) and invasive pneumococcal disease (IPD) have been associated with respiratory syncytial virus (RSV), human metapneumovirus (hMPV), influenza (flu), and parainfluenza (PIV) (collectively termed here as pneumonia and pneumococcal disease-associated viruses [PDA-viruses]). However, their contribution to the pathogenesis of these disease endpoints has not yet been elucidated. The COVID-19 pandemic provided a unique opportunity to examine the question.

**METHODS:** This prospective study comprised all children <5 years, living in southern Israel, during 2016 through 2021. The data were previously collected in multiple ongoing prospective surveillance programs and include: hospital visits for community-acquired alveolar pneumonia (CAAP), non-CAAP LRI; nasopharyngeal pneumococcal carriage (<3 years of age); respiratory virus activity; and nationwide, all-ages COVID-19 episodes and IPD in children <5 years. A hierarchical statistical model was developed to estimate the proportion of the different clinical endpoints attributable to each virus from monthly time series data, stratified by age and ethnicity. A separate model was fit for each endpoint, with covariates that included a linear time trend, 12-month harmonic variables to capture unexplained seasonal variations, and the proportion of tests positive for each virus in that month.

**FINDINGS:** During 2016 through 2021, 3,204, 26,695, 257, and 619 episodes of CAAP, non-CAAP LRI, pneumococcal bacteremic pneumonia and non-pneumonia IPD, respectively, were reported. Compared to 2016-2019, broad declines in the disease endpoints were observed shortly after the pandemic surge, coincident with a complete disappearance of all PDA-viruses and continued circulation of rhinovirus (RhV) and adenovirus (AdV). From April 2021, off-season and abrupt surges of all disease endpoints occurred, associated with similar dynamics among the PDA-viruses, which re-emerged sequentially. Using our model fit to the entire 2016-2021 period, 82% (95% CI, 75-88%) of CAAP episodes in 2021 were attributable to the common respiratory viruses, as were 22%-31% of the other disease endpoints. Virus-specific contributions to CAAP were: RSV, 49% (95% CI, 43-55%); hMPV, 13% (10-17%); PIV, 11% (7-15%); flu, 7% (1-13%). RhV and AdV did not contribute. RSV was the main contributor in all endpoints, especially in infants. Pneumococcal carriage prevalence remained largely stable throughout the study.

**INTERPRETATION:** RSV and hMPV play a critical role in the burden of CAAP and pneumococcal disease in children. Interventions targeting these viruses could have a secondary effect on the burden of disease typically attributed to bacteria.

**FUNDING:** There was no funding for this study.

**Research in Context:** *Evidence before this study:* Lower respiratory infections (LRI) and invasive pneumococcal disease (IPD) in young children, have often been associated with specific respiratory viruses, namely respiratory syncytial virus (RSV), human metapneumovirus (hMPV) influenza viruses (flu), and parainfluenza viruses (PIV) (termed in the current article pneumonia and pneumococcal disease-associated viruses [PDA-viruses]). However, their causative role as co-pathogens has not yet been fully elucidated. There is already ample evidence that bacteria and viruses interact to cause severe disease. This could be seen after the introduction of pneumococcal conjugate vaccines (PCVs), when there was a significant reduction in hospitalisation for viral lower respiratory infections (LRIs). This suggests that viral-pneumococcal coinfections are common and play a role in the pathogenesis of pneumococcal respiratory infections. To demonstrate the contribution of viruses to the burden of pneumococcal disease specifically, and pneumonia in general, it would be necessary to eliminate one or more of the respiratory viruses. Shortly after the start of the COVID-19 pandemic, multiple reports demonstrated reduced IPD and LRI rates among young children, coincident with dramatically reduced rates of the PDA-viruses globally. Initially, the reduced rates of pneumococcal disease were attributed to non-pharmaceutical interventions that might reduce pneumococcal transmission in the community. However, continuous, virtually unchanged pneumococcal carriage rates were reported in multiple studies, strongly suggesting the reduced circulation of *S. pneumoniae* was not significantly contributing to disease reduction. Surprisingly, pneumococcus-associated diseases and PDA-viruses simultaneously re-emerged in 2021 during the off-season. In contrast to PDA-viruses, other viruses, such as adenovirus and rhinovirus did not show any of the patterns discussed above. We searched PubMed on June 1^st^, 2022, for studies since 2020 using the following terms: (“COVID-19” or “SARS-Cov-2”) and (“*S. pneumoniae*” or “pneumococcus” or “IPD” or “respiratory virus” or respiratory syncytial virus” or “hMPV” or “influenza” or “parainfluenza” or “adenovirus” or “rhinovirus” or “lower respiratory infection”). The search was for English literature and unrestricted by date.

*Added value of this study:* Three unique characteristics of the COVID-19 pandemic-induced abnormal dynamics, coupled with multiple ongoing cohort studies in young children, contributed to the historic opportunity to model and quantify the attributable role of the various common respiratory viruses to four pneumococcus-associated disease endpoints (in particular community-acquired alveolar pneumonia (CAAP), non-CAAP LRIs, pneumococcal bacteremic pneumonia and non-pneumonia IPD): First, the full seasonal disappearance of all PDA-viruses shortly after the start of the pandemic, in the presence of continuous, uninterrupted pneumococcal carriage and continuous unchanged rhinovirus and adenovirus activity. Second, the off-season resurgence of the PDA-viruses in 2021. Third, the sequential, rather than simultaneous, re-emergence of the PDA-viruses. The analysis in this study suggests that several of the respiratory viruses, particularly RSV and hMPV, play an important causative role in the pathogenesis of pneumococcal diseases and other respiratory infections. Furthermore, the proportion attributable to each of the PDA-viruses for each of the four studied disease endpoints, and each of the age groups (<1, 1, and 2-4 years of age) could be estimated.

*Implication of all the available findings:* Our findings add evidence about the absolute and relative contribution of common respiratory viruses to the burden of pneumonia and pneumococcal diseases and related conditions in young children. The strong contribution of RSV to disease burden compared to other viruses in all studied disease endpoints suggests that interventions that target viruses could have secondary effects on the burden of diseases typically attributed to bacteria.

## Introduction

Respiratory viruses are major contributors to the burden of pneumonia in general and disease caused by pneumococcus specifically. Respiratory syncytial virus (RSV), human metapneumovirus (hMPV), influenza viruses (flu), and parainfluenza viruses (PIV) have been associated with both community-acquired alveolar pneumonia (CAAP; considered mostly bacterial, in particular pneumococcal),^1, 2^ and invasive pneumococcal disease (IPD).^1-12^ However, for other common viruses, such as adenoviruses (AdV) and rhinoviruses (RhV), there is little evidence of a causal association with pneumonia or pneumococcal disease in young children.^13^

There is compelling evidence that pneumococcus, a bacterial pathogen, contributes to the burden of viral pneumonia and non-pneumonia lower respiratory tract infections (LRIs) in young children. This was seen in declining rates of hospitalisation for viral pneumonia following use of pneumococcal conjugate vaccines (PCVs), both in clinical trials and observational studies^10, 14, 15^ This supports the notion that pneumococcus may increase the severity of viral infections. However, there is limited evidence about whether reducing or eliminating the activity of specific viruses could reduce severity or rates of pneumonia or pneumococcal disease. The COVID-19 pandemic provided a unique opportunity to examine this question.

Shortly after the emergence of the COVID-19 pandemic, temporary elimination of the circulation of RSV, hMPV, flu, and PIV occurred globally, while RhV and AdV continued to circulate.^13, 16^ At the same time, a drop in pneumococcal disease rates was seen.^17-21^ This was initially believed to result from reduced pneumococcal circulation, caused by the mitigation measures adopted during the COVID-19 pandemic. However, nasopharyngeal carriage rates of pneumococcus among young children were largely unchanged during the pandemic, suggesting that this was not the main factor behind the decline in rates of pneumococcal disease.^12, 13, 16, 22^ An alternative possibility, based on previous observations of bacterial-viral coinfections, is that the dramatic suppression of the respiratory viruses led to the decline in rates of IPD.

Starting in 2021, an off-season sequential re-emergence of the suppressed viruses occurred in Israel, starting with PIV, and followed by hMPV, RSV, and flu. This provided a unique opportunity to examine the association of these viruses with rates of community acquired alveolar pneumonia (CAAP), IPD and non-CAAP LRIs. We hypothesized that RSV, hMPV, flu, and PIV (collectively termed in the current study pneumonia and pneumococcus disease-associated viruses [PDA-viruses]) are important determinants of these disease endpoints in young children. In a previous study we described events that occurred from the start of the pandemic in 2020 through February 2021, mainly describing the period when the PDA-viruses were absent. The current study extends the period to the end of 2021, which includes the sequential re-emergence of the PDA-viruses and pneumococcus-associated disease and adds a model to quantify the respective role of individual viruses in the dynamics of each of the four studied endpoints by age group. This study provides a comprehensive picture of the unusual dynamics of the respiratory viruses, pneumococcal disease, and other respiratory disease endpoints.

## Methods

### Setting

The Soroka University Medical Center (SUMC) is the only hospital in the Negev, southern Israel, providing primary health care to >95% of the children living in the region, enabling incidence calculations. Two distinct paediatric ethnic populations reside side-by-side in southern Israel: The Bedouin population, with infectious disease epidemiology, including pneumococcal serotype distribution, similar to lower-middle income countries; and the Jewish population, whose epidemiology resembles higher-income Western countries. Higher rates of respiratory and invasive disease, and pneumococcal carriage were reported among Bedouin than among Jewish children.^23^ In 2020, there were ∼96,500 children <5 years in the Negev; ∼50% were Bedouin.^24^

Implementation of PCVs in the National Immunization Plan occurred in 2009 (PCV7) and 2010 (PCV13); since 2013 ∼90% of both Jewish and Bedouin children have received all three required doses.^25^

### The COVID-19 pandemic in 2019-2021

The details of the COVID-19 pandemic and associated mitigation measures are described in **Supplementary Figure 1**.

### Study design

The study population for respiratory disease and virus-testing comprised all children living in the region from January 2016 through December 2021. For IPD, we included all children nationwide. All disease databases included children <5 years old. However, for pneumococcal carriage, we included children <3 years since the complete database of children without respiratory symptoms has been created only for children in this age group. The data were derived from multiple ongoing, prospective long-standing cohort surveillance programs.

Monthly incidence rates of the various disease endpoints and prevalence rates of pneumococcal nasopharyngeal carriage and detection of respiratory viruses during 2020-2021 were compared to the corresponding values during 2016-2019. The total numbers for each database and the comparison of demographic characteristics are presented in **Table 1**.

**Table 1:** Demographics of the study population.

|  | **Total** | **2016-2019** | **2020-2021** |
| --- | --- | --- | --- |
| **Number of samples obtained for virus detection** |  |  |  |
| Cases | 27,138 | 17,555 | 9,583 |
| Age (months) |  |  |  |
| mean±SD | 14·9±14·9 | 14·5±14·8 | 15·5±15·1 |
| median | 9·5 | 9·1 | 10·3 |
| Ethnicity n (%) |  |  |  |
| Jewish | 9,200 (33·9%) | 6,102 (34·8%) | 3,098 (32·3%) |
| Bedouin | 17,938 (66·1%) | 11,453 (65·2%) | 6,458 (67·7%) |
| **Carriage** |  |  |  |
| Cases | 10,434 | 7,921 | 2,513 |
| Age (months) |  |  |  |
| mean±SD | 11·1±9·2 | 11·3±9·3 | 10·6±9·0 |
| median | 8·5 | 8·8 | 7·7 |
| Ethnicity n (%) |  |  |  |
| Jewish | 4,724 (45·3%) | 3,615 (45·6%) | 1,109 (44·1%) |
| Bedouin | 5,710 (54·7%) | 4,306 (54·4%) | 1,404 (55·9%) |
| **CAAP** |  |  |  |
| Cases | 3,204 | 2,286 | 918 |
| Age (months) |  |  |  |
| mean±SD | 17·4±14·6 | 17·1±14·7 | 18·0±14·5 |
| median | 13·3 | 12·7 | 14·8 |
| Ethnicity n (%) |  |  |  |
| Jewish | 1,259 (39·3%) | 871 (38·1%) | 388 (42·3%) |
| Bedouin | 1,945 (60·7%) | 1,415 (61·9%) | 530 (57·7%) |
| **Non-CAAP LRI** |  |  |  |
| Cases | 26,695 | 17,349 | 9,346 |
| Age (months) |  |  |  |
| mean±SD | 17·7±14·9 | 17·3±14·7 | 18·4±15·3 |
| median | 13·0 | 12·6 | 13·9 |
| Ethnicity n (%) |  |  |  |
| Jewish | 11,143 (41·7%) | 7,377 (42·5%) | 3,766 (40·3%) |
| Bedouin | 15,552 (58·3%) | 9,972 (57·5%) | 5,580 (59·7%) |
| **Pneumococcal Bacteremic Pneumonia\*** |  |  |  |
| Cases | 257 | 193 | 64 |
| Age (months) |  |  |  |
| mean±SD | 20·5±13·3 | 20·9±13·6 | 19·3±12·1 |
| median | 15·7 | 16·5 | 15·1 |
| Ethnicity n (%) |  |  |  |
| Jewish | 220 (85·6%) | 167 (86·5%) | 53 (82·8%) |
| Non-Jewish | 37 (14·4%) | 26 (13·5%) | 11 (17·2%) |
| **Non-pneumococcal IPD\*** |  |  |  |
| Cases | 619 | 434 | 185 |
| Age (months)mean±SD |  |  |  |
| mean±SD | 15·8±12·2 | 15·2±11·8 | 17·2±12·9 |
| median | 12·9 | 12·6 | 13·9 |
| Ethnicity n (%) |  |  |  |
| Jewish | 489 (79·0%) | 346 (79·7%) | 143 (77·3%) |
| Non-Jewish | 130 (21·0%) | 88 (20·3%) | 42 (22·7%) |

\* The cases are from the nationwide database, in contrast to all others which refer to southern Israel databases. Therefore, since the Bedouin population is concentrated almost exclusively in southern Israel, the ethnic division from the nationwide IPD study is Jewish vs. Non-Jewish, rather than Jewish vs. Bedouin. The non-Jewish population nationwide is almost exclusively Arab population and also includes the Bedouin population from southern Israel.

#### 1) Surveillance of community-acquired alveolar pneumonia (CAAP) and non-CAAP LRI requiring chest-radiography in children <5 years of age, southern Israel

This is an ongoing, prospective, population-based, active surveillance system, initiated in 2002. The current report deals with data from January 2016 through December 2021.

All children <5 years residing in the Negev district, seen at the SUMC paediatric emergency room (PER) (either ambulatory or hospitalised) who had chest radiological examination (CXR) within <48 hours from admission were included in the analysis.

Clinical practice in the SUMC PER regarding LRI referral, evaluation, and the need for CXR examination did not change throughout the study period. Chest radiographs were analysed according to the World Health Organization (WHO) Standardization of Interpretation of Chest Radiograph Working Group.^26, 27^ All chest radiographs were recorded daily and were evaluated separately by the 2 paediatric infectious disease specialists who each read all the chest radiographs, independently. Further evaluation was done by a paediatric radiologist who was unaware of the clinical data and of the paediatricians’ evaluation. CAAP diagnosis was confirmed by agreement on the presence of alveolar infiltrates or pleural effusion of at least one of the study paediatric infectious disease specialists and the paediatric radiologist.^28^ Since November 2020 the paediatric radiologist was replaced by a third paediatric infectious disease specialist. For this period the diagnosis of CAAP needed agreement of at least two of the three readers.

The diagnosis of non-CAAP LRI included all visits of LRI with CXR examination that did not show alveolar infiltrates. This group included both: 1) CXR with readings of infiltrates that were not alveolar (<5% of the group), including linear and patchy densities (interstitial infiltrates) in a lacy pattern involving both lungs, featuring peri bronchial thickening and multiple areas of atelectasis; or minor patchy infiltrate that are not of sufficient magnitude to constitute alveolar pneumonia, and small areas of atelectasis;^26^ and 2) absence of consolidation, other infiltrates or pleural effusion (but including CXR with hyperinflation, >95% of this group). All non-CAAP-LRIs were grouped together because previous studies showed a very low inter-reader consistency.^29^

#### 2) Nationwide Invasive Pneumococcal Diseases (IPD) Surveillance

This has been an ongoing, nationwide, prospective, population-based, active surveillance system, initiated in 1989. Surveillance is conducted by the Israeli Pediatric Bacteremia and Meningitis Group (IPBMG) in 27 medical centres routinely obtaining blood cultures from children: All 26 hospitals admitting children and one major outpatient health maintenance organisation (HMO, Maccabi Healthcare Services) central laboratory. Less than 1% of blood cultures in all ages and no cerebrospinal fluid (CSF) cultures are obtained outside these centres.

IPD episodes were defined as illness episodes during which *S. pneumoniae* was isolated from blood or CSF. Study population: All children <5 years in Israel. Non-culture diagnoses (polymerase chain reaction [PCR], antigen testing, gram stain or clinical diagnosis only) were excluded. Positive cultures from non-blood or CSF sterile sites (i.e., joint/pleural fluid) were excluded. Local investigators in each centre responded to a questionnaire sent monthly by the principal investigator, located at the Pediatric Infectious Disease Unit (PIDU), of the SUMC, which served as the study headquarters. Several measures were used to ensure consistent data collection; (1) weekly contact with all 27 laboratories; (2) assuring transportation of all *S. pneumoniae* isolates to the study headquarters’ laboratory; and (3) monthly contact with the local investigators. Completed reports included the following data: Isolate source (blood/CSF), culture date, birthdate, sex, ethnicity (Jewish/non-Jewish), main diagnoses, endpoint (mortality) and hospitalisation duration. IPD cases reported from each site have been constantly compared to the list of isolates obtained by the reference Ministry of Health laboratory. Since July 2009, >95% of the cases have been retrieved.^30, 31^

#### 3) Pneumococcal nasopharyngeal (NP) carriage surveillance

The detailed nasopharyngeal testing methodology was recently described.^25^ Nasopharyngeal samples were obtained using a flexible dacron-tipped swab, introduced through the nostrils and advanced until resistance was found. These swabs were inoculated into modified Stewart transport medium (Medical Wire and Equipment Co., Ltd., Corsham, England) and processed within 16 hours at the SUMC Clinical Microbiology Laboratory. Material from swabs was plated on Columbia agar with 5% sheep blood and 5.0 μg/mL gentamicin and incubated aerobically at 35°C in a CO_2_-enriched atmosphere for 48h. The presumptive identification of *S. pneumoniae* was based on the presence of α-hemolysis and the inhibition by optochin, and the identity of the bacteria present was confirmed by a positive slide agglutination test result (Phadebact; Pharmacia Diagnostics). One *S. pneumoniae* colony per plate was then subcultured, harvested, and kept frozen at -70°C for further testing. Density analysis was performed at the time of initial sample processing. A semiquantitative plating method was used and graded 0 (negative) to 4 (growth in all plate quadrants). The NP swabs were plated by rolling the swab over one quarter of the plate and streaking the sample onto 4 quadrants using a sterile loop. Growth was termed 1+, 2+, 3+, or 4+ when colonies were seen in 1, 2, 3, or 4 quadrants, respectively.^32^

##### a) Children <3 years without respiratory infections visiting the SUMC PER

This is an ongoing, prospective, population-based, active surveillance system, initiated in November 2009. Each working day, nasopharyngeal cultures were obtained from the first 4 Jewish and 4 Bedouin children <3 years old, resident of the Negev region, presenting at the PER, after a written informed consent was signed by the parents. Children not residing in the Negev region were excluded. In cases where multiple swabs were obtained from a child during the same month, only the first culture was included^25^ If <8 children were available, nasopharyngeal carriage evaluation was obtained from randomly selected hospitalized children. For the current analysis, we excluded those with any diagnosis of respiratory infections. These children constituted 3114/7921 (34·3%) of tested children during 2016-2019. In 2020 and 2021 these children constituted 331/1190 (27·8%) and 457/1323 (34·5%), respectively, of all children tested for carriage.

##### b) Healthy children <3 years presenting for vaccinations

Since 2011, nasopharyngeal cultures have been obtained from healthy children <3 years old, presenting to the Maternal and Child Health Care Centres in southern Israel for vaccination. A nasopharyngeal swab was obtained after a written informed consent was signed by the parents.^33^ These children constituted 4,807/7,921 (60·7%) of all tested children in 2016-2019. In 2020 and 2021 they constituted 859/1,190 (72·2%) and 866/1,323 (65·5%), respectively.

#### 4) Nasopharyngeal detection of respiratory viruses

Nasopharyngeal specimens for respiratory viruses were obtained from hospitalised children following the request of the treating physician for clinical indication. Until February 2019 the specimens were nasopharyngeal washes and the procedures, including the laboratory method for PCR testing, were previously published.^34^ The viruses tested were RSV, influenza A and influenza B viruses, parainfluenza viruses, adenovirus and human metapneumovirus. Since February 2019, nasal swabs were obtained and the detection method was switched to a commercial kit (Seeplex RV7 detection kit, Seengene Inc., Korea).

## Ethics

The study and all its components were approved by the SUMC Ethics Committee with the following approval numbers: SOR-0405-16; SOR-0220-20; SOR-0062-11; 10374; 4908, 3075.

## Statistical Analysis

Since respiratory infection rates, relative importance of viruses and clinical presentations differ by age and ethnic group, monthly incidence rates for all endpoints were stratified by age (<1, 1, 2-4 years) and ethnicity (Bedouin, Jewish). Viruses were divided into 2 groups: PDA-viruses (RSV, hMPV, flu, and PIV) and non-PDA-viruses (RhV and AdV). For the modelling we analysed each virus separately, even if they were detected as coinfections. The monthly proportion of positive pneumococcal nasopharyngeal cultures were calculated out of all collected samples for children <1 and 1-2 years, except during April-May 2020, due to disruptions in sample collection caused by a strict lockdown. No difference in the age distribution of the population <5 was observed during the study years. Ratios of incidence rates and proportions were calculated to compare the COVID-19 and pre-COVID-19 periods. All rate ratios were adjusted for ethnicity and age using the Cochran-Mantel-Haenszel method. The statistical significance threshold was p <0·05. Data were analysed using R 4.0.2.^35^

### Modelling

The goal was to estimate the proportion of each disease endpoint attributable to the different viruses. To do this, we developed hierarchical Bayesian regression models with a negative binomial likelihood and identity link.^36^ The clinical outcomes evaluated were CAAP, non-CAAP LRI, bacteremic pneumococcal pneumonia and non-pneumonia IPD. For each endpoint, we have a time series of the number of clinical cases, by month, stratified by age category (<1, 1, 2-4 years) and ethnicity (Jewish *vs*. Bedouin for CAAP and non-CAAP; Jewish *vs*. non-Jewish for IPD). The covariates are a linear trend for time, 12-month harmonic variables to capture unexplained seasonal variations, and the proportion of tests positive for each virus in that month for the corresponding ethnicity (combined across all three age groups). We use an identity link, rather than a log-link so that the effects of each virus would be additive. This model generally follows the structure used by Zheng et al..^36^ The magnitude of the effects of each virus and the seasonal and trends terms were allowed to vary by age and ethnicity. For age group a, ethnicity e, time point t, the model structure was

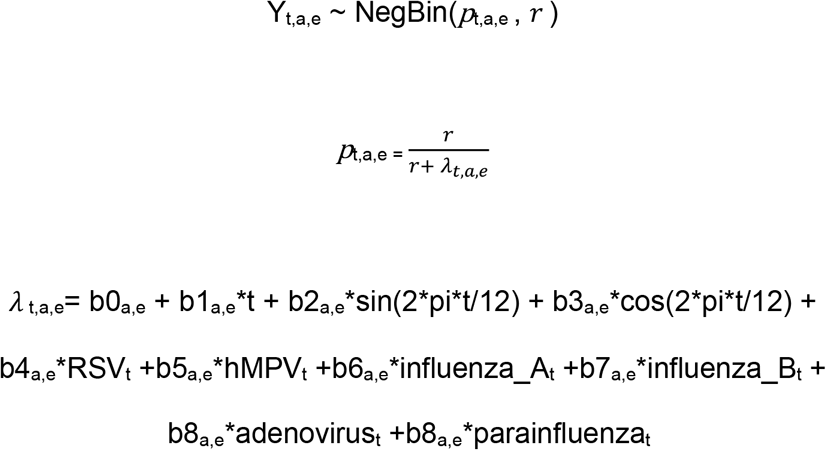

Y_t,a,e_ denotes the number of clinical cases for each time point *t*, age group *a*, and ethnicity *e. λ* _t,a,e_ represents the expected value of Y_t,a,e_ with a variance of *λ*_*ijk*_(1 + *λ*_*ijk*_/*r*) and an overdispersion parameter *r*. b0_a,e_ represents the intercept for the group, b1_a,e_ the time trend for the group, b3_a,e_ and b4_a,e_ capture the effect of baseline seasonality, and b4_a,e_ – b8_a,e_ represent the effect of each virus by group.

After fitting the models, the contribution of each virus was estimated from the fitted parameters. *λ*_*t,a,e*_ was recalculated by setting the bK_*a,e*_ term corresponding to the viruses to 0 and comparing these predicted values with the predicted values base on the fitted values of the parameters. Predicted values were summed by age group or overall to obtain aggregate estimates of the contribution of each virus. The viral time series were scaled to vary between 0 and 1. Each group-level effect was centred around a global effect, which allowed the coefficient to vary by group

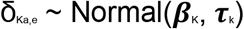

for the trend and harmonic coefficients (b1-b3),

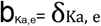

while for the intercept and for the effects of viruses, the effects were restricted to be non-negative:

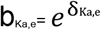

The model was fit in a Bayesian framework. The variance components (***τ***_k_) were given a uniform[0,100] prior on the standard deviation (e.g., sqrt(1/ ***τ***_k_^2^). ***β***_K_ were assigned minimally informative priors of Normal(0, 1). All model fitting was performed using rjags^37^ with three separate Markov chains and a burn-in period of 10,000 iterations (i.e., before convergence of the models) in each chain. Posterior inference was based on 30,000 samples (10,000 from each chain). The analysis code can be found at https://github.com/DanWeinberger/Israel_pneumo_covid.

Since RhV testing was initiated only in February 2019, we did not include this virus in the model. However, a separate regression analysis, adjusting for the same set of covariates as above, for the period of February 2019 through December 2021 showed no correlation of RhV to any of the studied clinical endpoints.

## Results

### I. Dynamics of Disease

During 2016 through December 2021, a total of 3,204, 26,695, 257, and 619 episodes of CAAP, non-CAAP LRI, pneumococcal bacteremic pneumonia, and non-pneumonia IPD, respectively, were recorded. The highest incidence occurred in children <1 year. (**Table 1, Supplementary Tables 1, 2**)

The dynamics of the four disease endpoints during the early pandemic period (through February 2021) were described in detail in a previous publication,^13^ and are included as part of **Figures 1 and 2; and Supplementary Tables 3 and 4**. In brief, broad declines in all four endpoints were observed. The largest decline in incidence was seen in CAAP and pneumococcal bacteremic pneumonia, while for non-CAAP-LRIs, although statistically significant, the reduction was less pronounced. For non-pneumomia IPD, the decline was not statistically significant.

**Figure 1:**
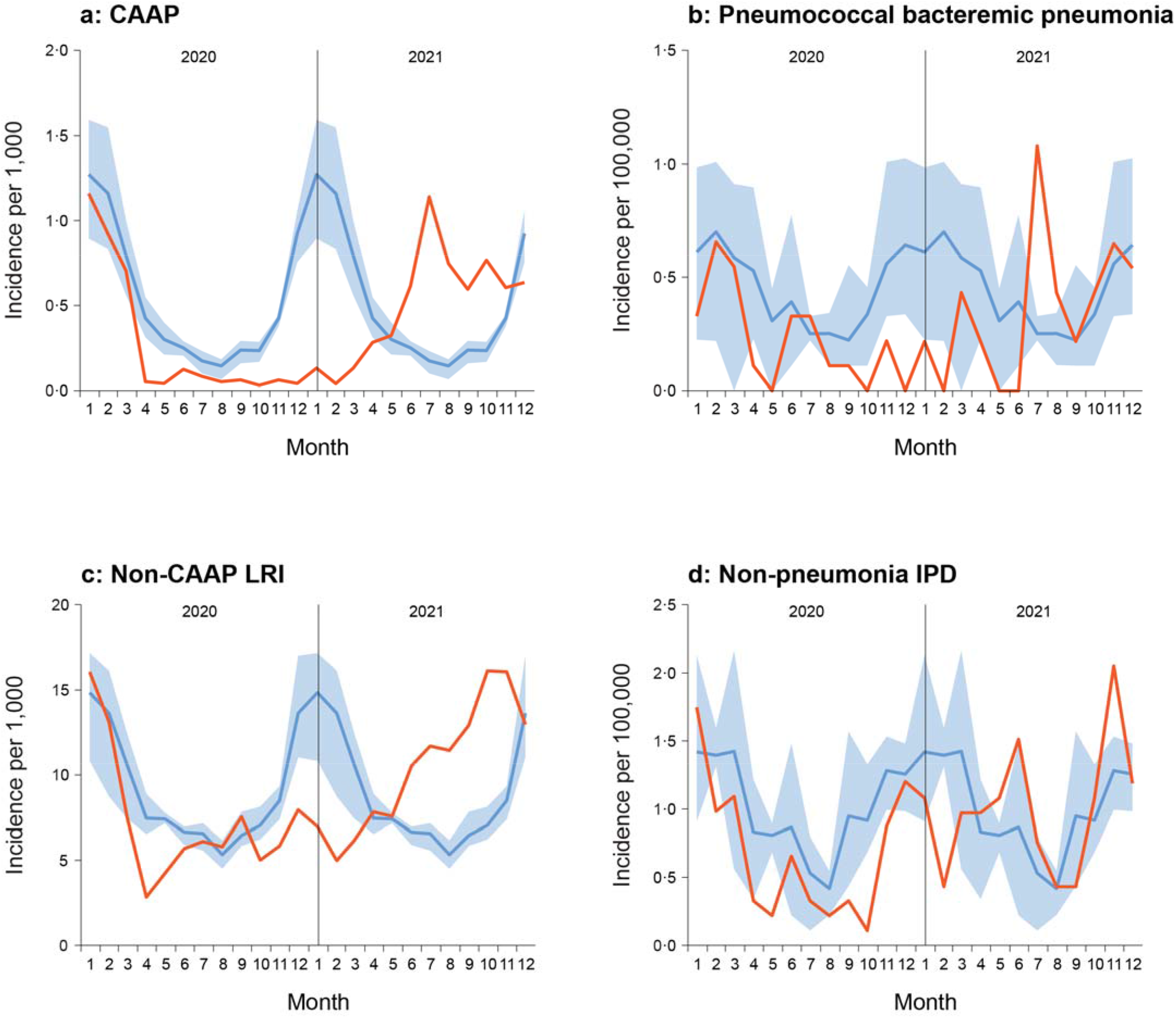
Monthly dynamics of community-acquired alveolar pneumonia (CAAP) **(a)**, pneumococcal bacteremic pneumonia **(b)**, non-community-acquired alveolar pneumonia lower respiratory infections (non-CAAP LRIs) **(c)**, and non-pneumonia invasive pneumococcal disease (non-pneumonia IPD) **(d)**. The red line represents incidences in 2020 and 2021, the blue line shows the mean incidence and the shaded blue area the minimum and maximum incidence during 2016 through 2019.

**Figure 2:**
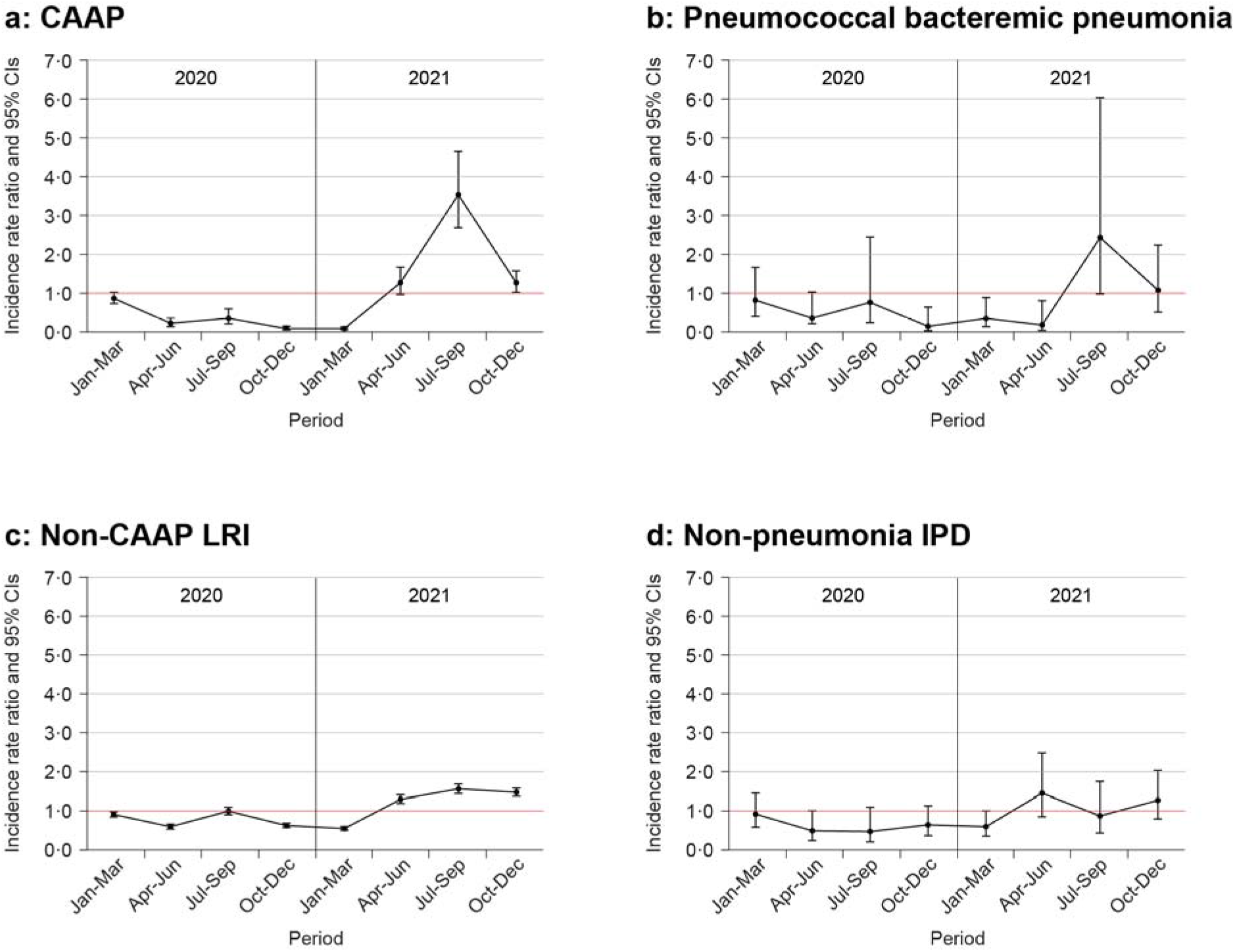
Incidence rates ratio (±95% CI) by quarters comparing incidence of 2020 and 2021 to the mean incidence of 2016 through 2019; community-acquired alveolar pneumonia (CAAP) **(a)**, pneumococcal bacteremic pneumonia **(b)**, non-community-acquired alveolar pneumonia lower respiratory infections (non-CAAP LRIs) **(c)**, and non-pneumonia invasive pneumococcal disease (non-pneumonia IPD) **(d)**. Black dots are quarterly IRRs; the vertical lines show 95% confidence intervals. The numerical values are presented in **Supplementary Table 3 and 4**.

Starting in spring 2021, an off-season, abrupt surge was observed with unexpectedly high rates, particularly for the 3 respiratory endpoints (CAAP, non-CAAP LRIs, and pneumococcal bacteremic pneumonia). In all endpoints, the off-season rates reached a magnitude similar to those observed during 2016-2019. (**Figures 1, 2, Supplementary Tables 3, 4**)

The dynamic patterns of all disease endpoints were similar when examined by age (<1, 1, and 2-4 years). (**Supplementary Figures 2-4**)

### II. Dynamics of pneumococcal nasopharyngeal carriage in children <3 year

A total of 7,921, 1,190, and 1,323 samples were obtained in 2016-2019, 2020, and 2021, respectively. (**Table 1**) During April and May 2020, we interrupted surveillance activities because of the severe lockdown but resumed from June 2020. (**Figure 3**)

**Figure 3:**
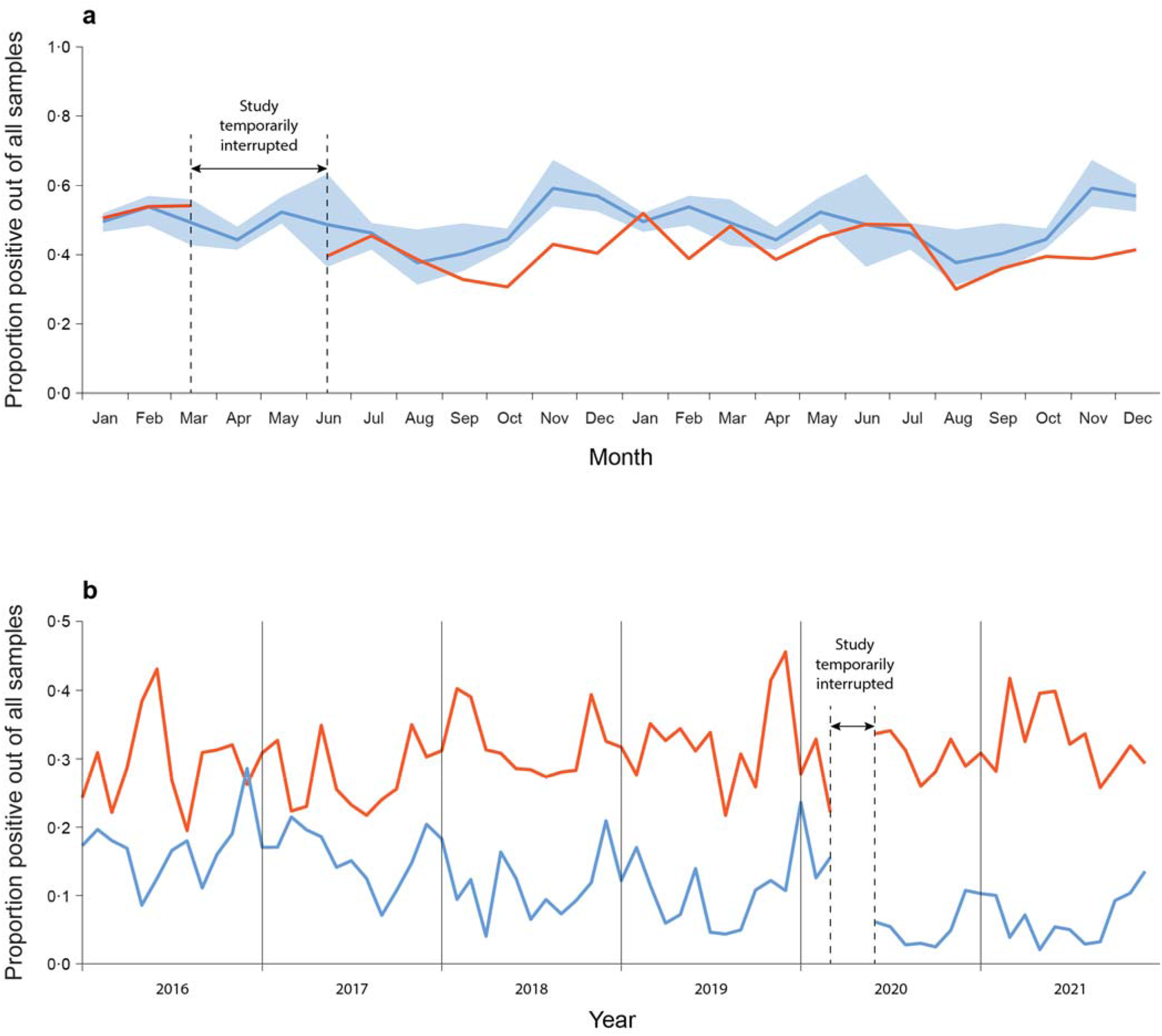
**(a)** Monthly dynamics of the proportion of all tested nasopharyngeal swabs that were positive for *S. pneumoniae* in children <3 years of age, in 2020 and 2021 compared to 2016 through 2019. The red line represents the proportion of 2020 and 2021, the blue line shows the mean proportion and the blue shaded area the minimum and maximum proportion during 2016 through 2019; **(b)** Dynamics of pneumococcus positive proportions of swab samples by semi quantitatively determined density: low density (1-2) represented by the blue line *vs*. high density (3-4), represented by the red line.

During 2016-2019, the mean monthly proportion of carriers ranged from 34·3% to 56·7% in 2016-2019, 29·0% to 52·1% in 2020, and 31·3% to 45·6% in 2021. Although from September 2020 to February 2021, carriage was somewhat lower than during 2016-2019 (*P*=0·067) (**Figure 3a, Supplementary Tables 5, 6**), no overall trend was observed, and changes in carriage prevalence did not correlate with changes in any of the disease endpoints. Furthermore, the semi-quantitative analysis showed a decline in lower-density colonization. but the prevalence of high-density colonization remained stable. (**Figure 3b, Supplementary Table 7**)

No significant changes in serotype distribution were found during the pandemic. (**Supplementary Table 8, Supplementary Figure 5**)

### III. Dynamics of viral activity in children <5 years

A total of 27,147 nasopharyngeal samples were obtained for virus detection, 17,555; 3,444; and 6,143 in 2016-2019, 2020, and 2021, respectively. (**Table 1**) Of these, 41·7%, 58·0%, and 63·4%, respectively, were positive for ≥1 virus: RSV 4,024, AdV 3,049, PIV 1,143, flu A 1,026, hMPV 883, and flu B 308. Testing for RhV was only initiated in February 2019 with 4,845 positive samples. (**Supplementary Table 9**) Co-detection of ≥2 viruses was common, especially starting in February 2019, when RhV testing was added. However, a co-detection of any PDA-virus with another PDA-virus was rare (1·3%). **(Supplementary Table 10)**

Strikingly, from April 2020 through October 2020, none of the PDA-viruses were detected. (**Figure 4, Supplementary Figure 6**) However, since November 2020, an off-season sequential re-emergence of the PDA-viruses was observed, each with a peak reaching an equal or higher magnitude compared to the pre-pandemic seasonal peaks: PIV (November 2020), hMPV (March 2021), RSV (May 2021), flu A (H_3_N_2_ only; September 2021). However, flu B was not detected from March 2020 through the end of the study. In contrast, although RhV and AdV had a short nadir during the time of the first lockdown (April-May 2020), they were frequently detected throughout the study. From March 2021, AdV had a higher-than-usual activity.

**Figure 4:**
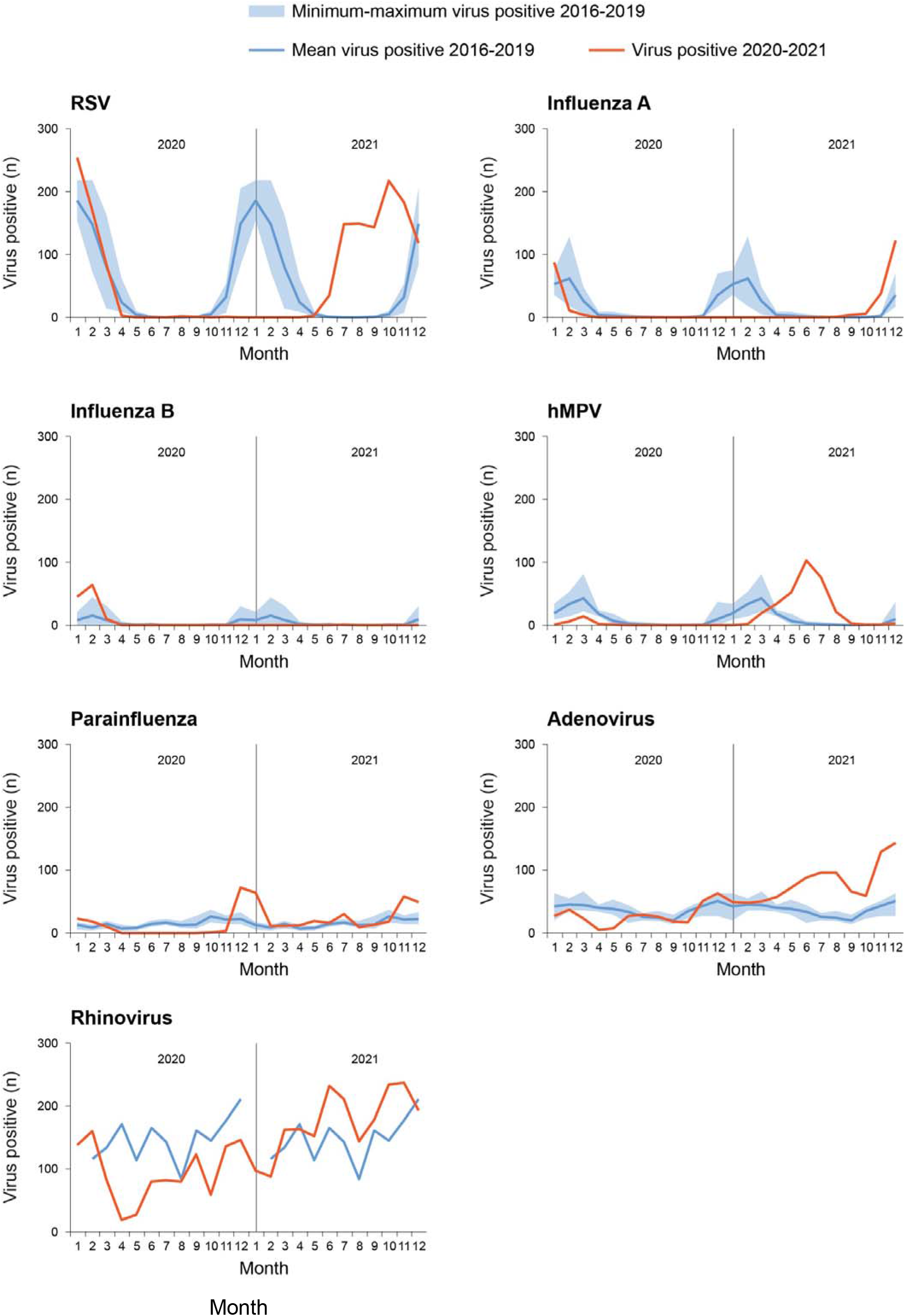
Dynamics of monthly numbers of all virus-positive nasal samples in children <5 years in southern Israel for specific viruses in 2020 and 2021 compared to 2016 through 2021. The red line represents the number of positive samples in 2020 and 2021, the blue line shows the mean number of positive samples and the shaded blue area the minimum and maximum number of positive samples during 2016 through 2019.

### IV. RSV and hMPV are major contributors to the burden of LRI and IPD

Due to the striking similarity of the dynamics of all 4 disease endpoints and the dynamics of the PDA-viruses, we sought to quantify the contribution of each virus. We first fit a model to CAAP data from the pre-pandemic period only. The main contributor was RSV (32%), followed by hMPV (5%), flu (4%), PIV (3%), and AdV (1%). (**Supplementary Table 11**) Projecting the model forward based on observed virus activity, this model accurately captured the surge in CAAP cases in 2021 among children <1 year but underestimated the number of CAAP cases among children aged 1 and 2-4 years (**Supplementary Figure 7**), suggesting that the model under-estimated the importance of the viruses in the older age groups. Since the activity of these viruses typically peaks around the same time in winter, it can be difficult to accurately attribute changes to specific viruses.

To disentangle the contribution of the viruses to the burden of pneumonia, we leveraged the different timing of the re-emergence of the different viruses in 2020-2021 and refit the models to the entire dataset. (**Figure 5, Table 2**) With these models, we estimated that 82% of CAAP cases could be attributed to the common respiratory viruses. The main contributor to this burden was RSV (49%), followed by hMPV (13%), PIV (11%), and flu (7%). AdV activity was not associated with variations in CAAP. The importance of the different viruses varied somewhat by age with smaller contributions overall from the viruses in older age groups. (**Figure 6, Table 2**) RSV was a particularly large contributor (62%) to CAAP among children <1 year.

**Figure 5:**
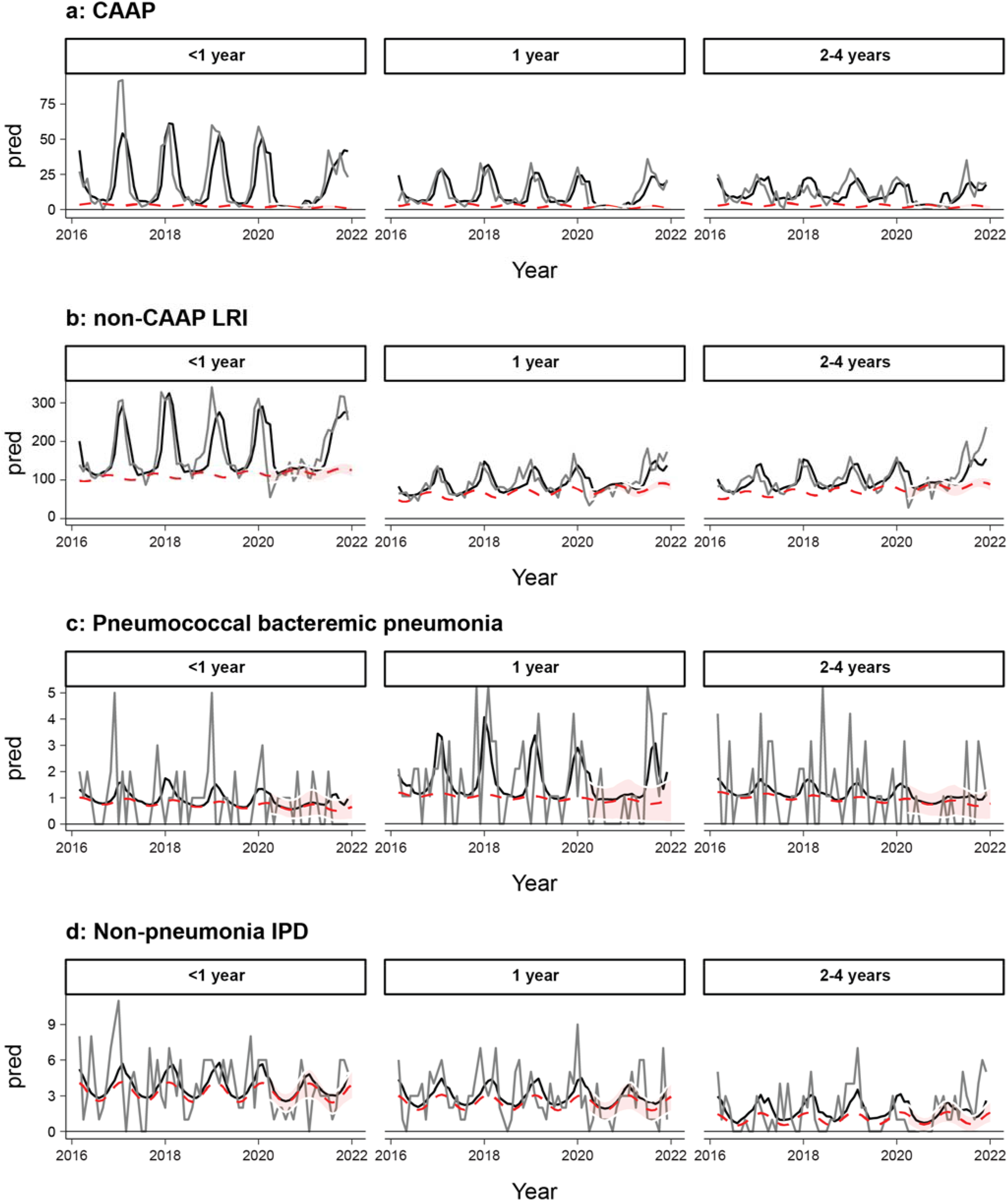
Observed (dark grey lines) and model-fitted values (black line) for the number of cases of community-acquired alveolar pneumonia (CAAP), non-community-acquired alveolar pneumonia lower respiratory infections (non-CAAP LRIs), pneumococcal bacteremic pneumonia, and non-pneumonia invasive pneumococcal disease (non-pneumonia IPD), based on a model fit to the entire data set. The red dashed line and shaded area represent the estimate for how many cases there would have been in the absence of viruses (±95% CI).

**Figure 6:**
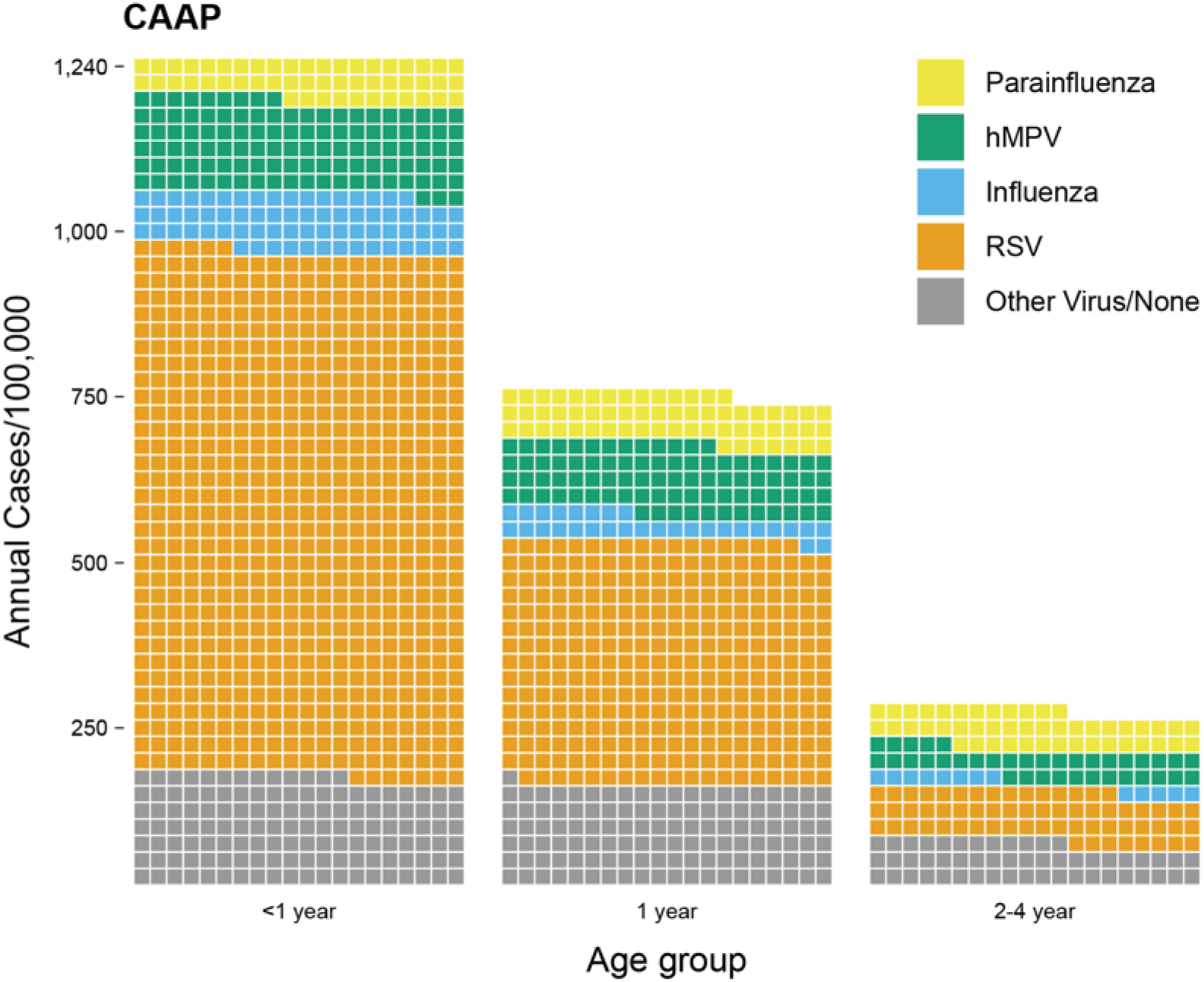
Estimated contribution of different viruses to the burden of community-acquired alveolar pneumonia (CAAP) among children <5 years of age in southern Israel. Parainfluenza – yellow; hMPV – green; influenza – blue; RSV – orange; other viruses/non – grey.

**Table 2:**
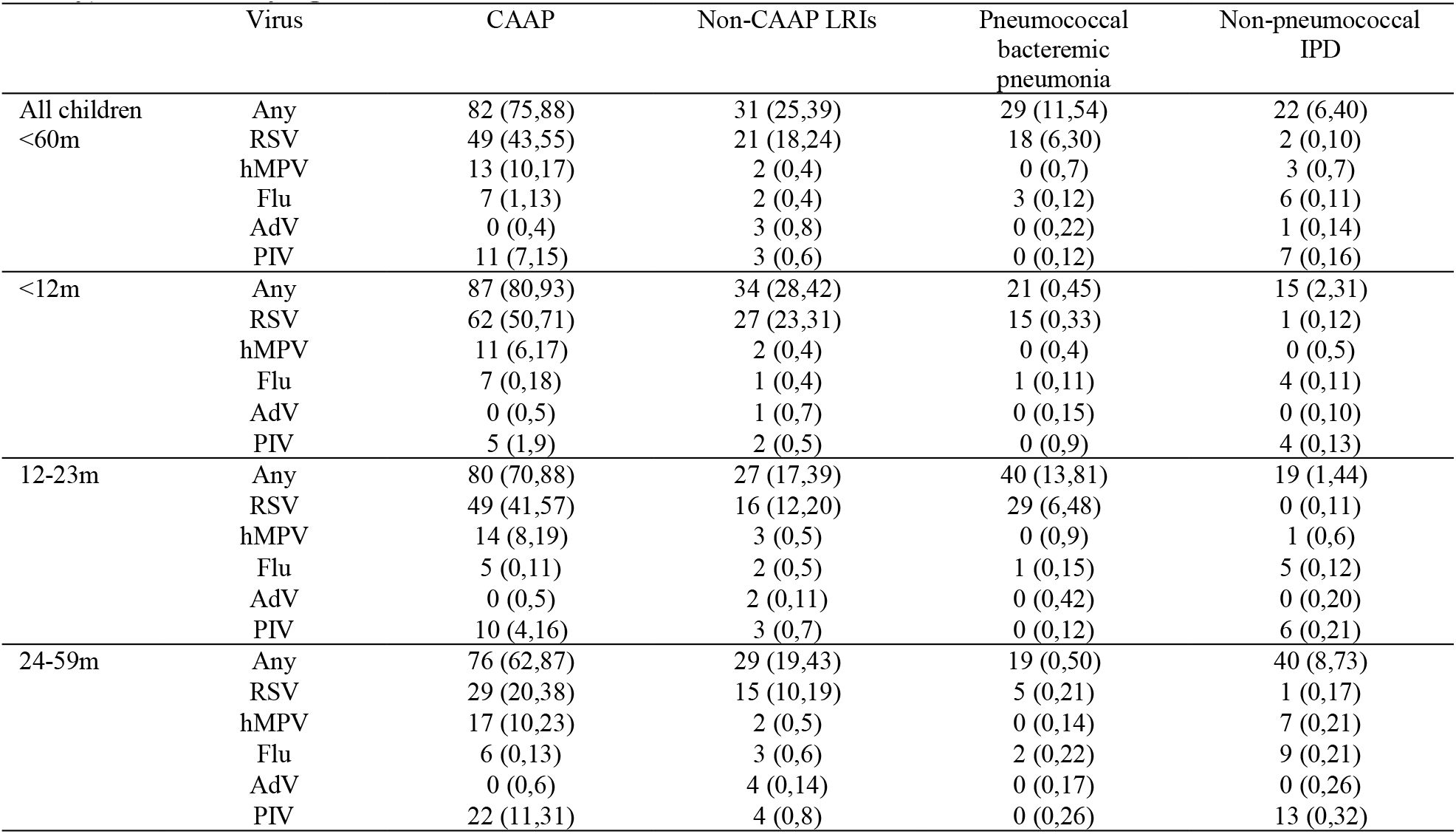
Percent of CAAP attributable to RSV, hMPV, Flu, AdV, PIV (adjusted for ethnicity), stratified by age.

|  | Virus | CAAP | Non-CAAP LRIs | Pneumococcal bacteremic pneumonia | Non-pneumococcal IPD |
| --- | --- | --- | --- | --- | --- |
| All children <60m | Any | 82 (75,88) | 31 (25,39) | 29 (11,54) | 22 (6,40) |
|  | RSV | 49 (43,55) | 21 (18,24) | 18 (6,30) | 2 (0,10) |
|  | hMPV | 13 (10,17) | 2 (0,4) | 0 (0,7) | 3 (0,7) |
|  | Flu | 7 (1,13) | 2 (0,4) | 3 (0,12) | 6 (0,11) |
|  | AdV | 0 (0,4) | 3 (0,8) | 0 (0,22) | 1 (0,14) |
|  | PIV | 11 (7,15) | 3 (0,6) | 0 (0,12) | 7 (0,16) |
| <12m | Any | 87 (80,93) | 34 (28,42) | 21 (0,45) | 15 (2,31) |
|  | RSV | 62 (50,71) | 27 (23,31) | 15 (0,33) | 1 (0,12) |
|  | hMPV | 11 (6,17) | 2 (0,4) | 0 (0,4) | 0 (0,5) |
|  | Flu | 7 (0,18) | 1 (0,4) | 1 (0,11) | 4 (0,11) |
|  | AdV | 0 (0,5) | 1 (0,7) | 0 (0,15) | 0 (0,10) |
|  | PIV | 5 (1,9) | 2 (0,5) | 0 (0,9) | 4 (0,13) |
| 12-23m | Any | 80 (70,88) | 27 (17,39) | 40 (13,81) | 19 (1,44) |
|  | RSV | 49 (41,57) | 16 (12,20) | 29 (6,48) | 0 (0,11) |
|  | hMPV | 14 (8,19) | 3 (0,5) | 0 (0,9) | 1 (0,6) |
|  | Flu | 5 (0,11) | 2 (0,5) | 1 (0,15) | 5 (0,12) |
|  | AdV | 0 (0,5) | 2 (0,11) | 0 (0,42) | 0 (0,20) |
|  | PIV | 10 (4,16) | 3 (0,7) | 0 (0,12) | 6 (0,21) |
| 24-59m | Any | 76 (62,87) | 29 (19,43) | 19 (0,50) | 40 (8,73) |
|  | RSV | 29 (20,38) | 15 (10,19) | 5 (0,21) | 1 (0,17) |
|  | hMPV | 17 (10,23) | 2 (0,5) | 0 (0,14) | 7 (0,21) |
|  | Flu | 6 (0,13) | 3 (0,6) | 2 (0,22) | 9 (0,21) |
|  | AdV | 0 (0,6) | 4 (0,14) | 0 (0,17) | 0 (0,26) |
|  | PIV | 22 (11,31) | 4 (0,8) | 0 (0,26) | 13 (0,32) |

The viruses also made important contributions to the other endpoints with 31% of non-CAAP cases, 29% of IPD pneumonia and 22% of non-pneumonia IPD associated with viruses. (**Table 2, Supplementary Figure 9**) For both non-CAAP and IPD pneumonia, RSV was the most important contributor, 21% and 18%, respectively. For non-pneumonia IPD, none of the PDA-viruses, individually, contributed significantly.

AdV did not contribute to any of the endpoints at any age.

## Discussion

The COVID-19 pandemic has temporarily but strikingly modified the epidemiology of both PDA-virus activity and the studied clinical endpoints in which pneumococcus plays an important causative role. These clinical endpoints declined sharply in 2020 and returned in 2021, together with the re-emergence of the PDA-viruses. These dramatic changes occurred in the presence of virtually unchanged nasopharyngeal carriage of pneumococcus, strongly suggesting its continuous circulation in the community. While PDA-virus activity was associated with the studied disease endpoints, that of RhV and AdV was not. A high proportion of the burden of CAAP and non-CAAP LRIs was associated with the activity of PDA-viruses, especially during the first year of life. We also showed that the modified dynamics of both disease and virus activity during the COVID-19 pandemic helped to improve estimates of the PDA-virus contribution to the studied disease endpoints. Finally, we showed that RSV had the highest attributable contribution to all studied endpoints, except for non-pneumonia IPD.

Multiple sites have described the abrupt reduction of pneumococcal diseases (mostly IPD) and all-cause respiratory diseases shortly following the start of the COVID-19 pandemic. The initial speculation was that the widely undertaken non-pharmaceutical interventions largely resulted in a reduction of pneumococcal transmission.^17, 18^ However, continuous active surveillance of pneumococcal carriage in young children showed normal or close to normal carriage rates, and no association with IPD or pneumococcal pneumonia could be demonstrated.^8, 13, 19, 22^ A recent report from Vietnam, while confirming no association of overall nasopharyngeal carriage during NPI, noted a marked decrease of the density of pneumococcus in carriage.^38^ In contrast, we found no reduction of carriage density, although measured by semi-quantitative methods. In addition, no change in serotype distribution was found, similar to findings from France.^12^ Therefore, it is unlikely that changes in carriage played an important role in the drastic disease incidence dynamics during COVID-19. No previous data on the carriage dynamics during the resurgence of pneumococcus-associated diseases have been reported.

It is not clear why the PDA-viruses largely disappeared during the initial phase of the pandemic, while other viruses and pneumococci continued to circulate in the presence of the various mitigation measures. It is possible that pneumococcus, AdV and RhV rely on different age groups or routes for transmission, they could be more transmissible, or additional mechanism such as viral-viral interactions could play a role.

During the study period, the dynamic patterns of the 4 studied disease endpoints were similar but not identical. The most affected rates were those of CAAP, with similar dynamics seen for pneumococcal bacteremic pneumonia. However, the smaller number of endpoint events of the latter resulted in a higher uncertainty range. Non-CAAP LRIs were also reduced during the first year of the pandemic, but the reduction was only 50% compared to that of CAAP. The non-pneumonia IPD incidence reductions, although exhibiting similar trends, did not reach significance in any of the time points, when compared to those in 2016-2019, consistent with the absence of demonstrable association with the PDA-viruses.

The contribution to the various disease endpoints differed between individual viruses, and between age groups. By far, the most important contributor to both CAAP, non-CAAP LRIs, and pneumococcal bacteremic pneumonia was RSV. In children <2 years, over half of the cases were attributable to RSV. Its contribution to non-CAAP LRIs was only around half compared to that in CAAP. This was somewhat unexpected, due to the common association with bronchiolitis. However, this difference suggests a potentially significant pathogenic role of RSV as a coinfecting agent with bacterial pathogens causing CAAP.

The extent of pneumococcal involvement in CAAP and non-CAAP-LRIs is still not fully clarified. This cannot be directly answered since detection of pneumococcus in the respiratory tract does not necessarily imply its causative role in CAAP. However, the profound impact of PCV on CAAP and even non-CAAP LRIs provides a powerful tool for inference on the likely causative role of *S. pneumoniae* in these disease endpoints.^14, 39^ Studies from Israel and elsewhere, have demonstrated a ∼50% reduction in CAAP in young children following PCV implementation,^14, 28^ thus strongly suggesting an extensive causative role of *S. pneumoniae* in CAAP during the pre-PCV era.

Our non-CAAP LRI cases were severe enough to warrant a chest radiograph, but per definition, those with CAAP were excluded. Studies have shown that such cases have a more diverse aetiology. Even so, a rate reduction of ≥25% was observed in young children post PCV implementation, including a 34% reduction in our region.^14^ Using a similar logic as per CAAP, it is plausible that vaccine-serotype pneumococci also played an important role in non-CAAP LRIs, although of a lesser magnitude than in CAAP. The similarity in dynamics between CAAP and non-CAAP LRIs is therefore not surprising.

Second to RSV, the estimated contribution of hMPV in CAAP is notable, contributing to approximately one in seven CAAP episodes in all age groups. The role of hMPV in CAAP has been well recognized.^11, 40-42^ However, in the current study we could compare its role in CAAP to that in other studied endpoints. Surprisingly, no significant contribution to any of the non-CAAP endpoints could be demonstrated. The specific mechanisms by which hMPV contributed mainly to CAAP are not elucidated. The attributable proportions of flu and PIV to both CAAP and non-CAAP, although significant, were modest, and the relative contribution of PIV increased with age. The lower relative contribution of flu to hospitalisation for LRIs compared to RSV and hMPV was described previously^41^ and is in contrast to reports from adults.^43^

We examined separately the dynamics of pneumococcal bacteremic pneumonia and non-pneumonia IPD. We have previously shown that after PCV7/PCV13 implementation in Israel, the dynamics of the two endpoints differed by specific pneumococcal serotype proportions and seasonality. The seasonality of bacteremic pneumonia was typical of respiratory viruses, while that of non-pneumonia IPD was not.^2^ In the current study we estimate that RSV played a significant role in pneumococcal bacteremic pneumonia, but its role in non-pneumonia IPD was negligible. This suggests some differences in the pathogenesis, further supporting previous observations.^44^ This might also explain, at least in part, the lack of significant reduction of the non-pneumonia IPD in the presence of significant reduction of the other three analysed endpoints.

The rates of these viruses were completely unrelated to those of the studied disease endpoints. Detection of RhV and AdV is common, both in healthy children and children with LRI, and their attributable role to LRI is often questionable.^45-47^ Furthermore, the co-detection of RhV and AdV with other viruses is common, and the co-presence with any of the PDA-viruses usually bears the clinical presentation typical of the specific coinfecting PDA-virus. In contrast, we found that co-detection of ≥2 PDA-viruses was rare (1·3%).

This study has a number of important strengths. We leverage unique and high-quality datasets that cover a long time period including pre-pandemic years. Together with the natural disruptions caused by the pandemic, these data provide a comprehensive understanding of the contributions of respiratory viruses to the aetiology of pneumococcal disease and other related respiratory diseases. These data are integrated using regressions models to estimate the contribution of each virus to the observed variations and estimates from models fit exclusively to the pre-pandemic period and those fit to the entire study period providing a range of estimates for the contribution of each virus.

The analyses also have important limitations. We did not account for the possible effects of coinfections or interactions between viruses. The data on viral activity are also limited by being conducted in the hospital setting. However, we expect that viral activity in the hospital likely reflects viral activity in the community. Finally, the causal relationship of the PDA-viruses with the studied endpoints could be confounded by a third causative factor. However, given the high degree of association, the multiple endpoints studied, and the results of the model, this possibility seems unlikely.

In conclusion, this study leverages the unique disruptions of pathogen circulation that followed COVID-19 mitigation measures to gain a deeper understanding of the role of respiratory viruses in driving the burden of pneumococcal disease and related conditions. These analyses suggest that several of the respiratory viruses, particularly RSV and hMPV, play a critical role in pathogenesis in children. This suggests that interventions that target these viruses could have secondary effects on the burden of disease typically attributed to bacteria.

## Data Availability

All data produced in the present study are available upon reasonable request to the authors

## Contributors

RD was responsible for concept, design, planning of the study; acquisition, analysis, and interpretation of the data; drafting and critically revising the manuscript. BAVDB contributed to the concept of the study, analysis, and interpretation of the data, as well as reviewing the manuscript for important intellectual content. SB-S was involved in the planning of the study and critically reviewed the manuscript for important intellectual content. DG reviewed the manuscript for important intellectual content.

YS-A contributed to the acquisition of the data and reviewed the manuscript for important intellectual content. DMW participated in the design of the study, analysis and interpretation of the data (modelling component), and reviewed the manuscript for important intellectual content. DD contributed to the design of the study, analysis, and interpretation of the data, as well as reviewing the manuscript for important intellectual content.

## Data sharing statement

All relevant data will be shared by the lead contact upon request.

