## Supplementary material for "The COVID-19 Pandemic as an Opportunity for Unravelling the Causative Association between Respiratory Viruses and Pneumococcus-Associated Disease in Young Children: A Prospective Study": Supplemenatry Tables

**Supplementary Table 1: Incidence of CAAP, non-LRI, pneumococcal bacteremic pneumonia, and non-pneumococcal IPD in children <5 years per month per study year**

| | CAAP incidence per 1,000 | | | | NA-LRI incidence per 1,000 | | | | Pneumococcal bacteremic pneumonia incidence per 100,000 | | | | Non-pneumonia IPD incidence per 100,000 | | | |
|---|---|---|---|---|---|---|---|---|---|---|---|---|---|---|---|---|
| | <1 year | 1 year | 2-4 years | <5 years | <1 year | 1 year | 2-4 years | <5 years | <1 year | 1 year | 2-4 years | <5 years | <1 year | 1 year | 2-4 years | <5 years |
| **2016** | | | | | | | | | | | | | | | | |
| Jan | 3·48 | 1·45 | 0·36 | 1·27 | 10·83 | 5·41 | 2·20 | 4·74 | 2·23 | 1·12 | 0·00 | 0·68 | 2·23 | 1·68 | 0·19 | 0·91 |
| Feb | 1·77 | 1·51 | 0·26 | 0·85 | 8·57 | 3·46 | 1·88 | 3·66 | 1·11 | 1·68 | 0·58 | 0·91 | 2·79 | 2·24 | 0·96 | 1·59 |
| Mar | 1·45 | 0·33 | 0·50 | 0·67 | 7·45 | 3·63 | 1·67 | 3·33 | 1·11 | 1·12 | 0·77 | 0·91 | 4·46 | 3·36 | 0·96 | 2·16 |
| Apr | 0·86 | 0·73 | 0·38 | 0·56 | 6·54 | 3·68 | 1·76 | 3·19 | 0·56 | 0·56 | 0·00 | 0·23 | 0·56 | 0·56 | 0·19 | 0·34 |
| May | 1·18 | 0·22 | 0·10 | 0·36 | 7·77 | 3·79 | 1·41 | 3·29 | 1·11 | 0·56 | 0·00 | 0·34 | 1·67 | 0·00 | 0·58 | 0·68 |
| Jun | 0·32 | 0·45 | 0·18 | 0·27 | 5·90 | 2·62 | 1·47 | 2·67 | 0·56 | 0·56 | 0·58 | 0·57 | 4·46 | 1·68 | 0·38 | 1·48 |
| Jul | 0·21 | 0·33 | 0·12 | 0·19 | 5·57 | 3·35 | 1·27 | 2·64 | 0·00 | 1·12 | 0·00 | 0·23 | 2·23 | 1·68 | 0·00 | 0·80 |
| Aug | 0·38 | 0·22 | 0·10 | 0·19 | 6·16 | 2·57 | 1·59 | 2·79 | 0·00 | 1·12 | 0·19 | 0·34 | 0·56 | 1·68 | 0·00 | 0·46 |
| Sep | 0·00 | 0·33 | 0·16 | 0·16 | 5·84 | 4·18 | 2·00 | 3·29 | 0·00 | 0·00 | 0·19 | 0·11 | 1·11 | 2·24 | 0·00 | 0·68 |
| Oct | 0·27 | 0·28 | 0·30 | 0·29 | 6·75 | 5·13 | 2·12 | 3·75 | 0·56 | 1·12 | 0·19 | 0·46 | 2·23 | 0·56 | 0·19 | 0·68 |
| Nov | 0·86 | 0·61 | 0·24 | 0·45 | 7·40 | 4·30 | 2·24 | 3·78 | 1·11 | 0·56 | 0·19 | 0·46 | 3·90 | 2·80 | 0·00 | 1·37 |
| Dec | 2·41 | 1·06 | 0·34 | 0·94 | 11·04 | 6·69 | 2·06 | 4·97 | 2·79 | 1·12 | 0·38 | 1·02 | 5·01 | 2·24 | 0·00 | 1·48 |
| **2017** | | | | | | | | | | | | | | | | |
| Jan | 4·72 | 1·39 | 0·49 | 1·60 | 15·72 | 7·29 | 2·53 | 6·38 | 0·00 | 1·10 | 0·00 | 0·22 | 6·05 | 2·76 | 0·57 | 2·13 |
| Feb | 4·77 | 1·55 | 0·35 | 1·56 | 15·93 | 5·04 | 1·88 | 5·59 | 1·10 | 1·66 | 0·76 | 1·01 | 2·20 | 3·31 | 0·38 | 1·35 |
| Mar | 2·39 | 1·02 | 0·33 | 0·92 | 10·74 | 4·98 | 1·88 | 4·45 | 0·00 | 0·00 | 0·00 | 0·00 | 0·00 | 2·21 | 0·19 | 0·56 |
| Apr | 0·62 | 0·38 | 0·20 | 0·33 | 7·21 | 3·43 | 1·21 | 2·98 | 1·10 | 1·66 | 0·57 | 0·90 | 2·20 | 1·10 | 0·00 | 0·67 |
| May | 0·42 | 0·43 | 0·06 | 0·21 | 7·37 | 3·32 | 1·55 | 3·18 | 0·55 | 0·00 | 0·57 | 0·45 | 2·75 | 1·10 | 0·19 | 0·09 |
| Jun | 0·31 | 0·38 | 0·25 | 0·29 | 6·80 | 3·27 | 1·78 | 3·18 | 0·00 | 0·00 | 0·19 | 0·11 | 1·65 | 1·66 | 0·00 | 0·67 |
| Jul | 0·10 | 0·21 | 0·06 | 0·10 | 6·59 | 3·59 | 1·29 | 2·92 | 0·00 | 1·10 | 0·00 | 0·22 | 0·00 | 0·55 | 0·76 | 0·56 |
| Aug | 0·16 | 0·11 | 0·02 | 0·07 | 4·51 | 2·89 | 1·23 | 2·29 | 0·00 | 0·55 | 0·00 | 0·11 | 0·00 | 0·55 | 0·19 | 0·22 |
| Sep | 0·26 | 0·27 | 0·24 | 0·25 | 5·97 | 5·14 | 1·53 | 3·25 | 0·00 | 0·55 | 0·00 | 0·11 | 3·85 | 1·66 | 0·76 | 1·57 |
| Oct | 0·42 | 0·38 | 0·14 | 0·25 | 6·23 | 4·61 | 1·68 | 3·28 | 0·55 | 0·55 | 0·19 | 0·34 | 1·65 | 1·66 | 0·19 | 0·78 |
| Nov | 0·62 | 0·75 | 0·16 | 0·38 | 9·34 | 5·90 | 1·78 | 4·28 | 1·65 | 2·76 | 0·19 | 1·01 | 3·30 | 2·76 | 0·00 | 1·23 |
| Dec | 2·33 | 1·77 | 0·33 | 1·07 | 17·02 | 7·77 | 3·00 | 7·03 | 0·55 | 0·00 | 0·38 | 0·34 | 1·65 | 3·87 | 0·00 | 1·12 |
| **2018** | | | | | | | | | | | | | | | | |
| Jan | 2·41 | 1·25 | 0·23 | 0·91 | 15·67 | 6·74 | 2·58 | 6·28 | 0·00 | 1·64 | 0·37 | 0·55 | 1·09 | 2·18 | 0·56 | 0·99 |
| Feb | 3·07 | 1·50 | 0·33 | 1·17 | 16·13 | 6·02 | 2·61 | 6·24 | 0·55 | 2·73 | 0·00 | 0·66 | 3·82 | 1·64 | 0·37 | 1·33 |
| Mar | 1·28 | 0·83 | 0·21 | 0·57 | 12·44 | 4·67 | 2·04 | 4·83 | 0·55 | 1·64 | 0·56 | 0·77 | 2·73 | 2·18 | 0·93 | 1·55 |
| Apr | 0·77 | 0·31 | 0·15 | 0·32 | 7·22 | 3·89 | 1·61 | 3·30 | 1·09 | 1·64 | 0·00 | 0·55 | 1·64 | 3·82 | 0·19 | 1·22 |
| May | 0·61 | 0·57 | 0·27 | 0·41 | 7·37 | 4·25 | 1·81 | 3·52 | 0·00 | 0·00 | 0·00 | 0·00 | 3·27 | 1·09 | 0·00 | 0·88 |
| Jun | 0·31 | 0·31 | 0·13 | 0·21 | 6·91 | 4·20 | 1·67 | 3·34 | 1·09 | 0·00 | 0·93 | 0·77 | 0·55 | 0·55 | 0·00 | 0·22 |
| Jul | 0·46 | 0·21 | 0·06 | 0·18 | 7·17 | 3·79 | 1·60 | 3·26 | 0·00 | 0·55 | 0·19 | 0·22 | 0·55 | 0·00 | 0·00 | 0·11 |
| Aug | 0·15 | 0·26 | 0·13 | 0·17 | 5·33 | 4·15 | 1·35 | 2·80 | 0·55 | 0·55 | 0·19 | 0·33 | 1·09 | 0·55 | 0·19 | 0·44 |
| Sep | 0·36 | 0·42 | 0·23 | 0·30 | 7·83 | 4·88 | 1·86 | 3·79 | 0·55 | 1·09 | 0·37 | 0·55 | 2·18 | 2·73 | 0·19 | 1·11 |
| Oct | 0·31 | 0·21 | 0·21 | 0·23 | 8·14 | 6·90 | 2·06 | 4·39 | 0·55 | 1·09 | 0·19 | 0·44 | 1·64 | 3·27 | 0·56 | 1·33 |
| Nov | 0·72 | 0·42 | 0·33 | 0·43 | 8·81 | 5·34 | 1·96 | 4·15 | 0·55 | 1·64 | 0·00 | 0·44 | 3·27 | 1·09 | 0·19 | 0·99 |
| Dec | 1·59 | 0·93 | 0·40 | 0·77 | 12·03 | 6·59 | 2·31 | 5·31 | 1·09 | 0·55 | 0·19 | 0·44 | 3·27 | 1·64 | 0·74 | 1·44 |
| **2019** | | | | | | | | | | | | | | | | |
| Jan | 3·03 | 1·69 | 0·54 | 1·31 | 17·17 | 7·83 | 2·97 | 7·00 | 2·74 | 0·00 | 0·73 | 0·98 | 3·29 | 2·70 | 0·73 | 1·64 |
| Feb | 2·83 | 1·02 | 0·46 | 1·08 | 13·74 | 4·97 | 2·39 | 5·34 | 0·00 | 1·08 | 0·00 | 0·22 | 2·74 | 1·62 | 0·73 | 1·31 |
| Mar | 2·78 | 1·13 | 0·33 | 1·02 | 11·77 | 5·38 | 2·15 | 4·87 | 0·00 | 2·16 | 0·37 | 0·66 | 2·19 | 1·08 | 1·28 | 1·42 |
| Apr | 1·21 | 0·46 | 0·24 | 0·49 | 8·89 | 4·35 | 1·69 | 3·77 | 0·55 | 1·08 | 0·18 | 0·44 | 3·29 | 1·08 | 0·37 | 1·09 |
| May | 0·30 | 0·26 | 0·19 | 0·23 | 7·17 | 3·64 | 1·37 | 3·08 | 0·55 | 0·00 | 0·55 | 0·44 | 2·19 | 0·54 | 0·37 | 0·77 |
| Jun | 0·25 | 0·46 | 0·13 | 0·23 | 6·97 | 4·10 | 2·11 | 3·56 | 0·00 | 0·54 | 0·00 | 0·11 | 3·29 | 2·16 | 0·00 | 1·09 |
| Jul | 0·25 | 0·41 | 0·17 | 0·24 | 6·92 | 3·79 | 1·65 | 3·22 | 1·10 | 0·54 | 0·00 | 0·33 | 2·19 | 1·08 | 0·00 | 0·66 |
| Aug | 0·10 | 0·20 | 0·15 | 0·15 | 5·30 | 2·77 | 1·22 | 2·41 | 0·00 | 0·54 | 0·18 | 0·22 | 1·10 | 1·62 | 0·00 | 0·55 |
| Sep | 0·15 | 0·15 | 0·32 | 0·25 | 6·16 | 5·02 | 1·37 | 3·15 | 0·00 | 0·00 | 0·18 | 0·11 | 1·10 | 1·08 | 0·00 | 0·44 |
| Oct | 0·30 | 0·26 | 0·09 | 0·17 | 7·07 | 5·79 | 1·95 | 3·84 | 0·00 | 0·54 | 0·00 | 0·11 | 2·74 | 1·08 | 0·18 | 0·88 |
| Nov | 0·76 | 0·67 | 0·24 | 0·44 | 8·43 | 4·97 | 2·08 | 4·03 | 0·00 | 1·08 | 0·18 | 0·33 | 4·39 | 2·70 | 0·18 | 1·53 |
| Dec | 2·32 | 1·08 | 0·37 | 0·93 | 14·44 | 8·60 | 2·47 | 6·29 | 0·55 | 2·16 | 0·37 | 0·77 | 1·10 | 2·16 | 0·55 | 0·98 |
| **2020** | | | | | | | | | | | | | | | | |
| Jan | 3·05 | 1·51 | 0·41 | 1·18 | 16·06 | 6·31 | 2·85 | 6·26 | 1·12 | 0·54 | 0·00 | 0·33 | 3·35 | 4·90 | 0·18 | 1·75 |
| Feb | 2·63 | 0·91 | 0·36 | 0·94 | 13·12 | 5·35 | 2·11 | 5·03 | 1·68 | 1·63 | 0·00 | 0·66 | 3·35 | 1·09 | 0·18 | 0·98 |
| Mar | 1·65 | 0·91 | 0·32 | 0·72 | 7·54 | 2·88 | 1·70 | 3·14 | 0·00 | 1·09 | 0·54 | 0·55 | 3·35 | 1·63 | 0·18 | 1·09 |
| Apr | 0·15 | 0·00 | 0·04 | 0·05 | 2·84 | 1·72 | 0·50 | 1·23 | 0·56 | 0·00 | 0·00 | 0·11 | 1·68 | 0·00 | 0·00 | 0·33 |
| May | 0·05 | 0·05 | 0·04 | 0·04 | 4·24 | 2·32 | 0·91 | 1·88 | 0·00 | 0·00 | 0·00 | 0·00 | 0·56 | 0·54 | 0·00 | 0·22 |
| Jun | 0·21 | 0·10 | 0·11 | 0·13 | 5·68 | 3·43 | 1·52 | 2·77 | 0·56 | 0·54 | 0·18 | 0·33 | 1·12 | 2·18 | 0·00 | 0·66 |

| | | | | | | | | | | | | | | | | |
|---|---|---|---|---|---|---|---|---|---|---|---|---|---|---|---|---|
| Jul | 0·21 | 0·10 | 0·04 | 0·08 | 6·10 | 3·43 | 1·31 | 2·73 | 0·56 | 1·09 | 0·00 | 0·33 | 1·12 | 0·54 | 0·00 | 0·33 |
| Aug | 0·15 | 0·05 | 0·02 | 0·05 | 5·79 | 3·79 | 1·36 | 2·77 | 0·56 | 0·00 | 0·00 | 0·11 | 0·56 | 0·54 | 0·00 | 0·22 |
| Sep | 0·05 | 0·2 | 0·02 | 0·06 | 7·54 | 4·09 | 1·92 | 3·51 | 0·56 | 0·00 | 0·00 | 0·11 | 1·12 | 0·54 | 0·00 | 0·33 |
| Oct | 0·05 | 0·05 | 0·02 | 0·03 | 5·01 | 2·88 | 0·91 | 2·16 | 0·00 | 0·00 | 0·00 | 0·00 | 0·56 | 0·00 | 0·00 | 0·11 |
| Nov | 0·05 | 0·10 | 0·05 | 0·06 | 5·84 | 4·24 | 1·59 | 3·01 | 1·12 | 0·00 | 0·00 | 0·22 | 1·68 | 1·09 | 0·54 | 0·87 |
| Dec | 0·05 | 0·05 | 0·04 | 0·04 | 7·95 | 4·34 | 1·66 | 3·50 | 0·00 | 0·00 | 0·00 | 0·00 | 3·35 | 1·63 | 0·36 | 1·20 |
| **2021** | | | | | | | | | | | | | | | | |
| Jan | 0·20 | 0·15 | 0·07 | 0·11 | 6·97 | 4·24 | 1·43 | 3·12 | 0·00 | 0·54 | 0·18 | 0·22 | 2·23 | 1·62 | 0·53 | 1·08 |
| Feb | 0·10 | 0·05 | 0·02 | 0·04 | 4·98 | 3·41 | 1·25 | 2·45 | 0·00 | 0·00 | 0·00 | 0·00 | 0·00 | 1·62 | 0·18 | 0·43 |
| Mar | 0·20 | 0·21 | 0·09 | 0·13 | 6·15 | 4·55 | 1·84 | 3·26 | 1·12 | 0·54 | 0·18 | 0·43 | 1·67 | 2·69 | 0·18 | 0·97 |
| Apr | 0·46 | 0·57 | 0·14 | 0·29 | 7·83 | 4·96 | 1·79 | 3·66 | 0·56 | 0·54 | 0·00 | 0·22 | 2·23 | 1·08 | 0·53 | 0·97 |
| May | 0·41 | 0·72 | 0·17 | 0·33 | 7·58 | 4·70 | 2·07 | 3·72 | 0·00 | 0·00 | 0·00 | 0·00 | 1·67 | 3·23 | 0·18 | 1·08 |
| Jun | 0·97 | 1·03 | 0·38 | 0·63 | 10·48 | 7·9 | 2·98 | 5·49 | 0 | 0 | 0 | 0 | 1·67 | 3·23 | 0·89 | 1·51 |
| Jul | 2·14 | 1·86 | 0·61 | 1·17 | 11·7 | 9·4 | 3·48 | 6·34 | 1·12 | 2·69 | 0·53 | 1·08 | 1·67 | 0·54 | 0·53 | 0·76 |
| Aug | 1·63 | 1·39 | 0·26 | 0·77 | 11·44 | 6·46 | 2·35 | 5·03 | 0 | 2·16 | 0 | 0·43 | 0·56 | 0·54 | 0·36 | 0·43 |
| Sep | 1·27 | 1·29 | 0·14 | 0·60 | 12·92 | 6·61 | 2·91 | 5·69 | 0 | 1·08 | 0 | 0·22 | 1·12 | 0·54 | 0·18 | 0·43 |
| Oct | 2·03 | 0·88 | 0·33 | 0·79 | 16·12 | 8·68 | 3·06 | 6·85 | 0 | 0·54 | 0·53 | 0·43 | 3·35 | 0 | 0·71 | 1·08 |
| Nov | 1·42 | 0·72 | 0·31 | 0·62 | 16·07 | 7·65 | 3·48 | 6·88 | 0 | 2·16 | 0·36 | 0·65 | 3·35 | 3·77 | 1·07 | 2·05 |
| Dec | 1·17 | 1·08 | 0·33 | 0·65 | 12·97 | 8·88 | 4·12 | 6·88 | 0 | 2·16 | 0·18 | 0·54 | 2·79 | 0·54 | 0·89 | 1·19 |

**Supplementary Table 2: IPD episodes in children under 5 years of age, nationwide January 2016 through December 2021, per age group**

| | | <1 year of age | | | 1 year of age | | | 2-4 years of age | | | <5 years of age | | |
|---|---|---|---|---|---|---|---|---|---|---|---|---|---|
| | | Pneumonic bacteremia | Other IPD | Total IPD | Pneumonic bacteremia | Other IPD | Total IPD | Pneumonic bacteremia | Other IPD | Total IPD | Pneumonic bacteremia | Other IPD | Total IPD |
| **2016** | Jan | 4 | 4 | 8 | 2 | 3 | 5 | 0 | 1 | 1 | 6 | 8 | 14 |
| | Feb | 2 | 5 | 7 | 3 | 4 | 7 | 3 | 5 | 8 | 8 | 14 | 22 |
| | Mar | 2 | 8 | 10 | 2 | 6 | 8 | 4 | 5 | 9 | 8 | 19 | 27 |
| | Apr | 1 | 1 | 2 | 1 | 1 | 2 | 0 | 1 | 1 | 2 | 3 | 5 |
| | May | 2 | 3 | 5 | 1 | 0 | 1 | 0 | 3 | 3 | 3 | 6 | 9 |
| | Jun | 1 | 8 | 9 | 1 | 3 | 4 | 3 | 2 | 5 | 5 | 13 | 18 |
| | Jul | 0 | 4 | 4 | 2 | 3 | 5 | 0 | 0 | 0 | 2 | 7 | 9 |
| | Aug | 0 | 1 | 1 | 2 | 3 | 5 | 1 | 0 | 1 | 3 | 4 | 7 |
| | Sep | 0 | 2 | 2 | 0 | 4 | 4 | 1 | 0 | 1 | 1 | 6 | 7 |
| | Oct | 1 | 4 | 5 | 2 | 1 | 3 | 1 | 1 | 2 | 4 | 6 | 10 |
| | Nov | 2 | 7 | 9 | 1 | 5 | 6 | 1 | 0 | 1 | 4 | 12 | 16 |
| | Dec | 5 | 9 | 14 | 2 | 4 | 6 | 2 | 0 | 2 | 9 | 13 | 22 |
| **2017** | Jan | 0 | 11 | 11 | 2 | 5 | 7 | 0 | 3 | 3 | 2 | 19 | 21 |
| | Feb | 2 | 4 | 6 | 3 | 6 | 9 | 4 | 2 | 6 | 9 | 12 | 21 |
| | Mar | 0 | 0 | 0 | 0 | 4 | 4 | 0 | 1 | 1 | 0 | 5 | 5 |
| | Apr | 2 | 4 | 6 | 3 | 2 | 5 | 3 | 0 | 3 | 8 | 6 | 14 |
| | May | 1 | 5 | 6 | 0 | 2 | 2 | 3 | 1 | 4 | 4 | 8 | 12 |
| | Jun | 0 | 3 | 3 | 0 | 3 | 3 | 1 | 0 | 1 | 1 | 6 | 7 |
| | Jul | 0 | 0 | 0 | 2 | 1 | 3 | 0 | 4 | 4 | 2 | 5 | 7 |
| | Aug | 0 | 0 | 0 | 1 | 1 | 2 | 0 | 1 | 1 | 1 | 2 | 3 |
| | Sep | 0 | 7 | 7 | 1 | 3 | 4 | 0 | 4 | 4 | 1 | 14 | 15 |
| | Oct | 1 | 3 | 4 | 1 | 3 | 4 | 1 | 1 | 2 | 3 | 7 | 10 |
| | Nov | 3 | 6 | 9 | 5 | 5 | 10 | 1 | 0 | 1 | 9 | 11 | 20 |
| | Dec | 1 | 3 | 4 | 0 | 7 | 7 | 2 | 0 | 2 | 3 | 10 | 13 |
| **2018** | Jan | 0 | 2 | 2 | 3 | 4 | 7 | 2 | 3 | 5 | 5 | 9 | 14 |
| | Feb | 1 | 7 | 8 | 5 | 3 | 8 | 0 | 2 | 2 | 6 | 12 | 18 |
| | Mar | 1 | 5 | 6 | 3 | 4 | 7 | 3 | 5 | 8 | 7 | 14 | 21 |
| | Apr | 2 | 3 | 5 | 3 | 7 | 10 | 0 | 1 | 1 | 5 | 11 | 16 |
| | May | 0 | 6 | 6 | 0 | 2 | 2 | 0 | 0 | 0 | 0 | 8 | 8 |
| | Jun | 2 | 1 | 3 | 0 | 1 | 1 | 5 | 0 | 5 | 7 | 2 | 9 |
| | Jul | 0 | 1 | 1 | 1 | 0 | 1 | 1 | 0 | 1 | 2 | 1 | 3 |
| | Aug | 1 | 2 | 3 | 1 | 1 | 2 | 1 | 1 | 2 | 3 | 4 | 7 |
| | Sep | 1 | 4 | 5 | 2 | 5 | 7 | 2 | 1 | 3 | 5 | 10 | 15 |
| | Oct | 1 | 3 | 4 | 2 | 6 | 8 | 1 | 3 | 4 | 4 | 12 | 16 |
| | Nov | 1 | 6 | 7 | 3 | 2 | 5 | 0 | 1 | 1 | 4 | 9 | 13 |
| | Dec | 2 | 6 | 8 | 1 | 3 | 4 | 1 | 4 | 5 | 4 | 13 | 17 |
| **2019** | Jan | 5 | 6 | 11 | 0 | 5 | 5 | 4 | 4 | 8 | 9 | 15 | 24 |
| | Feb | 0 | 5 | 5 | 2 | 3 | 5 | 0 | 4 | 4 | 2 | 12 | 14 |
| | Mar | 0 | 4 | 4 | 4 | 2 | 6 | 2 | 7 | 9 | 6 | 13 | 19 |
| | Apr | 1 | 6 | 7 | 2 | 2 | 4 | 1 | 2 | 3 | 4 | 10 | 14 |
| | May | 1 | 4 | 5 | 0 | 1 | 1 | 3 | 2 | 5 | 4 | 7 | 11 |
| | Jun | 0 | 6 | 6 | 1 | 4 | 5 | 0 | 0 | 0 | 1 | 10 | 11 |
| | Jul | 2 | 4 | 6 | 1 | 2 | 3 | 0 | 0 | 0 | 3 | 6 | 9 |
| | Aug | 0 | 2 | 2 | 1 | 3 | 4 | 1 | 0 | 1 | 2 | 5 | 7 |
| | Sep | 0 | 2 | 2 | 0 | 2 | 2 | 1 | 0 | 1 | 1 | 4 | 5 |
| | Oct | 0 | 5 | 5 | 1 | 2 | 3 | 0 | 1 | 1 | 1 | 8 | 9 |

| | | | | | | | | | | | | | |
|---|---|---|---|---|---|---|---|---|---|---|---|---|---|
| | Nov | 0 | 8 | 8 | 2 | 5 | 7 | 1 | 1 | 2 | 3 | 14 | 17 |
| | Dec | 1 | 2 | 3 | 4 | 4 | 8 | 2 | 3 | 5 | 7 | 9 | 16 |
| **2020** | Jan | 2 | 6 | 8 | 1 | 9 | 10 | 0 | 1 | 1 | 3 | 16 | 19 |
| | Feb | 3 | 6 | 9 | 3 | 2 | 5 | 0 | 1 | 1 | 6 | 9 | 15 |
| | Mar | 0 | 6 | 6 | 2 | 3 | 5 | 3 | 1 | 4 | 5 | 10 | 15 |
| | Apr | 1 | 3 | 4 | 0 | 0 | 0 | 0 | 0 | 0 | 1 | 3 | 4 |
| | May | 0 | 1 | 1 | 0 | 1 | 1 | 0 | 0 | 0 | 0 | 2 | 2 |
| | Jun | 1 | 2 | 3 | 1 | 4 | 5 | 1 | 0 | 1 | 3 | 6 | 9 |
| | Jul | 1 | 2 | 3 | 2 | 1 | 3 | 0 | 0 | 0 | 3 | 3 | 6 |
| | Aug | 1 | 1 | 2 | 0 | 1 | 1 | 0 | 0 | 0 | 1 | 2 | 3 |
| | Sep | 1 | 2 | 3 | 0 | 1 | 1 | 0 | 0 | 0 | 1 | 3 | 4 |
| | Oct | 0 | 1 | 1 | 0 | 0 | 0 | 0 | 0 | 0 | 0 | 1 | 1 |
| | Nov | 2 | 3 | 5 | 0 | 2 | 2 | 0 | 3 | 3 | 2 | 8 | 10 |
| | Dec | 0 | 6 | 6 | 0 | 3 | 3 | 0 | 2 | 2 | 0 | 11 | 11 |
| **2021** | Jan | 0 | 4 | 4 | 1 | 3 | 4 | 1 | 3 | 4 | 2 | 10 | 12 |
| | Feb | 0 | 0 | 0 | 0 | 3 | 3 | 0 | 1 | 1 | 0 | 4 | 4 |
| | Mar | 2 | 3 | 5 | 1 | 5 | 6 | 1 | 1 | 2 | 4 | 9 | 13 |
| | Apr | 1 | 4 | 5 | 1 | 2 | 3 | 0 | 3 | 3 | 2 | 9 | 11 |
| | May | 0 | 3 | 3 | 0 | 6 | 6 | 0 | 1 | 1 | 0 | 10 | 10 |
| | Jun | 0 | 3 | 3 | 0 | 6 | 6 | 0 | 5 | 5 | 0 | 14 | 14 |
| | Jul | 2 | 3 | 5 | 5 | 1 | 6 | 3 | 3 | 6 | 10 | 7 | 17 |
| | Aug | 0 | 1 | 1 | 4 | 1 | 5 | 0 | 2 | 2 | 4 | 4 | 8 |
| | Sep | 0 | 2 | 2 | 2 | 1 | 3 | 0 | 1 | 1 | 2 | 4 | 6 |
| | Oct | 0 | 6 | 6 | 1 | 0 | 1 | 3 | 4 | 7 | 4 | 10 | 14 |
| | Nov | 0 | 6 | 6 | 4 | 7 | 11 | 2 | 6 | 8 | 6 | 19 | 25 |
| | Dec | 0 | 5 | 5 | 4 | 1 | 5 | 1 | 5 | 6 | 5 | 11 | 16 |

### Supplementary Table 3: Quarterly incidences

| Year | Quarter | CAAP (Inc. per 1,000) | Pneumococcal bacteremic pneumonia (Inc. per 100,000) | Non-CAAP LRI (Inc. per 1,000) | Non-pneumonia IPD (Inc. per 100,000) |
|---|---|---|---|---|---|
| 2016 | Jan-Mar | 2.8 | 2.5 | 11.7 | 4.7 |
| | Apr-Jun | 1.2 | 2.0 | 9.1 | 2.5 |
| | Jul-Sep | 0.5 | 1.5 | 8.7 | 1.9 |
| | Oct-Dec | 1.7 | 1.1 | 12.5 | 3.5 |
| 2017 | Jan-Mar | 4.2 | 1.1 | 16.4 | 4.0 |
| | Apr-Jun | 0.9 | 1.1 | 9.3 | 2.2 |
| | Jul-Sep | 0.4 | 0.7 | 8.5 | 2.4 |
| | Oct-Dec | 1.8 | 0.9 | 14.6 | 3.1 |
| 2018 | Jan-Mar | 2.8 | 1.0 | 17.4 | 3.9 |
| | Apr-Jun | 1.0 | 1.9 | 10.2 | 2.3 |
| | Jul-Sep | 0.7 | 1.7 | 9.8 | 1.7 |
| | Oct-Dec | 1.5 | 2.2 | 13.9 | 3.8 |
| 2019 | Jan-Mar | 3.6 | 1.2 | 17.2 | 4.4 |
| | Apr-Jun | 1.0 | 1.9 | 10.4 | 3.0 |
| | Jul-Sep | 0.7 | 1.3 | 8.8 | 1.6 |
| | Oct-Dec | 1.6 | 1.4 | 14.2 | 3.4 |
| 2020 | Jan-Mar | 3.0 | 0.8 | 14.4 | 3.8 |
| | Apr-Jun | 0.2 | 0.4 | 5.9 | 1.2 |
| | Jul-Sep | 0.2 | 0.4 | 9.0 | 0.9 |
| | Oct-Dec | 0.1 | 0.5 | 8.7 | 2.2 |
| 2021 | Jan-Mar | 0.3 | 1.4 | 8.8 | 2.5 |
| | Apr-Jun | 1.4 | 1.6 | 12.9 | 3.6 |
| | Jul-Sep | 2.8 | 1.8 | 17.1 | 1.6 |
| | Oct-Dec | 2.2 | 1.5 | 20.6 | 4.3 |

### Supplementary Table 4: IRRs 2020-2021 *vs*. 2016-2019 per quarter; children <5 years

| | CAAP | non-CAAP LRI | Pneumococcal bacteremic pneumonia | Non-pneumonia IPD | Carriage |
|---|---|---|---|---|---|
| 2020 | | | | | |
| Jan-Mar 2020 | 0·86 (0·73 - 1·02) | 0·91 (0·84 - 0·98) | 0·82 (0·40 - 1·66) | 0·92 (0·58 - 1·45) | 1·01 (0·88 - 1·15) |
| Apr-Jun 2020 | 0·22 (0·14 - 0·36) | 0·60 (0·54 - 0·66) | 0·36 (0·11 - 1·13) | 0·49 (0·24 - 1·01) | 0·86 (0·67 - 1·11) |
| Jul-Sep 2020 | 0·35 (0·21 - 0·60) | 0·99 (0·90 - 1·09) | 0·76 (0·24 - 2·44) | 0·47 (0·20 - 1·09) | 1·00 (0·85 - 1·19) |
| Oct-Dec 2020 | 0·09 (0·05 - 0·15) | 0·62 (0·57 - 0·68) | 0·15 (0·03 - 0·64) | 0·64 (0·37 - 1·13) | 0·73 (0·63 - 0·86) |
| 2021 | | | | | |
| Jan-Mar 2021 | 0·09 (0·06 - 0·14) | 0·55 (0·50 - 0·60) | 0·35 (0·14 - 0·88) | 0·60 (0·36 - 1·00) | 0·87 (0·75 - 1·01) |
| Apr-Jun 2021 | 1·26 (0·96 - 1·66) | 1·29 (1·19 - 1·41) | 0·18 (0·04 - 0·81) | 1·45 (0·85 - 2·48) | 0·98 (0·85 - 1·14) |
| Jul-Sep 2021 | 4·49 (3·32 - 6·08) | 1·87 (1·72 - 2·04) | 2·43 (0·98 - 6·03) | 0·87 (0·43 - 1·75) | 1·03 (0·85 - 1·25) |
| Oct-Dec 2021 | 1·27 (1·02 - 1·57) | 1·47 (1·37 - 1·58) | 1·07 (0·52 - 2·24) | 1·27 (0·79 - 2·03) | 0·96 (0·83 - 1·11) |

### Supplementary Table 5: Pneumococcal carriage rates in children <3 years

| | **2016** | **2017** | **2018** | **2019** | **2020*** | **2021** |
|---|---|---|---|---|---|---|
| Jan | 0·43 | 0·5 | 0·51 | 0·46 | 0·53 | 0·44 |
| Feb | 0·51 | 0·52 | 0·51 | 0·45 | 0·48 | 0·39 |
| Mar | 0·42 | 0·46 | 0·53 | 0·49 | 0·42 | 0·46 |
| Apr | 0·48 | 0·43 | 0·38 | 0·40 | - | 0·40 |
| May | 0·48 | 0·56 | 0·48 | 0·42 | - | 0·42 |
| Jun | 0·57 | 0·40 | 0·43 | 0·45 | 0·40 | 0·47 |
| Jul | 0·44 | 0·39 | 0·36 | 0·38 | 0·43 | 0·39 |
| Aug | 0·40 | 0·34 | 0·38 | 0·27 | 0·37 | 0·38 |
| Sep | 0·43 | 0·33 | 0·37 | 0·37 | 0·29 | 0·32 |
| Oct | 0·47 | 0·38 | 0·39 | 0·4 | 0·31 | 0·40 |
| Nov | 0·53 | 0·52 | 0·53 | 0·54 | 0·38 | 0·44 |
| Dec | 0·56 | 0·51 | 0·53 | 0·57 | 0·40 | 0·47 |
| Yearly | 0·48 | 0·45 | 0·46 | 0·43 | 0·4 | 0·42 |

***No data collection during April and May 2020**

### Supplementary Table 6: Monthly number of nasopharyngeal swabs positive for *S. pneumoniae* in children <3 year of age; January 2016 through December 2021; southern Israel, by age group

| | | **<1 year of age** | | | **1 year of age** | | | **2 years of age** | | | **<3 years of age** | | |
|---|---|---|---|---|---|---|---|---|---|---|---|---|---|
| | | Jewish pos/total | Bedouin pos/total | Total pos/total | Jewish pos/total | Bedouin pos/total | Total pos/total | Jewish pos/total | Bedouin pos/total | Total pos/total | Jewish pos/total | Bedouin pos/total | Total pos/total |
| **2016** | Jan | 12/42* | 23/57 | 35/99 | 19/40 | 17/29 | 36/69 | 3/8 | 5/9 | 8/17 | 34/90 | 45/95 | 79/185 |
| | Feb | 14/39 | 18/39 | 32/78 | 14/28 | 26/33 | 40/61 | 7/14 | 12/25 | 19/39 | 35/81 | 56/97 | 91/178 |
| | Mar | 12/47 | 31/67 | 43/114 | 13/34 | 16/25 | 29/59 | 4/8 | 6/13 | 10/21 | 29/89 | 53/105 | 82/194 |
| | Apr | 7/31 | 21/47 | 28/78 | 10/21 | 13/18 | 23/39 | 6/9 | 8/10 | 14/19 | 23/61 | 42/75 | 65/136 |
| | May | 19/48 | 28/57 | 47/105 | 12/27 | 15/33 | 27/60 | 8/12 | 14/21 | 22/33 | 39/87 | 57/111 | 96/198 |
| | Jun | 26/65 | 41/71 | 67/136 | 15/26 | 24/32 | 39/58 | 6/14 | 21/24 | 27/38 | 47/105 | 86/127 | 133/232 |
| | Jul | 20/63 | 26/62 | 46/125 | 17/30 | 17/33 | 34/63 | 9/17 | 10/18 | 19/35 | 46/110 | 53/113 | 99/223 |
| | Aug | 10/47 | 34/65 | 44/112 | 8/22 | 10/24 | 18/46 | 4/14 | 14/28 | 18/42 | 22/83 | 58/117 | 80/200 |
| | Sep | 15/60 | 32/61 | 47/121 | 13/28 | 16/28 | 29/56 | 7/15 | 6/15 | 13/30 | 35/103 | 54/104 | 89/207 |
| | Oct | 8/40 | 22/38 | 30/78 | 13/21 | 9/17 | 22/38 | 5/8 | 5/7 | 10/15 | 26/69 | 36/62 | 62/131 |
| | Nov | 16/56 | 33/70 | 49/126 | 19/30 | 29/38 | 48/68 | 10/14 | 16/23 | 26/37 | 45/100 | 78/131 | 123/231 |
| | Dec | 26/60 | 33/65 | 59/125 | 11/23 | 21/31 | 32/54 | 15/18 | 15/20 | 30/38 | 52/101 | 69/116 | 121/217 |
| **2017** | Jan | 21/55 | 26/62 | 47/117 | 17/32 | 18/33 | 35/65 | 11/18 | 15/17 | 26/35 | 49/105 | 59/112 | 108/217 |
| | Feb | 19/60 | 27/54 | 46/114 | 15/34 | 25/26 | 40/60 | 7/14 | 16/23 | 23/37 | 41/108 | 68/103 | 109/211 |
| | Mar | 17/62 | 31/66 | 48/128 | 20/38 | 18/30 | 38/68 | 10/17 | 8/15 | 18/32 | 47/117 | 57/111 | 104/228 |
| | Apr | 13/56 | 32/63 | 45/119 | 12/29 | 19/34 | 31/63 | 6/9 | 6/13 | 12/22 | 31/94 | 57/110 | 88/204 |
| | May | 16/51 | 45/67 | 61/118 | 18/28 | 25/36 | 43/64 | 6/11 | 11/22 | 17/33 | 40/90 | 81/125 | 121/215 |
| | Jun | 13/46 | 24/66 | 37/112 | 13/23 | 17/31 | 30/54 | 5/6 | 2/12 | 7/18 | 31/75 | 43/109 | 74/184 |
| | Jul | 14/55 | 34/77 | 48/132 | 11/33 | 12/29 | 23/62 | 10/17 | 10/21 | 20/38 | 35/105 | 56/127 | 91/232 |
| | Aug | 11/40 | 19/60 | 30/100 | 7/21 | 11/29 | 18/50 | 8/15 | 7/19 | 15/34 | 26/76 | 37/108 | 63/184 |
| | Sep | 8/41 | 12/58 | 20/99 | 12/26 | 16/30 | 28/56 | 5/16 | 7/12 | 12/28 | 25/83 | 35/100 | 60/183 |
| | Oct | 15/51 | 21/54 | 36/105 | 8/15 | 13/25 | 21/40 | 3/9 | 3/14 | 6/23 | 26/75 | 37/93 | 63/168 |
| | Nov | 22/51 | 42/76 | 64/127 | 12/20 | 12/26 | 24/46 | 12/16 | 6/14 | 18/30 | 46/87 | 60/116 | 106/203 |
| | Dec | 13/43 | 24/41 | 37/84 | 8/14 | 16/26 | 24/40 | 2/8 | 9/10 | 11/18 | 23/65 | 49/77 | 72/142 |
| **2018** | Jan | 16/51 | 42/75 | 58/126 | 11/20 | 14/21 | 25/41 | 5/10 | 7/9 | 12/19 | 32/81 | 63/105 | 95/186 |
| | Feb | 18/49 | 28/53 | 46/102 | 7/14 | 16/25 | 23/39 | 7/8 | 6/12 | 13/20 | 32/71 | 50/90 | 82/161 |
| | Mar | 15/42 | 22/42 | 37/84 | 17/21 | 12/22 | 29/43 | 1/8 | 10/11 | 11/19 | 33/71 | 44/75 | 77/146 |

| | | | | | | | | | | | | | |
|---|---|---|---|---|---|---|---|---|---|---|---|---|---|
| | Apr | 8/29 | 17/42 | 25/71 | 3/8 | 3/6 | 6/14 | 5/9 | 2/5 | 7/14 | 16/46 | 22/53 | 38/99 |
| | May | 18/41 | 25/52 | 43/93 | 12/24 | 8/21 | 20/45 | 6/12 | 8/10 | 14/22 | 36/77 | 41/83 | 77/160 |
| | Jun | 11/42 | 15/30 | 26/72 | 6/13 | 3/9 | 9/22 | 8/11 | 5/7 | 13/18 | 25/66 | 23/46 | 48/112 |
| | Jul | 7/39 | 23/70 | 30/109 | 2/12 | 16/24 | 18/36 | 7/12 | 5/12 | 12/24 | 16/63 | 44/106 | 60/169 |
| | Aug | 10/36 | 12/39 | 22/75 | 9/19 | 9/13 | 18/32 | 3/5 | 1/5 | 4/10 | 22/60 | 22/57 | 44/117 |
| | Sep | 4/27 | 10/25 | 14/52 | 2/3 | 10/17 | 12/20 | 0/3 | 4/7 | 4/10 | 6/33 | 24/49 | 30/82 |
| | Oct | 16/53 | 19/52 | 35/105 | 6/16 | 12/28 | 18/44 | 5/11 | 10/13 | 15/24 | 27/80 | 41/93 | 68/173 |
| | Nov | 19/42 | 30/58 | 49/100 | 9/15 | 11/23 | 20/38 | 8/10 | 7/12 | 15/22 | 36/67 | 48/93 | 84/160 |
| | Dec | 19/47 | 36/67 | 55/114 | 7/12 | 19/27 | 26/39 | 3/8 | 8/11 | 11/19 | 29/67 | 63/105 | 92/172 |
| 2019 | Jan | 17/47 | 16/41 | 33/88 | 9/25 | 17/26 | 26/51 | 6/11 | 10/14 | 16/25 | 32/83 | 43/81 | 75/164 |
| | Feb | 15/35 | 19/55 | 34/90 | 6/13 | 13/19 | 19/32 | 5/11 | 6/8 | 11/19 | 26/59 | 38/82 | 64/141 |
| | Mar | 14/38 | 37/69 | 51/107 | 4/9 | 9/15 | 13/24 | 4/8 | 5/9 | 9/17 | 22/55 | 51/93 | 73/148 |
| | Apr | 14/36 | 11/32 | 25/68 | 8/13 | 4/11 | 12/24 | 2/5 | 1/4 | 3/9 | 24/54 | 16/47 | 40/101 |
| | May | 15/43 | 13/39 | 28/82 | 7/10 | 5/12 | 12/22 | 10/15 | 2/6 | 12/21 | 32/68 | 20/57 | 52/125 |
| | Jun | 9/30 | 18/41 | 27/71 | 11/17 | 8/15 | 19/32 | 4/14 | 5/5 | 9/19 | 24/61 | 31/61 | 55/122 |
| | Jul | 15/42 | 14/41 | 29/83 | 11/18 | 6/16 | 17/34 | 0/4 | 4/9 | 4/13 | 26/64 | 24/66 | 50/130 |
| | Aug | 10/40 | 7/33 | 17/73 | 2/14 | 5/12 | 7/26 | 6/13 | 1/4 | 7/17 | 18/67 | 13/49 | 31/116 |
| | Sep | 10/36 | 10/23 | 20/59 | 6/15 | 1/11 | 7/26 | 6/8 | 4/8 | 10/16 | 22/59 | 15/42 | 37/101 |
| | Oct | 10/41 | 22/51 | 32/92 | 7/13 | 8/19 | 15/32 | 3/6 | 5/9 | 8/15 | 20/60 | 35/79 | 55/139 |
| | Nov | 2/4 | 20/48 | 22/52 | 1/3 | 9/12 | 10/15 | 0/0 | 12/15 | 12/15 | 3/7 | 41/75 | 44/82 |
| | Dec | 9/22 | 23/42 | 32/64 | 10/13 | 14/23 | 24/36 | 1/2 | 7/10 | 8/12 | 20/37 | 44/75 | 64/112 |
| 2020 | Jan | 9/24 | 32/61 | 41/85 | 4/9 | 17/32 | 21/41 | 3/3 | 11/15 | 14/18 | 16/36 | 60/108 | 76/144 |
| | Feb | 8/39 | 26/51 | 34/90 | 12/24 | 13/17 | 25/41 | 3/4 | 7/9 | 10/13 | 23/67 | 46/77 | 69/144 |
| | Mar | 5/14 | 4/14 | 9/28 | 2/5 | 7/7 | 9/12 | 0/1 | 1/4 | 1/5 | 7/20 | 12/25 | 19/45 |
| | Apr** | - | - | - | - | - | - | - | - | - | - | - | - |
| | May** | - | - | - | - | - | - | - | - | - | - | - | - |
| | Jun | 7/30 | 11/32 | 18/62 | 3/7 | 10/13 | 13/20 | 5/10 | 3/6 | 8/16 | 15/47 | 24/51 | 39/98 |
| | Jul | 12/38 | 13/38 | 25/76 | 6/12 | 10/16 | 16/28 | 8/12 | 7/13 | 15/25 | 26/62 | 30/67 | 56/129 |
| | Aug | 9/39 | 16/56 | 25/95 | 8/20 | 11/17 | 19/37 | 2/4 | 7/9 | 9/13 | 19/63 | 34/82 | 53/145 |
| | Sep | 1/23 | 17/49 | 18/72 | 1/11 | 8/11 | 9/22 | 1/4 | 1/2 | 2/6 | 3/38 | 26/62 | 29/100 |
| | Oct | 8/39 | 11/42 | 19/81 | 6/14 | 4/11 | 10/25 | 4/5 | 4/10 | 8/15 | 18/58 | 19/63 | 37/121 |
| | Nov | 7/38 | 14/45 | 21/83 | 11/18 | 7/17 | 18/35 | 5/8 | 11/17 | 16/25 | 23/64 | 32/79 | 55/143 |
| | Dec | 6/27 | 13/39 | 19/66 | 4/11 | 15/23 | 19/34 | 4/11 | 6/10 | 10/21 | 14/49 | 34/72 | 48/121 |
| 2021 | Jan | 3/14 | 14/40 | 17/54 | 9/17 | 11/17 | 20/34 | 1/6 | 9/13 | 10/19 | 13/37 | 34/70 | 47/107 |
| | Feb | 5/28 | 14/37 | 19/65 | 5/11 | 11/22 | 16/33 | 3/6 | 5/7 | 8/13 | 13/45 | 30/66 | 43/111 |
| | Mar | 7/30 | 18/34 | 25/64 | 7/15 | 12/17 | 19/32 | 0/2 | 3/5 | 3/7 | 14/47 | 33/56 | 47/103 |
| | Apr | 10/39 | 19/43 | 29/82 | 6/15 | 6/12 | 12/27 | 4/8 | 6/9 | 10/17 | 20/62 | 31/64 | 51/126 |
| | May | 8/28 | 11/29 | 19/57 | 7/15 | 5/6 | 12/21 | 4/8 | 5/10 | 9/18 | 19/51 | 21/45 | 40/96 |
| | Jun | 10/39 | 33/52 | 43/91 | 9/22 | 8/14 | 17/36 | 4/11 | 5/10 | 9/21 | 23/72 | 46/76 | 69/148 |
| | Jul | 7/38 | 23/42 | 30/80 | 12/31 | 8/12 | 20/43 | 4/10 | 1/7 | 5/17 | 23/79 | 32/61 | 55/140 |
| | Aug | 7/21 | 14/40 | 21/61 | 5/8 | 8/22 | 13/30 | 2/7 | 3/6 | 5/13 | 14/36 | 25/68 | 39/104 |
| | Sep | 2/8 | 5/14 | 7/22 | 0/4 | 2/3 | 2/7 | 1/1 | 0/1 | 1/2 | 3/13 | 7/18 | 10/31 |
| | Oct | 9/40 | 15/33 | 24/73 | 6/9 | 7/16 | 13/25 | 2/4 | 4/6 | 6/10 | 17/53 | 26/55 | 43/108 |
| | Nov | 10/34 | 14/37 | 24/71 | 7/17 | 12/17 | 19/34 | 3/3 | 5/8 | 8/11 | 20/54 | 31/62 | 51/116 |
| | Dec | 7/35 | 28/53 | 35/88 | 8/14 | 13/20 | 21/34 | 3/7 | 3/4 | 6/11 | 18/56 | 44/77 | 62/133 |

* Number of positive samples/number of obtained samples
** During April and May 2021 sampling was temporarily interrupted

**Supplementary Table 7: Mean density of positive nasopharyngeal samples of *S· pneumoniae*, measured by semi quantitative method in children <3 years of age; January 2016 through December 2021; southern Israel, by age group**

| | | <1 year of age | | | 1 year of age | | | 2 years of age | | | <3 years of age | | |
|---|---|---|---|---|---|---|---|---|---|---|---|---|---|
| | | Jewish | Bedouin | Total | Jewish | Bedouin | Total | Jewish | Bedouin | Total | Jewish | Bedouin | Total |
| 2016 | Jan | 2·9 | 2·5 | 2·7 | 2·5 | 2·8 | 2·6 | 3·7 | 3 | 3·3 | 2·8 | 2·7 | 2·7 |
| | Feb | 2·6 | 2·5 | 2·5 | 3·1 | 3·2 | 3·1 | 2·7 | 2·9 | 2·8 | 2·8 | 2·9 | 2·9 |
| | Mar | 2·4 | 2·9 | 2·8 | 2·6 | 3·1 | 2·9 | 1·7 | 2·3 | 2·1 | 2·4 | 2·9 | 2·7 |
| | Apr | 2·6 | 3 | 2·9 | 2·1 | 3·4 | 2·8 | 3·2 | 3·4 | 3·3 | 2·5 | 3·2 | 3 |
| | May | 3·4 | 3·3 | 3·4 | 3·3 | 3·1 | 3·2 | 2·9 | 3·3 | 3·1 | 3·3 | 3·3 | 3·3 |
| | Jun | 3·3 | 3·2 | 3·2 | 3·3 | 3·1 | 3·2 | 3·3 | 3·5 | 3·4 | 3·3 | 3·2 | 3·2 |
| | Jul | 2·6 | 3 | 2·8 | 2·3 | 2·9 | 2·6 | 3 | 3·8 | 3·4 | 2·6 | 3·1 | 2·9 |
| | Aug | 2·5 | 2·6 | 2·5 | 2·8 | 3·4 | 3·1 | 3·8 | 2·1 | 2·5 | 2·8 | 2·6 | 2·7 |
| | Sep | 2·7 | 3·5 | 3·3 | 3·7 | 3·2 | 3·4 | 2·1 | 3·3 | 2·7 | 3 | 3·4 | 3·2 |
| | Oct | 3·5 | 2·7 | 2·9 | 2·5 | 3·6 | 2·9 | 2·8 | 3·4 | 3·1 | 2·8 | 3 | 3 |
| | Nov | 2·7 | 3 | 2·9 | 3·1 | 3·1 | 3·1 | 3·2 | 2·5 | 2·8 | 3 | 2·9 | 2·9 |
| | Dec | 2·4 | 2·8 | 2·6 | 2·8 | 2·1 | 2·4 | 2·1 | 2·6 | 2·3 | 2·4 | 2·5 | 2·5 |

| | | | | | | | | | | | | | |
|---|---|---|---|---|---|---|---|---|---|---|---|---|---|
| **2017** | Jan | 2·5 | 3·1 | 2·8 | 3 | 2·9 | 2·9 | 2·7 | 3·1 | 2·9 | 2·7 | 3 | 2·9 |
| | Feb | 2·2 | 3·2 | 2·8 | 3·1 | 3 | 3·1 | 2·4 | 2·8 | 2·7 | 2·6 | 3·1 | 2·9 |
| | Mar | 1·9 | 2·5 | 2·3 | 2·4 | 2·9 | 2·6 | 2·4 | 3·3 | 2·8 | 2·2 | 2·8 | 2·5 |
| | Apr | 2·8 | 2·8 | 2·8 | 2·3 | 2·8 | 2·6 | 2 | 2·5 | 2·3 | 2·5 | 2·8 | 2·7 |
| | May | 1·9 | 3·2 | 2·8 | 2·7 | 3·1 | 3 | 3·3 | 2·9 | 3·1 | 2·5 | 3·1 | 2·9 |
| | Jun | 2·9 | 3·1 | 3 | 2·6 | 2·7 | 2·7 | 2·6 | 2·5 | 2·6 | 2·7 | 2·9 | 2·8 |
| | Jul | 3·4 | 2·7 | 2·9 | 2·5 | 2·8 | 2·7 | 2·9 | 2·6 | 2·8 | 3 | 2·7 | 2·8 |
| | Aug | 2·8 | 3 | 2·9 | 2·4 | 2·5 | 2·4 | 2·5 | 3·3 | 2·9 | 2·6 | 2·9 | 2·8 |
| | Sep | 3·4 | 2·8 | 3·1 | 3·1 | 3·2 | 3·1 | 3·6 | 4 | 3·8 | 3·3 | 3·2 | 3·3 |
| | Oct | 2·9 | 3·3 | 3·1 | 2·9 | 3·4 | 3·2 | 2 | 4 | 3 | 2·8 | 3·4 | 3·1 |
| | Nov | 2·7 | 2·9 | 2·8 | 2·6 | 3·3 | 3 | 3·4 | 3·3 | 3·4 | 2·9 | 3 | 3 |
| | Dec | 2·3 | 3·1 | 2·8 | 2·8 | 2·3 | 2·4 | 3·5 | 3·3 | 3·4 | 2·6 | 2·9 | 2·8 |
| **2018** | Jan | 3·4 | 2·7 | 2·9 | 3·2 | 3·2 | 3·2 | 2·4 | 2·3 | 2·3 | 3·2 | 2·8 | 2·9 |
| | Feb | 3·1 | 3·5 | 3·3 | 3·3 | 3·4 | 3·3 | 3·9 | 3·2 | 3·5 | 3·3 | 3·4 | 3·4 |
| | Mar | 3·7 | 3·5 | 3·5 | 2·6 | 3·3 | 2·9 | 4 | 2·8 | 2·9 | 3·2 | 3·3 | 3·2 |
| | Apr | 3·3 | 3·3 | 3·3 | 4 | 4 | 4 | 3·2 | 3·5 | 3·3 | 3·4 | 3·4 | 3·4 |
| | May | 3·2 | 2·7 | 2·9 | 3·3 | 2·8 | 3·1 | 4 | 1·8 | 2·7 | 3·3 | 2·5 | 2·9 |
| | Jun | 2·9 | 3 | 3 | 3·3 | 3·7 | 3·4 | 3·1 | 2·4 | 2·8 | 3·1 | 3 | 3 |
| | Jul | 3·3 | 3·1 | 3·1 | 4 | 3·8 | 3·8 | 3·6 | 3·6 | 3·6 | 3·5 | 3·4 | 3·4 |
| | Aug | 3 | 2·9 | 3 | 3·2 | 3·2 | 3·2 | 3 | 4 | 3·3 | 3·1 | 3·1 | 3·1 |
| | Sep | 3 | 3·1 | 3·1 | 3 | 3·3 | 3·3 | - | 2·8 | 2·8 | 3 | 3·1 | 3·1 |
| | Oct | 3·2 | 3·3 | 3·2 | 2·8 | 3·1 | 3 | 3·2 | 3·2 | 3·2 | 3·1 | 3·2 | 3·2 |
| | Nov | 2·9 | 3·1 | 3 | 3·7 | 3·5 | 3·6 | 3·3 | 3·7 | 3·5 | 3·2 | 3·3 | 3·2 |
| | Dec | 2·9 | 2·5 | 2·6 | 3·3 | 2·9 | 3 | 3·7 | 2·8 | 3 | 3·1 | 2·6 | 2·8 |
| **2019** | Jan | 2·9 | 3·2 | 3 | 3·2 | 3·2 | 3·2 | 3·7 | 3·1 | 3·3 | 3·1 | 3·2 | 3·2 |
| | Feb | 2·9 | 2·9 | 2·9 | 2·5 | 3·2 | 3 | 2·8 | 3·5 | 3·2 | 2·8 | 3·1 | 3 |
| | Mar | 2·7 | 3·3 | 3·1 | 3·8 | 3·4 | 3·5 | 3·3 | 3·4 | 3·3 | 3 | 3·3 | 3·2 |
| | Apr | 4 | 2·6 | 3·4 | 3·9 | 3·8 | 3·8 | 2 | 4 | 2·7 | 3·8 | 3 | 3·5 |
| | May | 3·9 | 3·1 | 3·5 | 3 | 3·4 | 3·2 | 3·8 | 3·5 | 3·8 | 3·7 | 3·2 | 3·5 |
| | Jun | 2·9 | 3·3 | 3·2 | 2·8 | 2·9 | 2·8 | 4 | 2·8 | 3·3 | 3 | 3·1 | 3·1 |
| | Jul | 3·9 | 3·4 | 3·7 | 3·7 | 3 | 3·5 | - | 3·5 | 3·5 | 3·8 | 3·3 | 3·6 |
| | Aug | 3·8 | 2·9 | 3·4 | 4 | 4 | 4 | 3·3 | 1 | 3 | 3·7 | 3·2 | 3·5 |
| | Sep | 3·8 | 3·7 | 3·8 | 3·3 | 4 | 3·4 | 3·5 | 3 | 3·3 | 3·6 | 3·5 | 3·6 |
| | Oct | 2·9 | 3·2 | 3·1 | 2·9 | 3·3 | 3·1 | 2·7 | 3·4 | 3·1 | 2·9 | 3·2 | 3·1 |
| | Nov | 2 | 3·7 | 3·5 | 2 | 3·8 | 3·6 | - | 3 | 3 | 2 | 3·5 | 3·4 |
| | Dec | 3·2 | 3·3 | 3·3 | 3·8 | 3·9 | 3·9 | 2 | 3·4 | 3·3 | 3·5 | 3·5 | 3·5 |
| **2020** | Jan | 2·9 | 2·4 | 2·5 | 1·5 | 2·9 | 2·7 | 2·7 | 3·1 | 3 | 2·5 | 2·7 | 2·6 |
| | Feb | 3·6 | 3·4 | 3·4 | 2·8 | 3·5 | 3·2 | 3 | 3·3 | 3·2 | 3·1 | 3·4 | 3·3 |
| | Mar | 3 | 3 | 3 | 3 | 2·9 | 2·9 | - | 4 | 4 | 3 | 3 | 3 |
| | Apr* | - | - | - | - | - | - | - | - | - | - | - | - |
| | May* | - | - | - | - | - | - | - | - | - | - | - | - |
| | Jun | 4 | 3·4 | 3·6 | 2·3 | 3·6 | 3·3 | 3·4 | 4 | 3·6 | 3·5 | 3·5 | 3·5 |
| | Jul | 3·8 | 3·7 | 3·8 | 3·7 | 3 | 3·3 | 4 | 3·7 | 3·9 | 3·8 | 3·5 | 3·6 |
| | Aug | 3·7 | 3·6 | 3·6 | 3·9 | 3·7 | 3·8 | 4 | 3 | 3·3 | 3·8 | 3·5 | 3·6 |
| | Sep | 3 | 3·5 | 3·5 | 4 | 3·3 | 3·3 | 4 | 4 | 4 | 3·7 | 3·5 | 3·5 |
| | Oct | 3·6 | 3·6 | 3·6 | 3·7 | 2·8 | 3·3 | 3·8 | 3·5 | 3·6 | 3·7 | 3·4 | 3·5 |
| | Nov | 2·9 | 3·8 | 3·5 | 2·7 | 3·7 | 3·1 | 3·8 | 3·9 | 3·9 | 3 | 3·8 | 3·5 |
| | Dec | 3 | 2·9 | 2·9 | 3·3 | 2·7 | 2·8 | 3·5 | 2·8 | 3·1 | 3·2 | 2·8 | 2·9 |
| **2021** | Jan | 2·7 | 3·8 | 3·6 | 3·1 | 2·8 | 2·9 | 3 | 3·3 | 3·3 | 3 | 3·4 | 3·3 |
| | Feb | 2·2 | 3·2 | 2·9 | 2·2 | 3·2 | 2·9 | 3·3 | 3·8 | 3·6 | 2·5 | 3·3 | 3 |
| | Mar | 3·7 | 3·8 | 3·8 | 3·6 | 3·8 | 3·7 | - | 4 | 4 | 3·6 | 3·8 | 3·7 |
| | Apr | 3·1 | 3·6 | 3·4 | 3·5 | 3·8 | 3·6 | 3 | 2·7 | 2·8 | 3·2 | 3·4 | 3·3 |
| | May | 3·8 | 3·9 | 3·8 | 4 | 3·4 | 3·8 | 3·3 | 4 | 3·7 | 3·7 | 3·8 | 3·8 |
| | Jun | 3·6 | 3·6 | 3·6 | 3·7 | 3·6 | 3·6 | 4 | 4 | 4 | 3·7 | 3·6 | 3·7 |
| | Jul | 3·4 | 3·5 | 3·5 | 3·3 | 3·8 | 3·5 | 3 | 4 | 3·2 | 3·3 | 3·6 | 3·4 |
| | Aug | 3·6 | 3·6 | 3·6 | 3·4 | 3·4 | 3·4 | 4 | 4 | 4 | 3·6 | 3·6 | 3·6 |
| | Sep | 4 | 4 | 4 | -! | 3 | 3 | 4 | - | 4 | 4 | 3·7 | 3·8 |
| | Oct | 2·8 | 3·1 | 3 | 3·4 | 3·7 | 3·6 | 3·5 | 3 | 3·2 | 3·1 | 3·3 | 3·2 |
| | Nov | 3 | 3·4 | 3·2 | 3·7 | 3·1 | 3·3 | 2·3 | 3·4 | 3 | 3·2 | 3·3 | 3·2 |
| | Dec | 3·3 | 3·5 | 3·4 | 2·3 | 3·2 | 2·8 | 3 | 2·3 | 2·7 | 2·8 | 3·3 | 3·1 |

*During April and May 2021 sampling was temporarily interrupted

### Supplementary Table 8: Pneumococcal serotype distribution per study year

| | **2016** | | **2017** | | **2018** | | **2019** | | **2016-2019** | | **2020-2021** | |
|---|---|---|---|---|---|---|---|---|---|---|---|---|
| 15B/C | 128 | 11·43% | 142 | 13·42% | 94 | 11·84% | 83 | 12·97% | 447 | 12·38% | 113 | 10·88% |
| 23B | 47 | 4·20% | 65 | 6·14% | 65 | 8·19% | 42 | 6·56% | 219 | 6·06% | 94 | 9·05% |
| 11A | 55 | 4·91% | 60 | 5·67% | 54 | 6·80% | 42 | 6·56% | 211 | 5·84% | 74 | 7·12% |
| 16F | 55 | 4·91% | 55 | 5·20% | 43 | 5·42% | 36 | 5·63% | 189 | 5·23% | 66 | 6·35% |
| 35B | 53 | 4·73% | 56 | 5·29% | 40 | 5·04% | 35 | 5·47% | 184 | 5·09% | 48 | 4·62% |

| | | | | | | | | | | | | |
|---|---|---|---|---|---|---|---|---|---|---|---|---|
| 21 | 40 | 3·57% | 61 | 5·77% | 30 | 3·78% | 32 | 5·00% | 163 | 4·51% | 58 | 5·58% |
| 15A | 44 | 3·93% | 35 | 3·31% | 46 | 5·79% | 26 | 4·06% | 151 | 4·18% | 46 | 4·43% |
| 9N | 47 | 4·20% | 36 | 3·40% | 39 | 4·91% | 20 | 3·13% | 142 | 3·93% | 35 | 3·37% |
| 17F | 48 | 4·29% | 32 | 3·02% | 33 | 4·16% | 26 | 4·06% | 139 | 3·85% | 28 | 2·69% |
| 23A | 45 | 4·02% | 35 | 3·31% | 23 | 2·90% | 23 | 3·59% | 126 | 3·49% | 38 | 3·66% |
| 10A | 29 | 2·59% | 34 | 3·21% | 26 | 3·27% | 17 | 2·66% | 106 | 2·93% | 30 | 2·89% |
| 10B | 30 | 2·68% | 20 | 1·89% | 21 | 2·64% | 24 | 3·75% | 95 | 2·63% | 20 | 1·92% |
| 19F | 24 | 2·14% | 29 | 2·74% | 16 | 2·02% | 25 | 3·91% | 94 | 2·60% | 35 | 3·37% |
| 34 | 29 | 2·59% | 28 | 2·65% | 15 | 1·89% | 17 | 2·66% | 89 | 2·46% | 41 | 3·95% |
| 19A | 30 | 2·68% | 24 | 2·27% | 18 | 2·27% | 6 | 0·94% | 78 | 2·16% | 22 | 2·12% |
| 22F | 36 | 3·21% | 22 | 2·08% | 13 | 1·64% | 4 | 0·63% | 75 | 2·08% | 12 | 1·15% |
| 31 | 18 | 1·61% | 20 | 1·89% | 14 | 1·76% | 10 | 1·56% | 62 | 1·72% | 16 | 1·54% |
| 14 | 28 | 2·50% | 17 | 1·61% | 13 | 1·64% | 3 | 0·47% | 61 | 1·69% | 14 | 1·35% |
| 24F | 17 | 1·52% | 14 | 1·32% | 21 | 2·64% | 8 | 1·25% | 60 | 1·66% | 14 | 1·35% |
| 13 | 15 | 1·34% | 12 | 1·13% | 18 | 2·27% | 9 | 1·41% | 54 | 1·50% | 12 | 1·15% |
| 33F | 12 | 1·07% | 13 | 1·23% | 12 | 1·51% | 15 | 2·34% | 52 | 1·44% | 12 | 1·15% |
| 7B | 16 | 1·43% | 13 | 1·23% | 9 | 1·13% | 12 | 1·88% | 50 | 1·38% | 11 | 1·06% |
| 38 | 19 | 1·70% | 12 | 1·13% | 8 | 1·01% | 9 | 1·41% | 48 | 1·33% | 10 | 0·96% |
| 19B | 10 | 0·89% | 9 | 0·85% | 12 | 1·51% | 15 | 2·34% | 46 | 1·27% | 20 | 1·92% |
| 6C | 13 | 1·16% | 11 | 1·04% | 8 | 1·01% | 11 | 1·72% | 43 | 1·19% | 20 | 1·92% |
| 12F | 13 | 1·16% | 13 | 1·23% | 8 | 1·01% | 7 | 1·09% | 41 | 1·14% | 4 | 0·38% |
| 23F | 14 | 1·25% | 9 | 0·85% | 4 | 0·50% | 2 | 0·31% | 29 | 0·80% | 11 | 1·06% |
| 3 | 9 | 0·80% | 7 | 0·66% | 7 | 0·88% | 3 | 0·47% | 26 | 0·72% | 6 | 0·58% |
| 10F | 11 | 0·98% | 10 | 0·95% | 4 | 0·50% | 0 | 0·00% | 25 | 0·69% | 5 | 0·48% |
| 35F | 13 | 1·16% | 4 | 0·38% | 5 | 0·63% | 1 | 0·16% | 23 | 0·64% | 8 | 0·77% |
| 20 | 7 | 0·63% | 4 | 0·38% | 5 | 0·63% | 4 | 0·63% | 20 | 0·55% | 7 | 0·67% |
| 18C | 6 | 0·54% | 8 | 0·76% | 0 | 0·00% | 2 | 0·31% | 16 | 0·44% | 3 | 0·29% |
| 35A | 4 | 0·36% | 5 | 0·47% | 2 | 0·25% | 1 | 0·16% | 12 | 0·33% | 5 | 0·48% |
| 6A | 4 | 0·36% | 6 | 0·57% | 1 | 0·13% | 0 | 0·00% | 11 | 0·30% | 2 | 0·19% |
| 8 | 4 | 0·36% | 4 | 0·38% | 1 | 0·13% | 1 | 0·16% | 10 | 0·28% | 3 | 0·29% |
| 33A | 0 | 0·00% | 9 | 0·85% | 0 | 0·00% | 1 | 0·16% | 10 | 0·28% | 0 | 0·00% |
| 11B | 2 | 0·18% | 0 | 0·00% | 3 | 0·38% | 3 | 0·47% | 8 | 0·22% | 1 | 0·10% |
| 28A | 3 | 0·27% | 2 | 0·19% | 1 | 0·13% | 1 | 0·16% | 7 | 0·19% | 3 | 0·29% |
| 2 | 2 | 0·18% | 0 | 0·00% | 2 | 0·25% | 1 | 0·16% | 5 | 0·14% | 0 | 0·00% |
| 33B | 2 | 0·18% | 0 | 0·00% | 2 | 0·25% | 1 | 0·16% | 5 | 0·14% | 1 | 0·10% |
| 35B/D | 1 | 0·09% | 2 | 0·19% | 1 | 0·13% | 1 | 0·16% | 5 | 0·14% | 1 | 0·10% |
| 7C | 0 | 0·00% | 3 | 0·28% | 1 | 0·13% | 1 | 0·16% | 5 | 0·14% | 2 | 0·19% |
| 18A | 2 | 0·18% | 1 | 0·09% | 0 | 0·00% | 1 | 0·16% | 4 | 0·11% | 1 | 0·10% |
| 27 | 1 | 0·09% | 3 | 0·28% | 0 | 0·00% | 0 | 0·00% | 4 | 0·11% | 0 | 0·00% |
| **Serotype** | **2016** | | **2017** | | **2018** | | **2019** | | **2016-2019** | | **2020-2021** | |
| 6B | 0 | 0·00% | 3 | 0·28% | 1 | 0·13% | 0 | 0·00% | 4 | 0·11% | 0 | 0·00% |
| 9V | 2 | 0·18% | 0 | 0·00% | 1 | 0·13% | 0 | 0·00% | 3 | 0·08% | 0 | 0·00% |
| 35C | 1 | 0·09% | 1 | 0·09% | 1 | 0·13% | 0 | 0·00% | 3 | 0·08% | 1 | 0·10% |
| 24B | 1 | 0·09% | 0 | 0·00% | 0 | 0·00% | 1 | 0·16% | 2 | 0·06% | 0 | 0·00% |
| 29 | 1 | 0·09% | 0 | 0·00% | 0 | 0·00% | 0 | 0·00% | 1 | 0·03% | 0 | 0·00% |
| 36 | 1 | 0·09% | 0 | 0·00% | 0 | 0·00% | 0 | 0·00% | 1 | 0·03% | 0 | 0·00% |
| 22A | 0 | 0·00% | 1 | 0·09% | 0 | 0·00% | 0 | 0·00% | 1 | 0·03% | 0 | 0·00% |
| 4 | 0 | 0·00% | 1 | 0·09% | 0 | 0·00% | 0 | 0·00% | 1 | 0·03% | 0 | 0·00% |

| | | | | | | | | | | | | |
|---|---|---|---|---|---|---|---|---|---|---|---|---|
| 1 | 0 | 0·00% | 0 | 0·00% | 1 | 0·13% | 0 | 0·00% | 1 | 0·03% | 1 | 0·10% |
| 25A | 0 | 0·00% | 0 | 0·00% | 1 | 0·13% | 0 | 0·00% | 1 | 0·03% | 2 | 0·19% |
| 35D | 0 | 0·00% | 0 | 0·00% | 1 | 0·13% | 0 | 0·00% | 1 | 0·03% | 1 | 0·10% |
| 37 | 0 | 0·00% | 0 | 0·00% | 0 | 0·00% | 1 | 0·16% | 1 | 0·03% | 0 | 0·00% |
| OMNI NEG | 128 | 11·43% | 107 | 10·11% | 50 | 6·30% | 57 | 8·91% | 342 | 9·47% | 82 | 7·89% |
| **Total** | **1120** | | **1058** | | **794** | | **640** | | **3612** | | **1039** | |

### Supplementary Table 9: Number of detected virus samples per month and year

| **Year** | **Month** | **RSV** | **Influenza A** | **Influenza B** | **hMPV** | **Respiratory Adenovirus** | **Paraflu** | **Rhinovirus*** | **Total** |
|---|---|---|---|---|---|---|---|---|---|
| **2016** | Jan | 154 | 49 | 11 | 15 | 5 | 49 | - | 283 |
| | Feb | 73 | 24 | 44 | 14 | 4 | 36 | - | 195 |
| | Mar | 14 | 7 | 31 | 22 | 19 | 37 | - | 130 |
| | Apr | 10 | 1 | 6 | 14 | 13 | 45 | - | 89 |
| | May | 5 | 1 | 2 | 6 | 10 | 39 | - | 63 |
| | Jun | 1 | 0 | 0 | 5 | 14 | 39 | - | 59 |
| | Jul | 0 | 0 | 1 | 0 | 13 | 32 | - | 46 |
| | Aug | 0 | 0 | 0 | 3 | 13 | 35 | - | 51 |
| | Sep | 2 | 1 | 0 | 1 | 9 | 22 | - | 35 |
| | Oct | 4 | 1 | 0 | 0 | 17 | 40 | - | 62 |
| | Nov | 31 | 2 | 0 | 1 | 21 | 50 | - | 105 |
| | Dec | 106 | 32 | 0 | 0 | 20 | 51 | - | 209 |
| **2017** | Jan | 159 | 51 | 0 | 18 | 17 | 63 | - | 308 |
| | Feb | 119 | 18 | 0 | 53 | 9 | 55 | - | 254 |
| | Mar | 53 | 5 | 0 | 81 | 17 | 66 | - | 222 |
| | Apr | 9 | 1 | 1 | 24 | 5 | 34 | - | 74 |
| | May | 1 | 1 | 1 | 1 | 11 | 53 | - | 68 |
| | Jun | 0 | 0 | 4 | 0 | 13 | 44 | - | 61 |
| | Jul | 0 | 1 | 0 | 0 | 16 | 28 | - | 45 |
| | Aug | 0 | 0 | 1 | 0 | 10 | 19 | - | 30 |
| | Sep | 2 | 0 | 0 | 0 | 10 | 14 | - | 26 |
| | Oct | 5 | 0 | 3 | 0 | 22 | 23 | - | 53 |
| | Nov | 37 | 0 | 3 | 0 | 14 | 44 | - | 98 |
| | Dec | 200 | 17 | 30 | 0 | 14 | 63 | - | 324 |
| **2018** | Jan | 218 | 36 | 21 | 9 | 16 | 36 | - | 336 |
| | Feb | 218 | 77 | 17 | 34 | 13 | 39 | - | 398 |
| | Mar | 90 | 48 | 1 | 44 | 15 | 36 | - | 234 |
| | Apr | 15 | 9 | 1 | 16 | 4 | 36 | - | 81 |
| | May | 1 | 9 | 0 | 18 | 7 | 29 | - | 64 |
| | Jun | 1 | 5 | 0 | 6 | 10 | 16 | - | 38 |
| | Jul | 0 | 3 | 0 | 5 | 14 | 23 | - | 45 |
| | Aug | 0 | 1 | 0 | 0 | 8 | 19 | - | 28 |
| | Sep | 0 | 0 | 0 | 0 | 7 | 14 | - | 21 |
| | Oct | 0 | 1 | 0 | 2 | 29 | 35 | - | 67 |
| | Nov | 7 | 3 | 0 | 2 | 23 | 27 | - | 62 |
| | Dec | 83 | 23 | 0 | 37 | 22 | 27 | - | 192 |
| **2019** | Jan | 211 | 75 | 0 | 34 | 12 | 20 | - | 352 |
| | Feb | 182 | 128 | 0 | 34 | 8 | 51 | 116 | 519 |
| | Mar | 163 | 44 | 0 | 23 | 7 | 38 | 134 | 409 |
| | Apr | 62 | 3 | 0 | 18 | 5 | 45 | 171 | 304 |
| | May | 10 | 0 | 1 | 2 | 4 | 32 | 114 | 163 |
| | Jun | 2 | 0 | 1 | 0 | 20 | 36 | 165 | 224 |
| | Jul | 0 | 0 | 0 | 0 | 22 | 20 | 143 | 185 |
| | Aug | 0 | 1 | 0 | 0 | 19 | 23 | 84 | 127 |
| | Sep | 0 | 0 | 0 | 0 | 27 | 29 | 161 | 217 |
| | Oct | 10 | 0 | 1 | 0 | 37 | 41 | 145 | 234 |
| | Nov | 53 | 5 | 0 | 1 | 28 | 51 | 177 | 315 |
| | Dec | 205 | 70 | 7 | 1 | 33 | 62 | 211 | 589 |
| **2020** | Jan | 254 | 87 | 45 | 1 | 23 | 27 | 138 | 575 |
| | Feb | 170 | 11 | 64 | 6 | 18 | 37 | 160 | 466 |
| | Mar | 84 | 4 | 10 | 14 | 10 | 23 | 83 | 228 |
| | Apr | 3 | 0 | 0 | 2 | 0 | 5 | 19 | 29 |
| | May | 0 | 0 | 0 | 1 | 0 | 7 | 27 | 35 |
| | Jun | 0 | 0 | 0 | 0 | 0 | 27 | 80 | 107 |
| | Jul | 0 | 0 | 0 | 0 | 0 | 29 | 82 | 111 |
| | Aug | 2 | 0 | 0 | 0 | 0 | 26 | 80 | 108 |
| | Sep | 1 | 0 | 0 | 0 | 0 | 18 | 123 | 142 |
| | Oct | 0 | 0 | 0 | 0 | 1 | 17 | 59 | 77 |
| | Nov | 1 | 0 | 0 | 0 | 3 | 51 | 136 | 191 |

| | | | | | | | | | |
|---|---|---|---|---|---|---|---|---|---|
| | Dec | 0 | 0 | 0 | 0 | 72 | 63 | 146 | 281 |
| **2021** | Jan | 0 | 0 | 0 | 0 | 64 | 49 | 97 | 210 |
| | Feb | 0 | 0 | 0 | 2 | 11 | 48 | 88 | 149 |
| | Mar | 0 | 0 | 0 | 19 | 11 | 50 | 162 | 242 |
| | Apr | 0 | 0 | 0 | 34 | 12 | 57 | 163 | 266 |
| | May | 3 | 0 | 0 | 52 | 19 | 72 | 152 | 298 |
| | Jun | 35 | 0 | 0 | 103 | 16 | 88 | 232 | 474 |
| | Jul | 148 | 0 | 1 | 76 | 30 | 96 | 211 | 562 |
| | Aug | 149 | 1 | 0 | 21 | 9 | 96 | 144 | 420 |
| | Sep | 143 | 4 | 0 | 3 | 13 | 66 | 178 | 407 |
| | Oct | 217 | 6 | 0 | 1 | 18 | 59 | 234 | 535 |
| | Nov | 183 | 38 | 0 | 1 | 58 | 129 | 237 | 646 |
| | Dec | 118 | 122 | 0 | 3 | 49 | 143 | 193 | 628 |

***Rhinovirus testing started in February 2019**

### Supplementary Table 10: Proportions of detected viruses out of all virus-positive samples

| | Jan 2016 – Jan 2019 n=4,577 | | Feb 2019 – Dec 2021 n=8,631 | |
|---|---|---|---|---|
| Single PDA | 3,165 | 69·2% | 2,717 | 31·5% |
| Single AdV | 1,189 | 26·0% | 738 | 8·6% |
| Single RhV | - | - | 3,460 | 40·1% |
| PDA + PDA | 74 | 1·6% | 92 | 1·1% |
| PDA + AdV | 149 | 3·3% | 239 | 2·8% |
| PDA + RhV | - | - | 651 | 7·5% |
| RhV + AdV | - | - | 632 | 7·3% |
| PDA + RhV + AdV | - | - | 102 | 1·2% |

### Supplementary Table 11: Percent of CAAP attributable to RSV, hMPV, Flu, AdV, PIV (adjusted for ethnicity and age) among children <5 years, based on models fit to pre-pandemic data only

| **Virus** | **CAAP** |
|---|---|
| Any | 46 (34,58) |
| RSV | 32 (25,39) |
| hMPV | 5 (0,10) |
| Flu | 4 (0,8) |
| AdV | 1 (0,7) |
| PIV | 3 (0,8) |
