## Supplementary Figures for "The COVID-19 Pandemic as an Opportunity for Unravelling the Causative Association between Respiratory Viruses and Pneumococcus-Associated Disease in Young Children: A Prospective Study"

### Slide 1
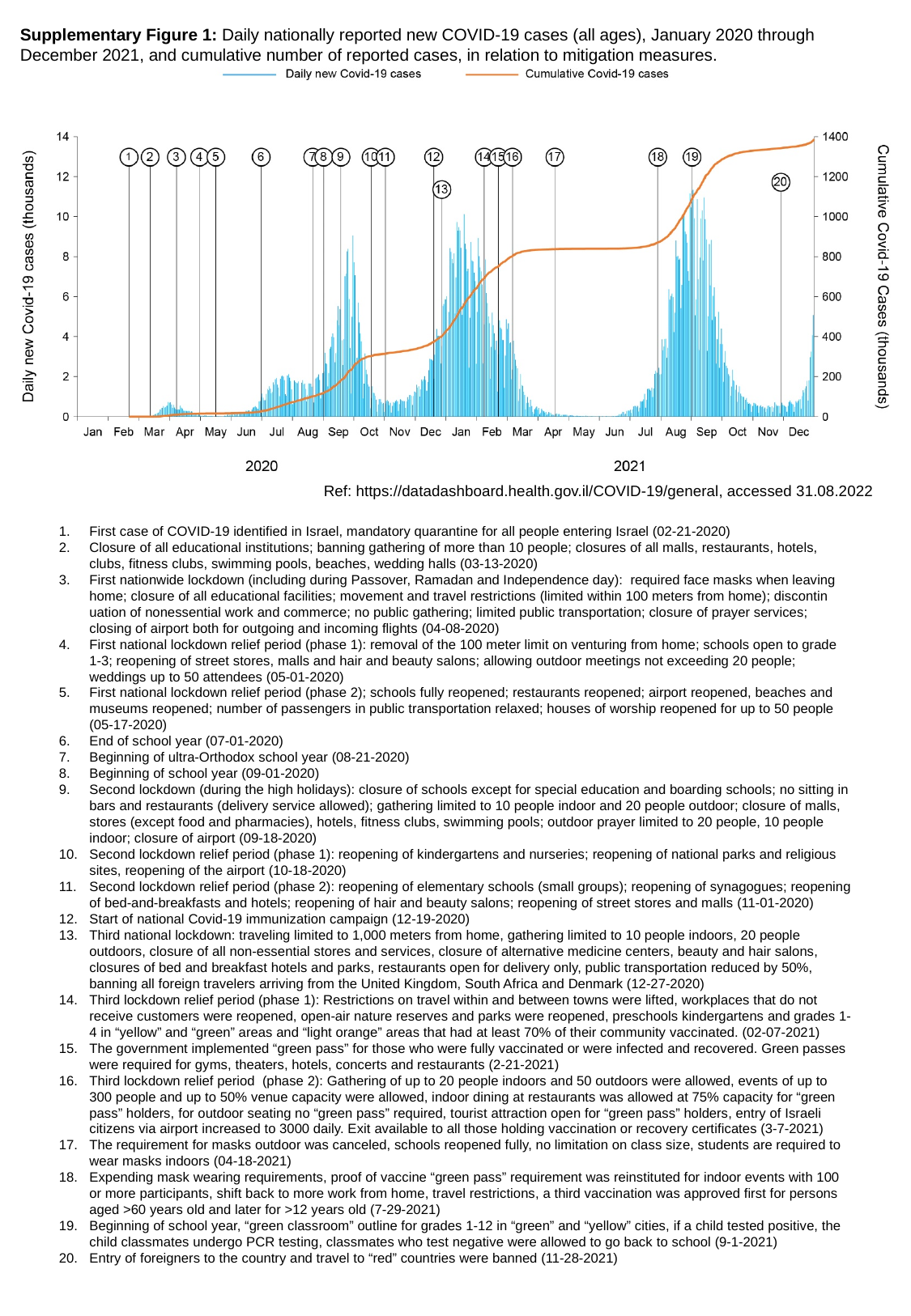

Supplementary Figure 1: Daily nationally reported new COVID-19 cases (all ages), January 2020 through December 2021, and cumulative number of reported cases, in relation to mitigation measures.
Ref: https://datadashboard.health.gov.il/COVID-19/general, accessed 31.08.2022
First case of COVID-19 identified in Israel, mandatory quarantine for all people entering Israel (02-21-2020)
Closure of all educational institutions; banning gathering of more than 10 people; closures of all malls, restaurants, hotels, clubs, fitness clubs, swimming pools, beaches, wedding halls (03-13-2020)
First nationwide lockdown (including during Passover, Ramadan and Independence day): required face masks when leaving home; closure of all educational facilities; movement and travel restrictions (limited within 100 meters from home); discontin­uation of nonessential work and commerce; no public gathering; limited public transportation; closure of prayer services; closing of airport both for outgoing and incoming flights (04-08-2020)
First national lockdown relief period (phase 1): removal of the 100 meter limit on venturing from home; schools open to grade 1-3; reopening of street stores, malls and hair and beauty salons; allowing outdoor meetings not exceeding 20 people; weddings up to 50 attendees (05-01-2020)
First national lockdown relief period (phase 2); schools fully reopened; restaurants reopened; airport reopened, beaches and museums reopened; number of passengers in public transportation relaxed; houses of worship reopened for up to 50 people (05-17-2020)
End of school year (07-01-2020)
Beginning of ultra-Orthodox school year (08-21-2020)
Beginning of school year (09-01-2020)
Second lockdown (during the high holidays): closure of schools except for special education and boarding schools; no sitting in bars and restaurants (delivery service allowed); gathering limited to 10 people indoor and 20 people outdoor; closure of malls, stores (except food and pharmacies), hotels, fitness clubs, swimming pools; outdoor prayer limited to 20 people, 10 people indoor; closure of airport (09-18-2020)
Second lockdown relief period (phase 1): reopening of kindergartens and nurseries; reopening of national parks and religious sites, reopening of the airport (10-18-2020)
Second lockdown relief period (phase 2): reopening of elementary schools (small groups); reopening of synagogues; reopening of bed-and-breakfasts and hotels; reopening of hair and beauty salons; reopening of street stores and malls (11-01-2020)
Start of national Covid-19 immunization campaign (12-19-2020)
Third national lockdown: traveling limited to 1,000 meters from home, gathering limited to 10 people indoors, 20 people outdoors, closure of all non-essential stores and services, closure of alternative medicine centers, beauty and hair salons, closures of bed and breakfast hotels and parks, restaurants open for delivery only, public transportation reduced by 50%, banning all foreign travelers arriving from the United Kingdom, South Africa and Denmark (12-27-2020)
Third lockdown relief period (phase 1): Restrictions on travel within and between towns were lifted, workplaces that do not receive customers were reopened, open-air nature reserves and parks were reopened, preschools kindergartens and grades 1-4 in “yellow” and “green” areas and “light orange” areas that had at least 70% of their community vaccinated. (02-07-2021)
The government implemented “green pass” for those who were fully vaccinated or were infected and recovered. Green passes were required for gyms, theaters, hotels, concerts and restaurants (2-21-2021)
Third lockdown relief period (phase 2): Gathering of up to 20 people indoors and 50 outdoors were allowed, events of up to 300 people and up to 50% venue capacity were allowed, indoor dining at restaurants was allowed at 75% capacity for “green pass” holders, for outdoor seating no “green pass” required, tourist attraction open for “green pass” holders, entry of Israeli citizens via airport increased to 3000 daily. Exit available to all those holding vaccination or recovery certificates (3-7-2021)
The requirement for masks outdoor was canceled, schools reopened fully, no limitation on class size, students are required to wear masks indoors (04-18-2021)
Expending mask wearing requirements, proof of vaccine “green pass” requirement was reinstituted for indoor events with 100 or more participants, shift back to more work from home, travel restrictions, a third vaccination was approved first for persons aged >60 years old and later for >12 years old (7-29-2021)
Beginning of school year, “green classroom” outline for grades 1-12 in “green” and “yellow” cities, if a child tested positive, the child classmates undergo PCR testing, classmates who test negative were allowed to go back to school (9-1-2021)
Entry of foreigners to the country and travel to “red” countries were banned (11-28-2021)

### Slide 2
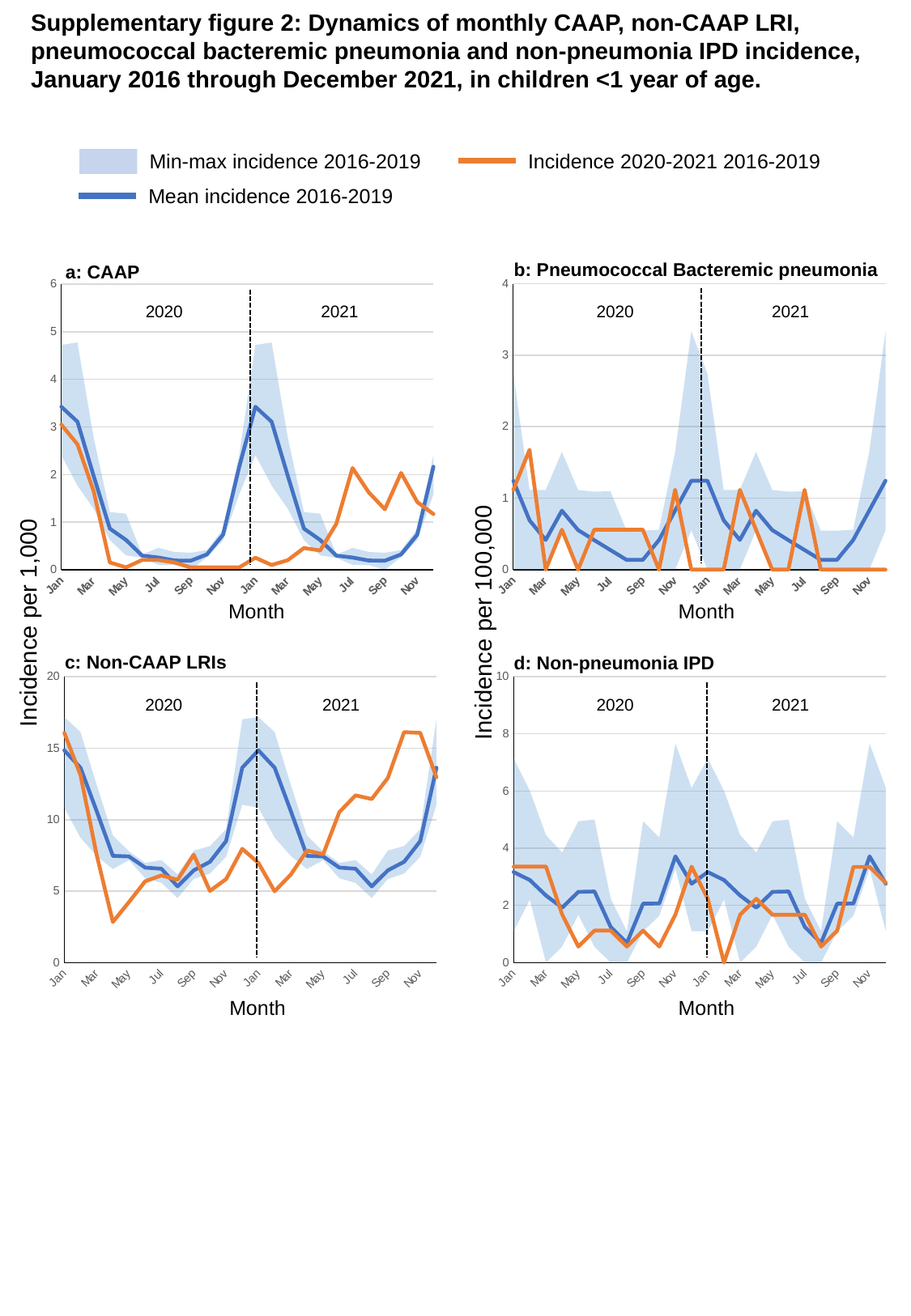

Supplementary figure 2: Dynamics of monthly CAAP, non-CAAP LRI,
pneumococcal bacteremic pneumonia and non-pneumonia IPD incidence,
January 2016 through December 2021, in children <1 year of age.
Min-max incidence 2016-2019
Incidence 2020-2021 2016-2019
Mean incidence 2016-2019
b: Pneumococcal Bacteremic pneumonia
a: CAAP
#### Chart
| Category | Min | Min-Max bacteremic pneumonia 2016-2019 | Mean bacteremic pneumonia 2016-2019 | Bacteremic pneumonia 2020-2021 |
|---|---|---|---|---|
| Jan | 0.0 | 2.741228070175439 | 1.242410081610712 | 1.1179429849077698 |
| Feb | 0.0 | 1.1142061281337048 | 0.6897419053076557 | 1.6769144773616544 |
| Mar | 0.0 | 1.1142061281337048 | 0.41486559964520375 | 0.0 |
| Apr | 0.5482456140350876 | 1.0995052226498077 | 0.823841610411492 | 0.5589714924538849 |
| May | 0.0 | 1.1142061281337048 | 0.5530510883734241 | 0.0 |
| Jun | 0.0 | 1.0905125408942202 | 0.41190390124026816 | 0.5589714924538849 |
| Jul | 0.0 | 1.0964912280701753 | 0.2741228070175438 | 0.5589714924538849 |
| Aug | 0.0 | 0.5452562704471101 | 0.13631406761177753 | 0.5589714924538849 |
| Sep | 0.0 | 0.5452562704471101 | 0.13631406761177753 | 0.5589714924538849 |
| Oct | 0.0 | 0.5571030640668524 | 0.4130279864597166 | 0.0 |
| Nov | 0.0 | 1.6492578339747113 | 0.8271800581388815 | 1.1179429849077698 |
| Dec | 0.5482456140350876 | 2.785515320334262 | 1.2435065216471184 | 0.0 |
| Jan | 0.0 | 2.741228070175439 | 1.242410081610712 | 0.0 |
| Feb | 0.0 | 1.1142061281337048 | 0.6897419053076557 | 0.0 |
| Mar | 0.0 | 1.1142061281337048 | 0.41486559964520375 | 1.1154489682097044 |
| Apr | 0.5482456140350876 | 1.0995052226498077 | 0.823841610411492 | 0.5577244841048522 |
| May | 0.0 | 1.1142061281337048 | 0.5530510883734241 | 0.0 |
| Jun | 0.0 | 1.0905125408942202 | 0.41190390124026816 | 0.0 |
| Jul | 0.0 | 1.0964912280701753 | 0.2741228070175438 | 1.1154489682097044 |
| Aug | 0.0 | 0.5452562704471101 | 0.13631406761177753 | 0.0 |
| Sep | 0.0 | 0.5452562704471101 | 0.13631406761177753 | 0.0 |
| Oct | 0.0 | 0.5571030640668524 | 0.4130279864597166 | 0.0 |
| Nov | 0.0 | 1.6492578339747113 | 0.8271800581388815 | 0.0 |
| Dec | 0.5482456140350876 | 2.785515320334262 | 1.2435065216471184 | 0.0 |
#### Chart
| Category | Min | Min-Max bacteremic pneumonia 2016-2019 | Mean bacteremic pneumonia 2016-2019 | Bacteremic pneumonia 2020-2021 |
|---|---|---|---|---|
| Jan | 2.406677 | 2.313974 | 3.423585 | 3.047678 |
| Feb | 1.768584 | 3.003943 | 3.11033 | 2.634434 |
| Mar | 1.280147 | 1.497209 | 1.972698 | 1.652978 |
| Apr | 0.622504 | 0.589434 | 0.865006 | 0.154967 |
| May | 0.302984 | 0.876071 | 0.627878 | 0.051656 |
| Jun | 0.252487 | 0.069074 | 0.298134 | 0.206622 |
| Jul | 0.103751 | 0.357103 | 0.257866 | 0.206622 |
| Aug | 0.100995 | 0.274159 | 0.196348 | 0.154967 |
| Sep | 0.0 | 0.358441 | 0.192327 | 0.051656 |
| Oct | 0.267967 | 0.147035 | 0.323297 | 0.051656 |
| Nov | 0.622504 | 0.234992 | 0.738586 | 0.051656 |
| Dec | 1.587383 | 0.824322 | 2.164089 | 0.051656 |
| Jan | 2.406677 | 2.313974 | 3.423585 | 0.254285 |
| Feb | 1.768584 | 3.003943 | 3.11033 | 0.101714 |
| Mar | 1.280147 | 1.497209 | 1.972698 | 0.203428 |
| Apr | 0.622504 | 0.589434 | 0.865006 | 0.457712 |
| May | 0.302984 | 0.876071 | 0.627878 | 0.406856 |
| Jun | 0.252487 | 0.069074 | 0.298134 | 0.966282 |
| Jul | 0.103751 | 0.357103 | 0.257866 | 2.135991 |
| Aug | 0.100995 | 0.274159 | 0.196348 | 1.627422 |
| Sep | 0.0 | 0.358441 | 0.192327 | 1.271423 |
| Oct | 0.267967 | 0.147035 | 0.323297 | 2.034278 |
| Nov | 0.622504 | 0.234992 | 0.738586 | 1.423994 |
| Dec | 1.587383 | 0.824322 | 2.164089 | 1.16971 |2020
2021
2020
2021
Month
Month
Incidence per 1,000
Incidence per 100,000
c: Non-CAAP LRIs
d: Non-pneumonia IPD
#### Chart
| Category | Min | Min-Max bacteremic pneumonia 2016-2019 | Mean bacteremic pneumonia 2016-2019 | Bacteremic pneumonia 2020-2021 |
|---|---|---|---|---|
| Jan | 10.82587491291066 | 6.343240877626126 | 14.845552296945474 | 16.064879384265716 |
| Feb | 8.735730746556621 | 7.394127413103371 | 13.631649030197641 | 13.120512423162353 |
| Mar | 7.449488182646443 | 4.993545254805551 | 10.599150269711854 | 7.541711865282298 |
| Apr | 6.538399699876735 | 2.349142591695249 | 7.464159824774956 | 2.8410558396611396 |
| May | 7.170630712518306 | 0.6004181111056823 | 7.420405103986904 | 4.235755979131154 |
| Jun | 5.895278417921646 | 1.073362697060932 | 6.643094778162466 | 5.682111679322279 |
| Jul | 5.573717776944101 | 1.595108071793673 | 6.562212349923025 | 6.0953561650911725 |
| Aug | 4.513150386470923 | 1.6500952322653433 | 5.326009107038449 | 5.785422800764502 |
| Sep | 5.841684977758723 | 1.9928175569332733 | 6.450632198529942 | 7.541711865282298 |
| Oct | 6.225035015821963 | 1.9167029123873673 | 7.0472955795775425 | 5.010589389947827 |
| Nov | 7.39589474248352 | 1.9416577812494271 | 8.493481894374154 | 5.837078361485614 |
| Dec | 11.040248673562356 | 5.9748470363510116 | 13.632746713343938 | 7.954956351051191 |
| Jan | 10.82587491291066 | 6.343240877626126 | 14.845552296945474 | 6.967400701825764 |
| Feb | 8.735730746556621 | 7.394127413103371 | 13.631649030197641 | 4.983980064079744 |
| Mar | 7.449488182646443 | 4.993545254805551 | 10.599150269711854 | 6.153689670955602 |
| Apr | 6.538399699876735 | 2.349142591695249 | 7.464159824774956 | 7.831968672125312 |
| May | 7.170630712518306 | 0.6004181111056823 | 7.420405103986904 | 7.577683974978386 |
| Jun | 5.895278417921646 | 1.073362697060932 | 6.643094778162466 | 10.527386461882724 |
| Jul | 5.573717776944101 | 1.595108071793673 | 6.562212349923025 | 11.697096068758583 |
| Aug | 4.513150386470923 | 1.6500952322653433 | 5.326009107038449 | 11.442811371611658 |
| Sep | 5.841684977758723 | 1.9928175569332733 | 6.450632198529942 | 12.917662615063826 |
| Oct | 6.225035015821963 | 1.9167029123873673 | 7.0472955795775425 | 16.121649799115087 |
| Nov | 7.39589474248352 | 1.9416577812494271 | 8.493481894374154 | 16.070792859685707 |
| Dec | 11.040248673562356 | 5.9748470363510116 | 13.632746713343938 | 12.968519554493211 |
#### Chart
| Category | Min | Min-Max bacteremic pneumonia 2016-2019 | Mean bacteremic pneumonia 2016-2019 | Bacteremic pneumonia 2020-2021 |
|---|---|---|---|---|
| Jan | 1.0905125408942202 | 6.047278724573942 | 3.163919301486524 | 3.353828954723309 |
| Feb | 2.1990104452996153 | 3.816793893129771 | 2.885636932234772 | 3.353828954723309 |
| Mar | 0.0 | 4.456824512534819 | 2.3440220802276803 | 3.353828954723309 |
| Apr | 0.5571030640668524 | 3.289473684210526 | 1.9203390012295811 | 1.6769144773616544 |
| May | 1.6713091922005572 | 3.271537622682661 | 2.471148081912022 | 0.5589714924538849 |
| Jun | 0.5452562704471101 | 4.456824512534819 | 2.4852030752917917 | 1.1179429849077698 |
| Jul | 0.0 | 2.2284122562674096 | 1.2416627457137177 | 1.1179429849077698 |
| Aug | 0.0 | 1.0964912280701753 | 0.686026708257812 | 0.5589714924538849 |
| Sep | 1.0964912280701753 | 3.848268279274327 | 2.0599976793166617 | 1.1179429849077698 |
| Oct | 1.6357688113413305 | 2.741228070175439 | 2.0636667429397226 | 0.5589714924538849 |
| Nov | 3.271537622682661 | 4.385964912280701 | 3.713934912845188 | 1.6769144773616544 |
| Dec | 1.0964912280701753 | 5.013927576601671 | 2.7578035653323045 | 3.353828954723309 |
| Jan | 1.0905125408942202 | 6.047278724573942 | 3.163919301486524 | 2.230897936419409 |
| Feb | 2.1990104452996153 | 3.816793893129771 | 2.885636932234772 | 0.0 |
| Mar | 0.0 | 4.456824512534819 | 2.3440220802276803 | 1.6731734523145567 |
| Apr | 0.5571030640668524 | 3.289473684210526 | 1.9203390012295811 | 2.230897936419409 |
| May | 1.6713091922005572 | 3.271537622682661 | 2.471148081912022 | 1.6731734523145567 |
| Jun | 0.5452562704471101 | 4.456824512534819 | 2.4852030752917917 | 1.6731734523145567 |
| Jul | 0.0 | 2.2284122562674096 | 1.2416627457137177 | 1.6731734523145567 |
| Aug | 0.0 | 1.0964912280701753 | 0.686026708257812 | 0.5577244841048522 |
| Sep | 1.0964912280701753 | 3.848268279274327 | 2.0599976793166617 | 1.1154489682097044 |
| Oct | 1.6357688113413305 | 2.741228070175439 | 2.0636667429397226 | 3.3463469046291134 |
| Nov | 3.271537622682661 | 4.385964912280701 | 3.713934912845188 | 3.3463469046291134 |
| Dec | 1.0964912280701753 | 5.013927576601671 | 2.7578035653323045 | 2.7886224205242613 |2021
2020
2021
2020
Month
Month

### Slide 3
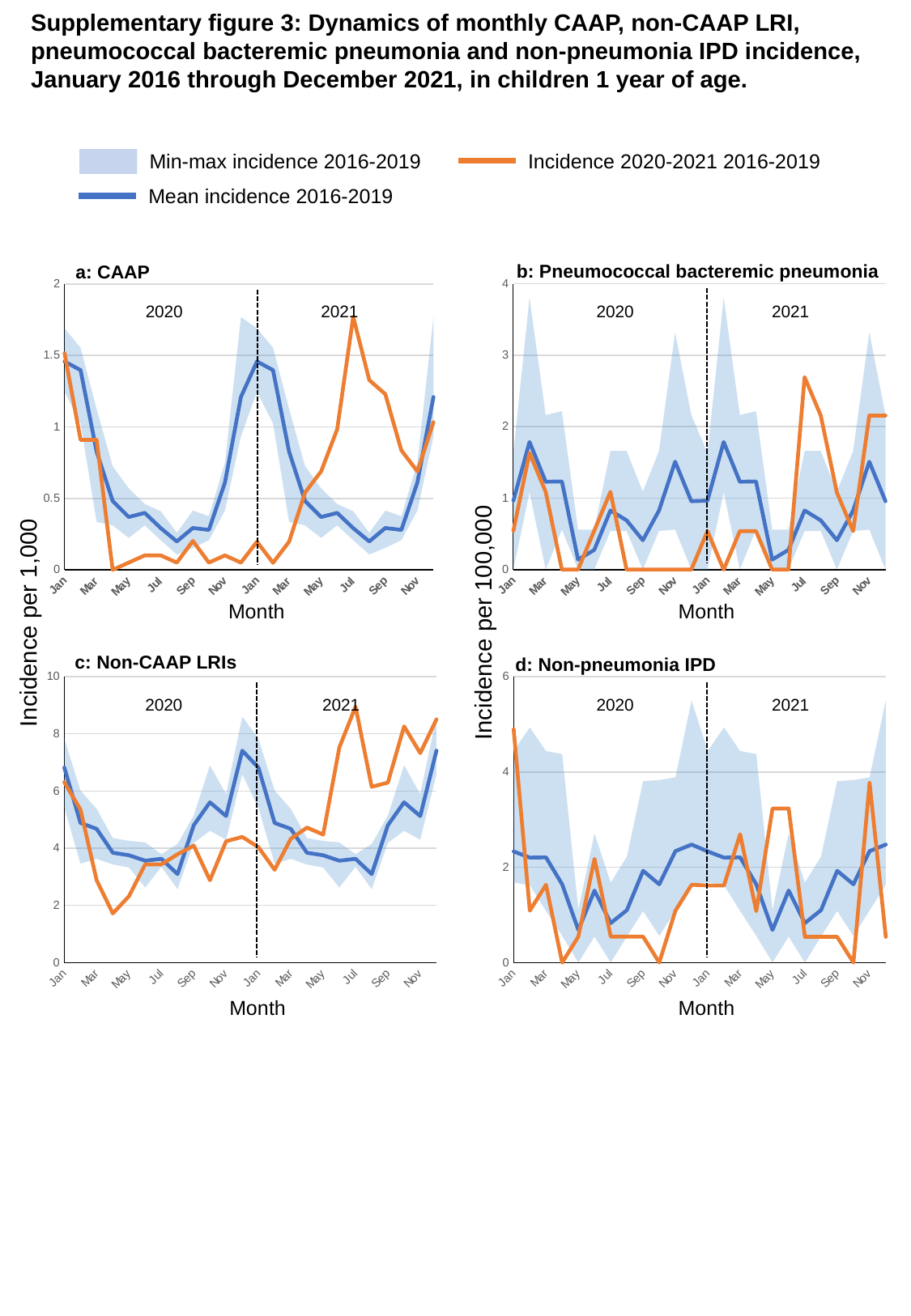

Supplementary figure 3: Dynamics of monthly CAAP, non-CAAP LRI,
pneumococcal bacteremic pneumonia and non-pneumonia IPD incidence,
January 2016 through December 2021, in children 1 year of age.
Min-max incidence 2016-2019
Incidence 2020-2021 2016-2019
Mean incidence 2016-2019
b: Pneumococcal bacteremic pneumonia
a: CAAP
#### Chart
| Category | Min | Min-Max bacteremic pneumonia 2016-2019 | Mean bacteremic pneumonia 2016-2019 | Bacteremic pneumonia 2020-2021 |
|---|---|---|---|---|
| Jan | 0.0 | 1.6357688113413305 | 0.9648313055858249 | 0.544069640914037 |
| Feb | 1.0816657652785289 | 2.7262813522355507 | 1.7858204335042582 | 1.632208922742111 |
| Mar | 0.0 | 2.1633315305570577 | 1.2295736305221627 | 1.088139281828074 |
| Apr | 0.5595970900951315 | 1.6565433462175592 | 1.2333937532331374 | 0.0 |
| May | 0.0 | 0.5595970900951315 | 0.13989927252378287 | 0.0 |
| Jun | 0.0 | 0.5595970900951315 | 0.27510749318359895 | 0.544069640914037 |
| Jul | 0.5408328826392644 | 1.119194180190263 | 0.8274113910220859 | 1.088139281828074 |
| Aug | 0.5408328826392644 | 1.119194180190263 | 0.6893661121706226 | 0.0 |
| Sep | 0.0 | 1.0905125408942202 | 0.41067341407501834 | 0.0 |
| Oct | 0.5408328826392644 | 1.119194180190263 | 0.8256801797824002 | 0.0 |
| Nov | 0.5595970900951315 | 2.760905577029266 | 1.509484310936064 | 0.0 |
| Dec | 0.0 | 2.1633315305570577 | 0.9569454952986077 | 0.0 |
| Jan | 0.0 | 1.6357688113413305 | 0.9648313055858249 | 0.5387931034482758 |
| Feb | 1.0816657652785289 | 2.7262813522355507 | 1.7858204335042582 | 0.0 |
| Mar | 0.0 | 2.1633315305570577 | 1.2295736305221627 | 0.5387931034482758 |
| Apr | 0.5595970900951315 | 1.6565433462175592 | 1.2333937532331374 | 0.5387931034482758 |
| May | 0.0 | 0.5595970900951315 | 0.13989927252378287 | 0.0 |
| Jun | 0.0 | 0.5595970900951315 | 0.27510749318359895 | 0.0 |
| Jul | 0.5408328826392644 | 1.119194180190263 | 0.8274113910220859 | 2.6939655172413794 |
| Aug | 0.5408328826392644 | 1.119194180190263 | 0.6893661121706226 | 2.155172413793103 |
| Sep | 0.0 | 1.0905125408942202 | 0.41067341407501834 | 1.0775862068965516 |
| Oct | 0.5408328826392644 | 1.119194180190263 | 0.8256801797824002 | 0.5387931034482758 |
| Nov | 0.5595970900951315 | 2.760905577029266 | 1.509484310936064 | 2.155172413793103 |
| Dec | 0.0 | 2.1633315305570577 | 0.9569454952986077 | 2.155172413793103 |
#### Chart
| Category | Min | Min-Max bacteremic pneumonia 2016-2019 | Mean bacteremic pneumonia 2016-2019 | Bacteremic pneumonia 2020-2021 |
|---|---|---|---|---|
| Jan | 1.245007 | 0.444788 | 1.458038 | 1.514922 |
| Feb | 1.024118 | 0.530092 | 1.397205 | 0.908953 |
| Mar | 0.334691 | 0.791839 | 0.827375 | 0.908953 |
| Apr | 0.311252 | 0.413911 | 0.481504 | 0.0 |
| May | 0.223127 | 0.347501 | 0.369633 | 0.050497 |
| Jun | 0.311252 | 0.149601 | 0.398378 | 0.100995 |
| Jul | 0.207501 | 0.202146 | 0.291553 | 0.100995 |
| Aug | 0.107187 | 0.15219 | 0.198629 | 0.050497 |
| Sep | 0.153618 | 0.261385 | 0.292819 | 0.20199 |
| Oct | 0.207501 | 0.167653 | 0.279398 | 0.050497 |
| Nov | 0.415002 | 0.335306 | 0.611147 | 0.100995 |
| Dec | 0.933755 | 0.834828 | 1.209379 | 0.050497 |
| Jan | 1.245007 | 0.444788 | 1.458038 | 0.196734 |
| Feb | 1.024118 | 0.530092 | 1.397205 | 0.049184 |
| Mar | 0.334691 | 0.791839 | 0.827375 | 0.196734 |
| Apr | 0.311252 | 0.413911 | 0.481504 | 0.541019 |
| May | 0.223127 | 0.347501 | 0.369633 | 0.68857 |
| Jun | 0.311252 | 0.149601 | 0.398378 | 0.983671 |
| Jul | 0.207501 | 0.202146 | 0.291553 | 1.770608 |
| Aug | 0.107187 | 0.15219 | 0.198629 | 1.327956 |
| Sep | 0.153618 | 0.261385 | 0.292819 | 1.229589 |
| Oct | 0.207501 | 0.167653 | 0.279398 | 0.83612 |
| Nov | 0.415002 | 0.335306 | 0.611147 | 0.68857 |
| Dec | 0.933755 | 0.834828 | 1.209379 | 1.032855 |2020
2021
2020
2021
Month
Month
Incidence per 1,000
Incidence per 100,000
c: Non-CAAP LRIs
d: Non-pneumonia IPD
#### Chart
| Category | Min | Min-Max bacteremic pneumonia 2016-2019 | Mean bacteremic pneumonia 2016-2019 | Bacteremic pneumonia 2020-2021 |
|---|---|---|---|---|
| Jan | 5.4108328220003346 | 2.4236697126916615 | 6.819457788164282 | 6.312174922991465 |
| Feb | 3.4584704635466057 | 2.5590633850812923 | 4.8835883307122945 | 5.352724334696763 |
| Mar | 3.6258158085569256 | 1.7508035779964057 | 4.677248708072899 | 2.878351764884109 |
| Apr | 3.42998017042714 | 0.9225212377350798 | 3.8386815134262795 | 1.7169115790536786 |
| May | 3.3227932901012918 | 0.9309806373770502 | 3.7513367986088304 | 2.3228803716608595 |
| Jun | 2.6217437384950073 | 1.5801548971848178 | 3.560726894460142 | 3.4338231581073573 |
| Jul | 3.3469069002063927 | 0.44232961984071695 | 3.6284500531152775 | 3.4338231581073573 |
| Aug | 2.565961956824901 | 1.5840613870564084 | 3.0937874027900274 | 3.7873049537948793 |
| Sep | 4.183633625257991 | 0.9613366303827187 | 4.805764851018921 | 4.09028935009847 |
| Oct | 4.609035854011469 | 2.2903779551912082 | 5.60666003869343 | 2.878351764884109 |
| Nov | 4.295197188598204 | 1.6000812293234423 | 5.125150714240981 | 4.241781548250265 |
| Dec | 6.588162058411578 | 2.014428960073751 | 7.41390392523342 | 4.393273746402061 |
| Jan | 5.4108328220003346 | 2.4236697126916615 | 6.819457788164282 | 4.033051347629352 |
| Feb | 3.4584704635466057 | 2.5590633850812923 | 4.8835883307122945 | 3.2461144993114304 |
| Mar | 3.6258158085569256 | 1.7508035779964057 | 4.677248708072899 | 4.328152665748574 |
| Apr | 3.42998017042714 | 0.9225212377350798 | 3.8386815134262795 | 4.721621089907535 |
| May | 3.3227932901012918 | 0.9309806373770502 | 3.7513367986088304 | 4.475703324808184 |
| Jun | 2.6217437384950073 | 1.5801548971848178 | 3.560726894460142 | 7.525083612040134 |
| Jul | 3.3469069002063927 | 0.44232961984071695 | 3.6284500531152775 | 8.951406649616368 |
| Aug | 2.565961956824901 | 1.5840613870564084 | 3.0937874027900274 | 6.147944127483769 |
| Sep | 4.183633625257991 | 0.9613366303827187 | 4.805764851018921 | 6.2954947865433795 |
| Oct | 4.609035854011469 | 2.2903779551912082 | 5.60666003869343 | 8.262836907338187 |
| Nov | 4.295197188598204 | 1.6000812293234423 | 5.125150714240981 | 7.328349399960652 |
| Dec | 6.588162058411578 | 2.014428960073751 | 7.41390392523342 | 8.508754672437536 |
#### Chart
| Category | Min | Min-Max bacteremic pneumonia 2016-2019 | Mean bacteremic pneumonia 2016-2019 | Bacteremic pneumonia 2020-2021 |
|---|---|---|---|---|
| Jan | 1.6787912702853944 | 2.760905577029266 | 2.331221585574856 | 4.896626768226333 |
| Feb | 1.6224986479177936 | 3.3130866924351183 | 2.202435628018692 | 1.088139281828074 |
| Mar | 1.0816657652785289 | 3.357582540570789 | 2.2072494623152927 | 1.632208922742111 |
| Apr | 0.5595970900951315 | 3.816793893129771 | 1.6406047448287844 | 0.0 |
| May | 0.0 | 1.1043622308117063 | 0.6839269135862976 | 0.544069640914037 |
| Jun | 0.5452562704471101 | 2.1633315305570577 | 1.5109806043767804 | 2.176278563656148 |
| Jul | 0.0 | 1.6787912702853944 | 0.828159537742444 | 0.544069640914037 |
| Aug | 0.5452562704471101 | 1.6787912702853944 | 1.0996818260140377 | 0.544069640914037 |
| Sep | 1.0816657652785289 | 2.7262813522355507 | 1.925719706028041 | 0.544069640914037 |
| Oct | 0.5595970900951315 | 3.271537622682661 | 1.64233595606847 | 0.0 |
| Nov | 1.0905125408942202 | 2.797985450475658 | 2.3383919953988666 | 1.088139281828074 |
| Dec | 1.6357688113413305 | 3.865267807840972 | 2.4756891275299715 | 1.632208922742111 |
| Jan | 1.6787912702853944 | 2.760905577029266 | 2.331221585574856 | 1.6163793103448276 |
| Feb | 1.6224986479177936 | 3.3130866924351183 | 2.202435628018692 | 1.6163793103448276 |
| Mar | 1.0816657652785289 | 3.357582540570789 | 2.2072494623152927 | 2.6939655172413794 |
| Apr | 0.5595970900951315 | 3.816793893129771 | 1.6406047448287844 | 1.0775862068965516 |
| May | 0.0 | 1.1043622308117063 | 0.6839269135862976 | 3.2327586206896552 |
| Jun | 0.5452562704471101 | 2.1633315305570577 | 1.5109806043767804 | 3.2327586206896552 |
| Jul | 0.0 | 1.6787912702853944 | 0.828159537742444 | 0.5387931034482758 |
| Aug | 0.5452562704471101 | 1.6787912702853944 | 1.0996818260140377 | 0.5387931034482758 |
| Sep | 1.0816657652785289 | 2.7262813522355507 | 1.925719706028041 | 0.5387931034482758 |
| Oct | 0.5595970900951315 | 3.271537622682661 | 1.64233595606847 | 0.0 |
| Nov | 1.0905125408942202 | 2.797985450475658 | 2.3383919953988666 | 3.771551724137931 |
| Dec | 1.6357688113413305 | 3.865267807840972 | 2.4756891275299715 | 0.5387931034482758 |2021
2020
2021
2020
Month
Month

### Slide 4
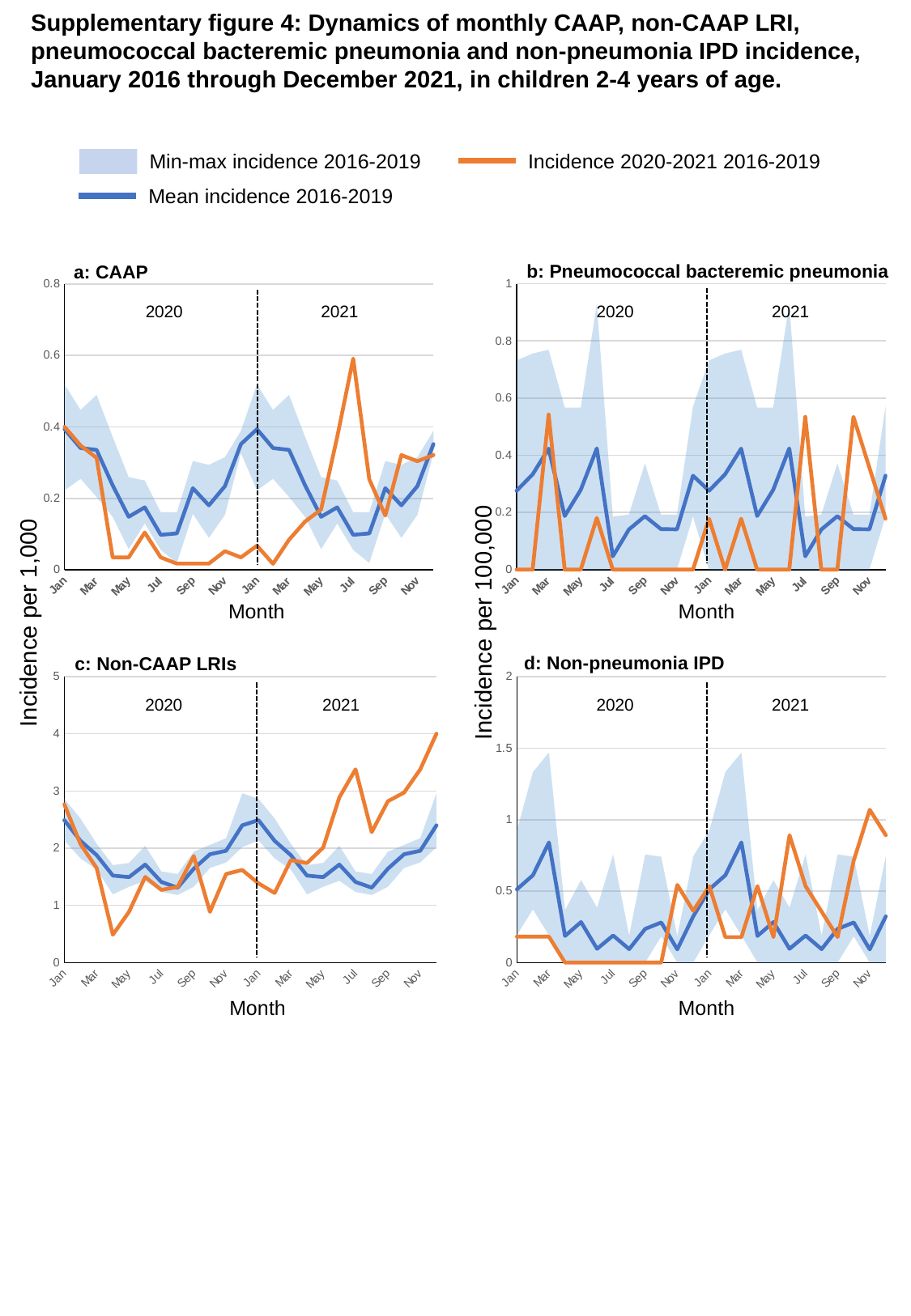

Supplementary figure 4: Dynamics of monthly CAAP, non-CAAP LRI,
pneumococcal bacteremic pneumonia and non-pneumonia IPD incidence,
January 2016 through December 2021, in children 2-4 years of age.
Min-max incidence 2016-2019
Incidence 2020-2021 2016-2019
Mean incidence 2016-2019
b: Pneumococcal bacteremic pneumonia
a: CAAP
#### Chart
| Category | Min | Min-Max bacteremic pneumonia 2016-2019 | Mean bacteremic pneumonia 2016-2019 | Bacteremic pneumonia 2020-2021 |
|---|---|---|---|---|
| Jan | 0.0 | 0.7316627034936893 | 0.27585247884740005 | 0.0 |
| Feb | 0.0 | 0.756000756000756 | 0.3332309582309582 | 0.0 |
| Mar | 0.0 | 0.7692307692307692 | 0.4231707347053699 | 0.5427899402931066 |
| Apr | 0.0 | 0.567000567000567 | 0.18747906071849735 | 0.0 |
| May | 0.0 | 0.567000567000567 | 0.27893689865520854 | 0.0 |
| Jun | 0.0 | 0.929368029739777 | 0.4238228239157607 | 0.1809299800977022 |
| Jul | 0.0 | 0.1858736059479554 | 0.04646840148698885 | 0.0 |
| Aug | 0.0 | 0.1923076923076923 | 0.14027424353226753 | 0.0 |
| Sep | 0.0 | 0.3717472118959108 | 0.18674264501925636 | 0.0 |
| Oct | 0.0 | 0.1923076923076923 | 0.14179537181395918 | 0.0 |
| Nov | 0.0 | 0.1923076923076923 | 0.14105588929532592 | 0.0 |
| Dec | 0.1858736059479554 | 0.3846153846153846 | 0.32858018007764067 | 0.0 |
| Jan | 0.0 | 0.7316627034936893 | 0.27585247884740005 | 0.17815784785319794 |
| Feb | 0.0 | 0.756000756000756 | 0.3332309582309582 | 0.0 |
| Mar | 0.0 | 0.7692307692307692 | 0.4231707347053699 | 0.17815784785319794 |
| Apr | 0.0 | 0.567000567000567 | 0.18747906071849735 | 0.0 |
| May | 0.0 | 0.567000567000567 | 0.27893689865520854 | 0.0 |
| Jun | 0.0 | 0.929368029739777 | 0.4238228239157607 | 0.0 |
| Jul | 0.0 | 0.1858736059479554 | 0.04646840148698885 | 0.5344735435595939 |
| Aug | 0.0 | 0.1923076923076923 | 0.14027424353226753 | 0.0 |
| Sep | 0.0 | 0.3717472118959108 | 0.18674264501925636 | 0.0 |
| Oct | 0.0 | 0.1923076923076923 | 0.14179537181395918 | 0.5344735435595939 |
| Nov | 0.0 | 0.1923076923076923 | 0.14105588929532592 | 0.3563156957063959 |
| Dec | 0.1858736059479554 | 0.3846153846153846 | 0.32858018007764067 | 0.17815784785319794 |
#### Chart
| Category | Min | Min-Max bacteremic pneumonia 2016-2019 | Mean bacteremic pneumonia 2016-2019 | Bacteremic pneumonia 2020-2021 |
|---|---|---|---|---|
| Jan | 0.222436 | 0.296691 | 0.3937 | 0.400244 |
| Feb | 0.254652 | 0.192871 | 0.340839 | 0.348038 |
| Mar | 0.2039 | 0.285816 | 0.335667 | 0.313234 |
| Apr | 0.148291 | 0.223893 | 0.236361 | 0.034804 |
| May | 0.057677 | 0.201832 | 0.148535 | 0.034804 |
| Jun | 0.129755 | 0.120178 | 0.174798 | 0.104411 |
| Jul | 0.055609 | 0.105499 | 0.097982 | 0.034804 |
| Aug | 0.019226 | 0.141883 | 0.102008 | 0.017402 |
| Sep | 0.156709 | 0.147607 | 0.228542 | 0.017402 |
| Oct | 0.089505 | 0.204325 | 0.180453 | 0.017402 |
| Nov | 0.153805 | 0.161314 | 0.234175 | 0.052206 |
| Dec | 0.326835 | 0.062429 | 0.351781 | 0.034804 |
| Jan | 0.222436 | 0.296691 | 0.3937 | 0.067574 |
| Feb | 0.254652 | 0.192871 | 0.340839 | 0.016894 |
| Mar | 0.2039 | 0.285816 | 0.335667 | 0.084468 |
| Apr | 0.148291 | 0.223893 | 0.236361 | 0.135149 |
| May | 0.057677 | 0.201832 | 0.148535 | 0.168936 |
| Jun | 0.129755 | 0.120178 | 0.174798 | 0.371659 |
| Jul | 0.055609 | 0.105499 | 0.097982 | 0.591276 |
| Aug | 0.019226 | 0.141883 | 0.102008 | 0.253404 |
| Sep | 0.156709 | 0.147607 | 0.228542 | 0.152042 |
| Oct | 0.089505 | 0.204325 | 0.180453 | 0.320978 |
| Nov | 0.153805 | 0.161314 | 0.234175 | 0.304085 |
| Dec | 0.326835 | 0.062429 | 0.351781 | 0.320978 |2020
2021
2020
2021
Month
Month
Incidence per 1,000
Incidence per 100,000
d: Non-pneumonia IPD
c: Non-CAAP LRIs
#### Chart
| Category | Min | Min-Max bacteremic pneumonia 2016-2019 | Mean bacteremic pneumonia 2016-2019 | Bacteremic pneumonia 2020-2021 |
|---|---|---|---|---|
| Jan | 2.1351616062683645 | 0.728988188765916 | 2.490821567991416 | 2.766901592273558 |
| Feb | 1.8217433888344758 | 0.6992027074063487 | 2.128867033014302 | 2.0708257200034805 |
| Mar | 1.6258570029382957 | 0.45065159846155733 | 1.8782193971135421 | 1.6531801966414341 |
| Apr | 1.1919867727919407 | 0.5122247845048273 | 1.5205596169525646 | 0.4872531105890542 |
| May | 1.3246692802033546 | 0.4177493451395682 | 1.494175775265851 | 0.8874967371443487 |
| Jun | 1.4299706170421156 | 0.6107361119198091 | 1.713023989735586 | 1.4965631253806666 |
| Jul | 1.2340842311459355 | 0.35909909234188286 | 1.408668818326244 | 1.2703384668928912 |
| Aug | 1.1814617904516405 | 0.3660406581281832 | 1.3094305974406972 | 1.3225441573131471 |
| Sep | 1.3246692802033546 | 0.6146059401688295 | 1.6403921233818244 | 1.862002958322457 |
| Oct | 1.653401007421079 | 0.403406044488813 | 1.8932994440690987 | 0.8874967371443487 |
| Nov | 1.7495289729688162 | 0.4248099104787839 | 1.9548705712802579 | 1.5487688158009223 |
| Dec | 2.0176297747306564 | 0.9431115641396475 | 2.400365192376901 | 1.61837640302793 |
| Jan | 2.1351616062683645 | 0.728988188765916 | 2.490821567991416 | 1.3852755346825691 |
| Feb | 1.8217433888344758 | 0.6992027074063487 | 2.128867033014302 | 1.2163394938676217 |
| Mar | 1.6258570029382957 | 0.45065159846155733 | 1.8782193971135421 | 1.7907220326384432 |
| Apr | 1.1919867727919407 | 0.5122247845048273 | 1.5205596169525646 | 1.7400412203939588 |
| May | 1.3246692802033546 | 0.4177493451395682 | 1.494175775265851 | 2.010338885697875 |
| Jun | 1.4299706170421156 | 0.6107361119198091 | 1.713023989735586 | 2.8888062979356013 |
| Jul | 1.2340842311459355 | 0.35909909234188286 | 1.408668818326244 | 3.3787208162989493 |
| Aug | 1.1814617904516405 | 0.3660406581281832 | 1.3094305974406972 | 2.2806365510017907 |
| Sep | 1.3246692802033546 | 0.6146059401688295 | 1.6403921233818244 | 2.8212318816096227 |
| Oct | 1.653401007421079 | 0.403406044488813 | 1.8932994440690987 | 2.973274318343075 |
| Nov | 1.7495289729688162 | 0.4248099104787839 | 1.9548705712802579 | 3.3787208162989493 |
| Dec | 2.0176297747306564 | 0.9431115641396475 | 2.400365192376901 | 4.003784167314255 |
#### Chart
| Category | Min | Min-Max bacteremic pneumonia 2016-2019 | Mean bacteremic pneumonia 2016-2019 | Bacteremic pneumonia 2020-2021 |
|---|---|---|---|---|
| Jan | 0.1923076923076923 | 0.7316627034936893 | 0.5121479451614537 | 0.1809299800977022 |
| Feb | 0.3717472118959108 | 0.9615384615384616 | 0.6107371887321099 | 0.1809299800977022 |
| Mar | 0.189000189000189 | 1.2804097311139564 | 0.8400791028480961 | 0.1809299800977022 |
| Apr | 0.0 | 0.36583135174684467 | 0.1860031625006231 | 0.0 |
| May | 0.0 | 0.5769230769230769 | 0.28293865441752764 | 0.0 |
| Jun | 0.0 | 0.3846153846153846 | 0.09615384615384615 | 0.0 |
| Jul | 0.0 | 0.756000756000756 | 0.189000189000189 | 0.0 |
| Aug | 0.0 | 0.189000189000189 | 0.09371844873703611 | 0.0 |
| Sep | 0.0 | 0.756000756000756 | 0.23546859048717786 | 0.0 |
| Oct | 0.18291567587342233 | 0.5576208178438661 | 0.2804610937562924 | 0.0 |
| Nov | 0.0 | 0.1858736059479554 | 0.09219732045534443 | 0.5427899402931066 |
| Dec | 0.0 | 0.7434944237918216 | 0.3230603628530222 | 0.3618599601954044 |
| Jan | 0.1923076923076923 | 0.7316627034936893 | 0.5121479451614537 | 0.5344735435595939 |
| Feb | 0.3717472118959108 | 0.9615384615384616 | 0.6107371887321099 | 0.17815784785319794 |
| Mar | 0.189000189000189 | 1.2804097311139564 | 0.8400791028480961 | 0.17815784785319794 |
| Apr | 0.0 | 0.36583135174684467 | 0.1860031625006231 | 0.5344735435595939 |
| May | 0.0 | 0.5769230769230769 | 0.28293865441752764 | 0.17815784785319794 |
| Jun | 0.0 | 0.3846153846153846 | 0.09615384615384615 | 0.8907892392659896 |
| Jul | 0.0 | 0.756000756000756 | 0.189000189000189 | 0.5344735435595939 |
| Aug | 0.0 | 0.189000189000189 | 0.09371844873703611 | 0.3563156957063959 |
| Sep | 0.0 | 0.756000756000756 | 0.23546859048717786 | 0.17815784785319794 |
| Oct | 0.18291567587342233 | 0.5576208178438661 | 0.2804610937562924 | 0.7126313914127917 |
| Nov | 0.0 | 0.1858736059479554 | 0.09219732045534443 | 1.0689470871191877 |
| Dec | 0.0 | 0.7434944237918216 | 0.3230603628530222 | 0.8907892392659896 |2021
2020
2021
2020
Month
Month

### Slide 5
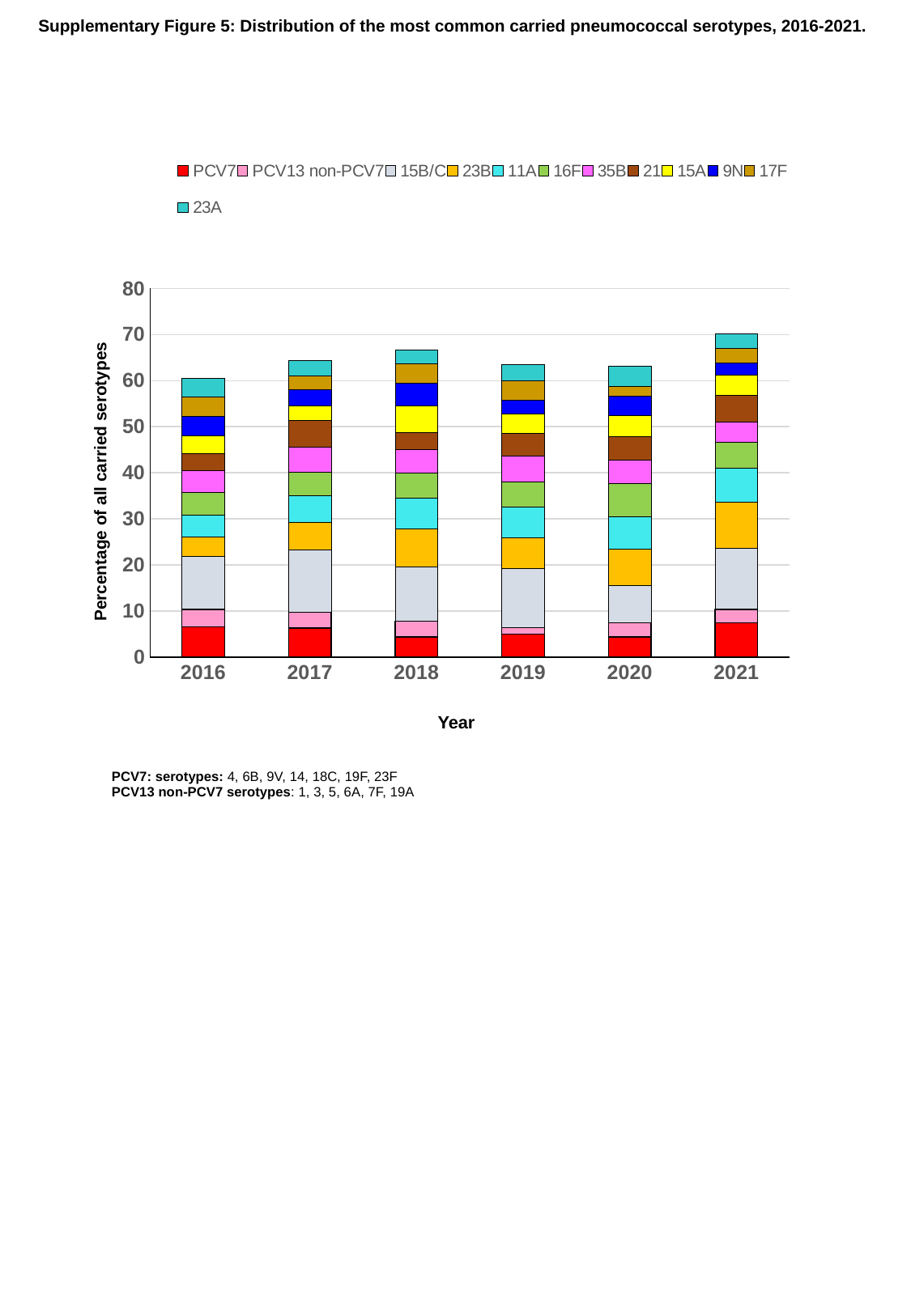

Supplementary Figure 5: Distribution of the most common carried pneumococcal serotypes, 2016-2021.
#### Chart
| Category | PCV7 | PCV13 non-PCV7 | 15B/C | 23B | 11A | 16F | 35B | 21 | 15A | 9N | 17F | 23A |
|---|---|---|---|---|---|---|---|---|---|---|---|---|
| 2016 | 6.6 | 3.8 | 11.4 | 4.2 | 4.9 | 4.9 | 4.7 | 3.6 | 3.9 | 4.2 | 4.3 | 4.0 |
| 2017 | 6.3 | 3.5 | 13.4 | 6.1 | 5.7 | 5.2 | 5.3 | 5.8 | 3.3 | 3.4 | 3.0 | 3.3 |
| 2018 | 4.4 | 3.4 | 11.8 | 8.2 | 6.8 | 5.4 | 5.0 | 3.8 | 5.8 | 4.9 | 4.2 | 2.9 |
| 2019 | 5.0 | 1.4 | 12.9 | 6.6 | 6.6 | 5.6 | 5.5 | 5.0 | 4.1 | 3.1 | 4.1 | 3.6 |
| 2020 | 4.4 | 3.1 | 8.11 | 7.9 | 6.9 | 7.3 | 5.0 | 5.2 | 4.6 | 4.2 | 2.1 | 4.4 |
| 2021 | 7.5 | 2.9 | 13.26 | 10.0 | 7.4 | 5.6 | 4.3 | 5.9 | 4.3 | 2.7 | 3.2 | 3.1 |Percentage of all carried serotypes
Year
PCV7: serotypes: 4, 6B, 9V, 14, 18C, 19F, 23F
PCV13 non-PCV7 serotypes: 1, 3, 5, 6A, 7F, 19A

### Slide 6
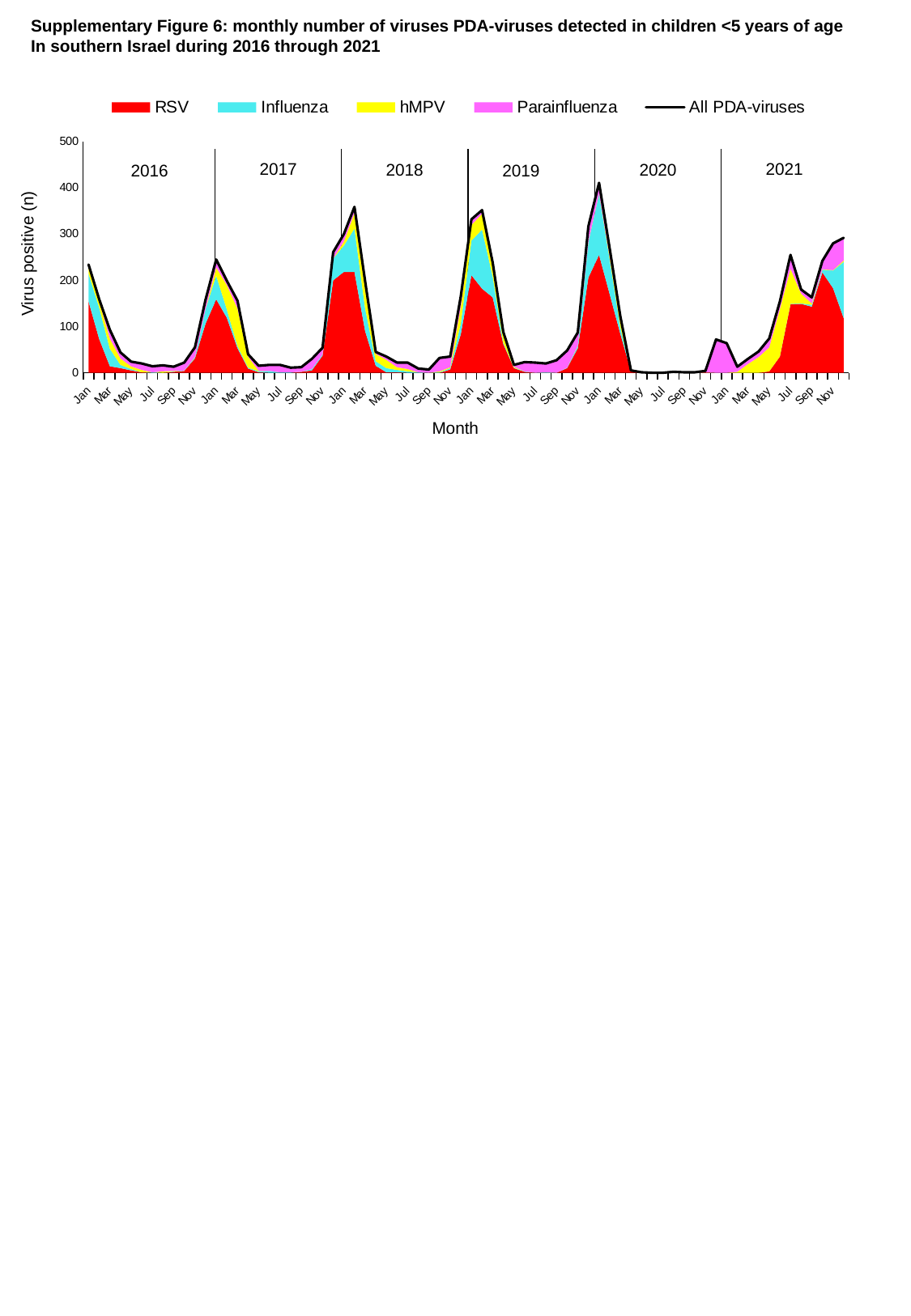

Supplementary Figure 6: monthly number of viruses PDA-viruses detected in children <5 years of age
In southern Israel during 2016 through 2021
#### Chart
| Category | RSV | Influenza | hMPV | Parainfluenza | All PDA-viruses |
|---|---|---|---|---|---|
| Jan | 154.0 | 60.0 | 15.0 | 5.0 | 234.0 |
| Feb | 73.0 | 68.0 | 14.0 | 4.0 | 159.0 |
| Mar | 14.0 | 38.0 | 22.0 | 19.0 | 93.0 |
| Apr | 10.0 | 7.0 | 14.0 | 13.0 | 44.0 |
| May | 5.0 | 3.0 | 6.0 | 10.0 | 24.0 |
| Jun | 1.0 | 0.0 | 5.0 | 14.0 | 20.0 |
| Jul | 0.0 | 1.0 | 0.0 | 13.0 | 14.0 |
| Aug | 0.0 | 0.0 | 3.0 | 13.0 | 16.0 |
| Sep | 2.0 | 1.0 | 1.0 | 9.0 | 13.0 |
| Oct | 4.0 | 1.0 | 0.0 | 17.0 | 22.0 |
| Nov | 31.0 | 2.0 | 1.0 | 21.0 | 55.0 |
| Dec | 106.0 | 32.0 | 0.0 | 20.0 | 158.0 |
| Jan | 159.0 | 51.0 | 18.0 | 17.0 | 245.0 |
| Feb | 119.0 | 18.0 | 53.0 | 9.0 | 199.0 |
| Mar | 53.0 | 5.0 | 81.0 | 17.0 | 156.0 |
| Apr | 9.0 | 2.0 | 24.0 | 5.0 | 40.0 |
| May | 1.0 | 2.0 | 1.0 | 11.0 | 15.0 |
| Jun | 0.0 | 4.0 | 0.0 | 13.0 | 17.0 |
| Jul | 0.0 | 1.0 | 0.0 | 16.0 | 17.0 |
| Aug | 0.0 | 1.0 | 0.0 | 10.0 | 11.0 |
| Sep | 2.0 | 0.0 | 0.0 | 10.0 | 12.0 |
| Oct | 5.0 | 3.0 | 0.0 | 22.0 | 30.0 |
| Nov | 37.0 | 3.0 | 0.0 | 14.0 | 54.0 |
| Dec | 200.0 | 47.0 | 0.0 | 14.0 | 261.0 |
| Jan | 218.0 | 57.0 | 9.0 | 16.0 | 300.0 |
| Feb | 218.0 | 94.0 | 34.0 | 13.0 | 359.0 |
| Mar | 90.0 | 49.0 | 44.0 | 15.0 | 198.0 |
| Apr | 15.0 | 10.0 | 16.0 | 4.0 | 45.0 |
| May | 1.0 | 9.0 | 18.0 | 7.0 | 35.0 |
| Jun | 1.0 | 5.0 | 6.0 | 10.0 | 22.0 |
| Jul | 0.0 | 3.0 | 5.0 | 14.0 | 22.0 |
| Aug | 0.0 | 1.0 | 0.0 | 8.0 | 9.0 |
| Sep | 0.0 | 0.0 | 0.0 | 7.0 | 7.0 |
| Oct | 0.0 | 1.0 | 2.0 | 29.0 | 32.0 |
| Nov | 7.0 | 3.0 | 2.0 | 23.0 | 35.0 |
| Dec | 83.0 | 23.0 | 37.0 | 22.0 | 165.0 |
| Jan | 211.0 | 75.0 | 34.0 | 12.0 | 332.0 |
| Feb | 182.0 | 128.0 | 34.0 | 8.0 | 352.0 |
| Mar | 163.0 | 45.0 | 23.0 | 7.0 | 238.0 |
| Apr | 62.0 | 3.0 | 18.0 | 5.0 | 88.0 |
| May | 10.0 | 1.0 | 2.0 | 4.0 | 17.0 |
| Jun | 2.0 | 1.0 | 0.0 | 20.0 | 23.0 |
| Jul | 0.0 | 0.0 | 0.0 | 22.0 | 22.0 |
| Aug | 0.0 | 1.0 | 0.0 | 19.0 | 20.0 |
| Sep | 0.0 | 0.0 | 0.0 | 27.0 | 27.0 |
| Oct | 10.0 | 1.0 | 0.0 | 37.0 | 48.0 |
| Nov | 53.0 | 5.0 | 1.0 | 28.0 | 87.0 |
| Dec | 206.0 | 77.0 | 1.0 | 33.0 | 317.0 |
| Jan | 255.0 | 132.0 | 1.0 | 23.0 | 411.0 |
| Feb | 170.0 | 75.0 | 6.0 | 18.0 | 269.0 |
| Mar | 84.0 | 14.0 | 14.0 | 10.0 | 122.0 |
| Apr | 3.0 | 0.0 | 2.0 | 0.0 | 5.0 |
| May | 0.0 | 0.0 | 1.0 | 0.0 | 1.0 |
| Jun | 0.0 | 0.0 | 0.0 | 0.0 | 0.0 |
| Jul | 0.0 | 0.0 | 0.0 | 0.0 | 0.0 |
| Aug | 2.0 | 0.0 | 0.0 | 0.0 | 2.0 |
| Sep | 1.0 | 0.0 | 0.0 | 0.0 | 1.0 |
| Oct | 0.0 | 0.0 | 0.0 | 1.0 | 1.0 |
| Nov | 1.0 | 0.0 | 0.0 | 3.0 | 4.0 |
| Dec | 0.0 | 0.0 | 0.0 | 72.0 | 72.0 |
| Jan | 0.0 | 0.0 | 0.0 | 64.0 | 64.0 |
| Feb | 0.0 | 0.0 | 2.0 | 11.0 | 13.0 |
| Mar | 0.0 | 0.0 | 19.0 | 11.0 | 30.0 |
| Apr | 0.0 | 0.0 | 34.0 | 12.0 | 46.0 |
| May | 3.0 | 0.0 | 52.0 | 19.0 | 74.0 |
| Jun | 35.0 | 0.0 | 103.0 | 16.0 | 154.0 |
| Jul | 148.0 | 1.0 | 76.0 | 30.0 | 255.0 |
| Aug | 149.0 | 1.0 | 21.0 | 9.0 | 180.0 |
| Sep | 143.0 | 4.0 | 3.0 | 13.0 | 163.0 |
| Oct | 217.0 | 6.0 | 1.0 | 18.0 | 242.0 |
| Nov | 183.0 | 38.0 | 1.0 | 58.0 | 280.0 |
| Dec | 118.0 | 122.0 | 3.0 | 49.0 | 292.0 |2017
2021
2018
2020
2016
2019
Virus positive (n)
Month

### Slide 7
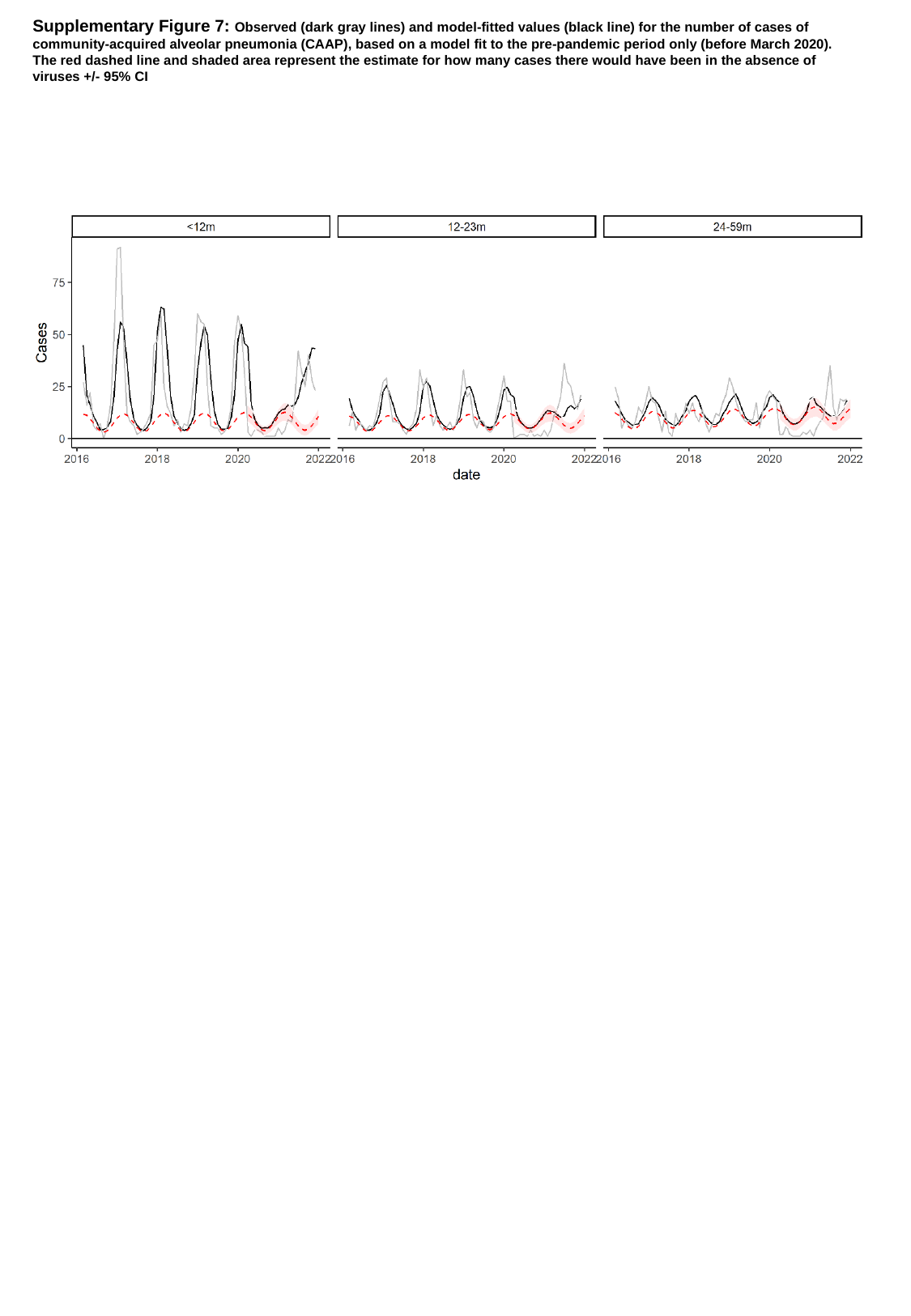

Supplementary Figure 7: Observed (dark gray lines) and model-fitted values (black line) for the number of cases of
community-acquired alveolar pneumonia (CAAP), based on a model fit to the pre-pandemic period only (before March 2020).
The red dashed line and shaded area represent the estimate for how many cases there would have been in the absence of
viruses +/- 95% CI

### Slide 8
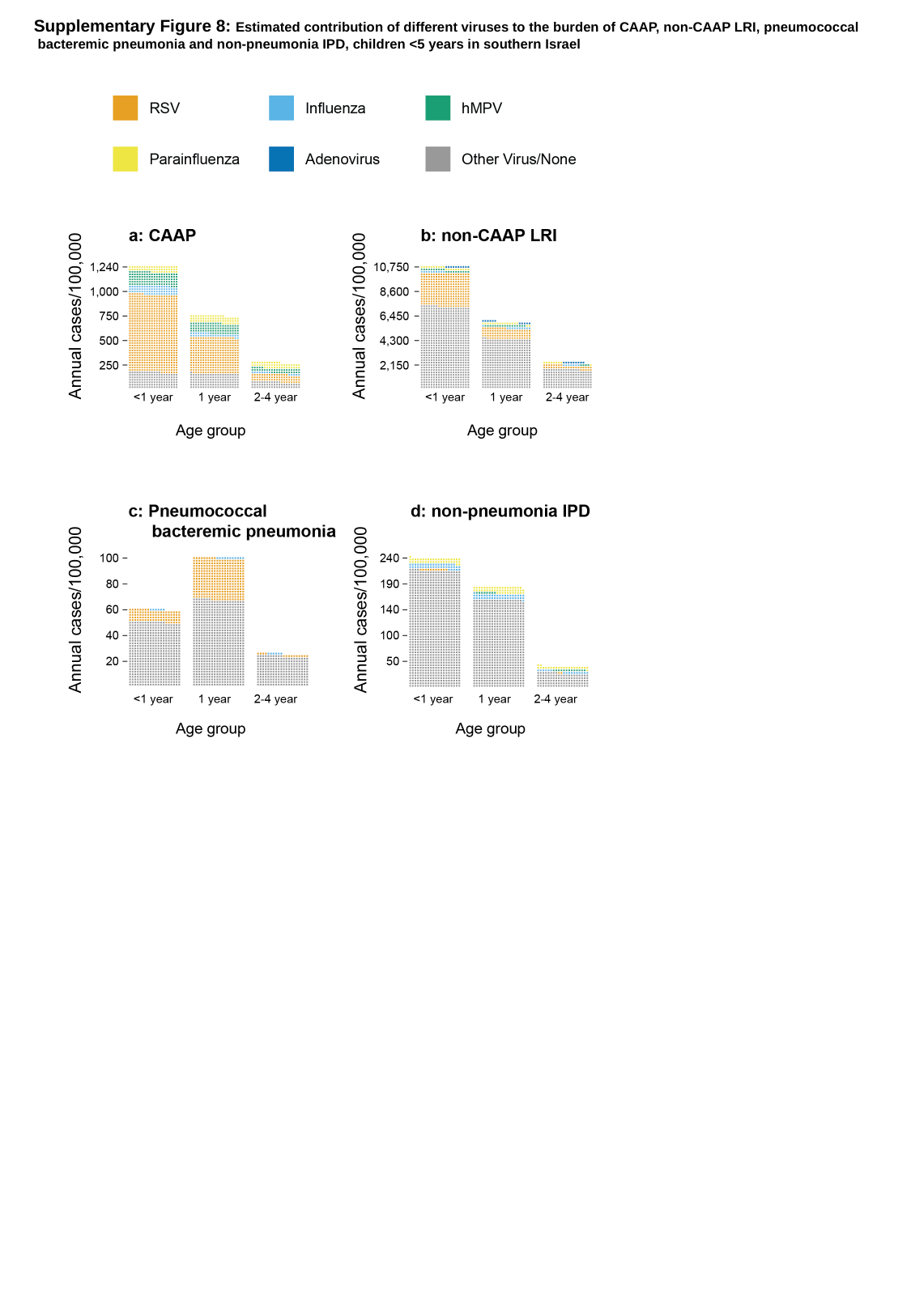

Supplementary Figure 8: Estimated contribution of different viruses to the burden of CAAP, non-CAAP LRI, pneumococcal
 bacteremic pneumonia and non-pneumonia IPD, children <5 years in southern Israel
